# Genetic landscape of idiopathic pulmonary fibrosis: A systematic review, meta-analysis and epidemiological evidence of case-control studies

**DOI:** 10.1101/2023.05.18.23290164

**Authors:** Pooja Singh, Debleena Guin, Bijay Pattnaik, Ritushree Kukreti

**Affiliations:** Academy of Scientific and Innovative Research (AcSIR), Ghaziabad, India; Centre of Excellence for Translational Research in Asthma and Lung Diseases, CSIR-Institute of Genomics and Integrative Biology, New Delhi, India; Department of Bioinformatics, Delhi Technological University, Shahbad Daulatpur, Main Bawana Road, New Delhi, India; Department of Pulmonary, Critical Care and Sleep Medicine, All India Institute of Medical Sciences (AIIMS), New Delhi, India; Genomics and Molecular Medicine Unit, Institute of Genomics and Integrative Biology (IGIB), Council of Scientific and Industrial Research (CSIR), New Delhi, India

**Keywords:** Idiopathic pulmonary fibrosis, Meta-analysis, Systematic review, Gene polymorphism, *MUC5B*, *DSP*, *TERT*, Venice criteria

## Abstract

**Background:** Idiopathic pulmonary fibrosis (IPF) is a fibrotic lung disorder of unknown cause, affecting about three million people worldwide. Being a multifactorial disease, complex genetic and environmental factors contribute to its susceptibility. Therefore, we conducted a two-staged systematic literature search and meta-analyses of published genetic association studies on IPF.

**Methods:** The first search was performed using PubMed and Web of Science, retrieving a total of 5642 articles, of which 57 publications were eligible for inclusion in the first stage. The Second search was performed using PubMed, Web of Science and Scopus for all genetic variants, identified from the first search, with 2 or more studies. Thus, six variants [rs35705950 (*MUC5B*), rs2736100 (*TERT*), rs2609255 (*FAM13A*), rs2076295 (*DSP*), rs12610495 (*DPP9*) and rs1800470 (*TGF-β1*)] from this search qualified for meta-analyses. Additionally, the epidemiological credibility of these six variants was evaluated using the Venice criteria.

**Results:** In this systematic review, 291 polymorphisms were found to be associated with IPF susceptibility. Meta-analyses findings revealed significant (p < 0.05) risk association of rs35705950 [T vs C; OR = 3.85(3.24-4.47)], rs2609255 [G vs T; OR = 1.37(1.27-1.47)], rs2076295 [G vs T; OR = 1.31(1.00-1.63)], rs12610495 [G vs A; OR = 1.29(1.21-1.37)] and rs1800470 [T vs C; OR = 1.08(0.82-1.34)] and protective association of rs2736100 [C vs A; OR = 0.70(0.61-0.79)] with IPF susceptibility. Cumulative epidemiological evidence was graded as strong for rs35705950, moderate for rs2736100, rs2609255 and rs12610495, and weak for rs2076295 and rs1800470.

**Conclusions:** Our findings present the most prominent IPF-associated genetic risk variants involved in alveolar epithelial injuries (*MUC5B, TERT, FAM13A, DSP, DPP9*) and epithelial-mesenchymal transition (*TGF-β1*), providing genetic and biological insights into IPF pathogenesis.

## 1. INTRODUCTION

Idiopathic pulmonary fibrosis (IPF) is one of the most common and life-threatening form of interstitial lung disease (ILD), mainly characterised by irreversible lung scarring occurring in the lung interstitium.^1^ It is an age-related disease with most patients older than 60 years at the time of diagnosis. IPF is associated with progressive decline in lung function leading to respiratory failure with a median survival of 3-5 years after diagnosis.^2–4^ IPF patients show high mortality due to the unavailability of effective medical treatment options, with only two food and drug administration approved drugs available (Pirfenidone and Nintedanib), both decelerating disease progression.^5^ As the name suggests, the aetiology of IPF is unknown and is currently proposed to result from repetitive micro-injury of alveolar epithelium coupled with an aberrant repair process which leads to excessive accumulation of extracellular matrix (ECM) that subsequently remodels the lung architecture.^6^

Being a complex genetic disorder, IPF is believed to result from a complex interaction between multiple genes with environmental factors. Initial proof favouring the hypothesis that genetic factors can contribute to IPF development comes from initial investigations that PF can also occur with several rare genetic disorders,^7, 8^ familial clustering of the disease, and evidence showing that individuals with similar environmental exposures present considerable differences in the risk of pulmonary fibrosis.^9, 10^ Genetic studies conducted have successfully identified genetic risk factors, contributing to IPF risk like telomerase-related genes (*TERT*, *TERC*, *RTEL1*), surfactant-associated genes (*SFTPA*, *SFTPC*), mucin-related genes (*MUC5B, MUC2*), cell adhesion molecules (*DSP, DPP9*), immune response and inflammation-related genes (*TGFB1*, *IL1RN*, *TLR3*, *IL8*, *HLA, TOLLIP*), cell-cycle progression related genes (*CDKN1A*, *TP53*) and many more.^11–17^ Biological pathways implicated in genetic studies include several IPF susceptibility factors, such as telomere maintenance, host defense, cell-cell adhesion, epithelial integrity, and DNA repair. Discovery of these disease-associated genetic variants has shed light on inherited risk factors influencing disease risk.

This study was undertaken given the increasing number of genetic association studies in IPF and the necessity of replicability of genetic variants to identify variants truly associated with IPF risk. Meta-analysis is a useful quantitative approach for determining the true effect of the single nucleotide polymorphism (SNP) with greater statistical power minimizing between-study heterogeneity. Thus we attempted to perform a comprehensive systematic review and meta-analyses and to the best of our knowledge, this is the largest and most comprehensive assessment of currently available literature on genetic association studies in IPF. This study is an attempt to evaluate the genetic basis of IPF in understanding the pathophysiology as well as in determining the role of genetics in disease risk.

## 2. METHODS

This study was performed based on Human Genome Epidemiology Network for systematic review of genetic-association studies and it has been conducted based on the Preferred Reporting Items for Systematic Reviews and Meta-Analyses (PRISMA) guidelines (Supplementary Table S1).^18, 19^ This study has been registered on PROSPERO [registration number: CRD42022297970].

### 2.1. Search for publications

A systematic two-stage literature search was conducted on electronic databases to identify relevant studies (Supplementary Fig. S1). Articles were selected and assessed by two authors (PS and DG) individually. In case of any discrepancy, it was resolved by consensus or the involvement of a third author (RK). The first literature search was done on PubMed and Web of Science to retrieve all articles that discuss the genetic association of IPF until November 30, 2021, using standard medical subject heading (MeSH) terms related to “idiopathic pulmonary fibrosis”, “genetic variants” with AND/OR Boolean operators (Supplementary Table S2). A manual search was done on the bibliography of finally selected articles for additional references. This search yielded 5642 publications, which were screened by title/abstract and later by full text to identify publications that met our inclusion criteria. Detailed inclusion and exclusion criteria are described in Supplementary file S1. The second targeted literature search was performed on PubMed, Web of Science and Scopus databases independently for “gene” OR “genetic variant” with IPF, for the genes/genetic variants identified from the first systematic search which were replicated in two or more studies. These articles were retrieved until December 25, 2021, using the MeSH terms (Supplementary Table S2). Finally, the bibliography of all included articles was screened for additional studies.

### 2.2. Data extraction and quality assessment

Data extraction and quality assessment were done by one author (PS) and checked by a second author (DG). A third investigator (RK) was consulted on every conflicting opinion. Data concerning the demographic and clinical characteristics of the included studies were extracted. Detailed information is provided in Supplementary file S2.

Quality assessment of studies included in meta-analyses was assessed using modified Newcastle-Ottawa scale (NOS) for case-control studies of genetic association.^20^ Quality assessment was done based on 3 criteria – Selection, Comparability and Exposure. Studies scoring a score of ≤3, 4-6 and ≥7 out of 9 were deemed poor, moderate and of high quality, respectively.^21^

### 2.3. Statistical analysis

Statistical analysis and data visualization were performed with STATA16.0 software [StataCorp LP, College Station, TX, USA] and R Studio version 3.4.2 [R Project for Statistical Computing]. The overall pooled odds ratio (OR) and their corresponding 95% confidence interval (CI) were calculated to estimate the strength of association of SNP with IPF susceptibility. Only those SNPs that have been investigated in at least three studies were included in the meta-analysis. For the remaining SNPs that didn’t qualify for a meta-analysis, a pooled OR estimate has been calculated. Meta-analyses were performed for six SNPs [rs35705950 (*MUC5B*), rs2736100 (*TERT*), rs2609255 (*FAM13A*), rs2076295 (*DSP*), rs12610495 (*DPP9*) and rs1800470 (*TGF-β1*)]. The pooled OR was calculated using the fixed-effect and random-effects model according to heterogeneity. Heterogeneity was assessed by Cochran Q statistics and Higgins I^2^ test statistics.^22, 23^ Fixed-effect model (inverse variance) was used when lack of significant heterogeneity was indicated (p > 0.10 and I^2^ < 50%); otherwise, the random-effects model (restricted maximum likelihood) was used.^24^ The significance of the pooled OR was assessed by z-test, and p < 0.05 was considered statistically significant. Only allelic model of OR assessment is reported as for many publications raw data to calculate pooled OR in other models was unavailable. Potential publication bias was assessed by visually inspecting the asymmetry of funnel plots of effect sizes (OR) versus standard errors. Egger’s test was used to assess publication bias statistically.^25^ The significance of the publication bias was assessed by z-test, and p < 0.10 was considered statistically significant. Subgroup analysis was performed to evaluate ethnicity-specific (Europeans and Asians) effects for polymorphisms. Sensitivity analysis was performed by excluding a single study each time to understand the impact of the removal of respective studies and to check the stability of the significant results.

### 2.4. Assessment of cumulative evidence

Venice criteria were applied to all significant associations identified by meta-analysis, to evaluate the epidemiological credibility of each of the associations.^26^ The grading criteria included three categories: the amount of evidence, replication, and protection from bias, which were rated as A, B, or C, detailed description was given in table S3. Based on these ratings, credibility was defined as strong (if all three grades were A), moderate (if all three grades were A or B), or weak (if any grades were C).

## 3. RESULTS

### 3.1. Literature search and study selection

A total of 5642 (PubMed, N = 2687; Web of Science, N = 2955) articles were obtained from our first systematic literature search. 1508 articles were removed when screened for duplicates. On screening the remaining 4134 articles by title and abstract, based on the pre-determined inclusion and exclusion criteria, 3834 and 198 articles were excluded for each title and abstract screening step, respectively. Detailed reasoning for the same is included in Supplementary file S3. Finally, full texts of the remaining 102 articles were screened resulting in the further exclusion of 46 articles. One additional article was added from cross-references of the included studies. Thus, finally, 57 studies investigating the role of genetic variants in IPF susceptibility were included. The flow diagram of this search and study selection strategy is shown in Supplementary Fig. S2. Out of 106 significantly associated genes with IPF risk, ten SNPs [rs35705950 (*MUC5B*), rs2736100 (*TERT*), rs2609255 (*FAM13A*), rs2076295 (*DSP*), rs12610495 (*DPP9*) and rs1800470 (*TGF-β1*), rs7934606 *(MUC2*), rs1278769 *(ATP11A*), rs6793295 *(LRRC34*), rs111521887 *(TOLLIP*)] were replicated in two or more studies and were included for second literature search, independently. Systematic literature search for *MUC5B, TERT, FAM13A, DSP, DPP9, TGF-β1, MUC2, ATP11A, LRRC34* and *TOLLIP* with their respective genetic variants obtained from the first search yielded 3812, 2275, 212, 779, 51, 6214, 2727, 53, 8, 87 articles, respectively. Followed by title, abstract and full-text screening of these articles based on inclusion/exclusion criteria. Articles were excluded if they were grey literature (review, systematic review, meta-analysis), do not report case-control studies in humans, discuss IPF with some other comorbid condition, or unavailability of sufficient data for conducting meta-analysis. Articles from cross-references were also included. Finally, articles having sufficient data for conducting meta-analysis were included. Meta-analyses was performed for six SNPs [rs35705950 (*MUC5B*), rs2736100 (*TERT*), rs2609255 (*FAM13A*), rs2076295 (*DSP*), rs12610495 (*DPP9*) and rs1800470 (*TGF-β1*)] having three or more independent studies. The individual study selection is represented by the PRISMA flow diagram (Fig. 1).

**Fig. 1:**
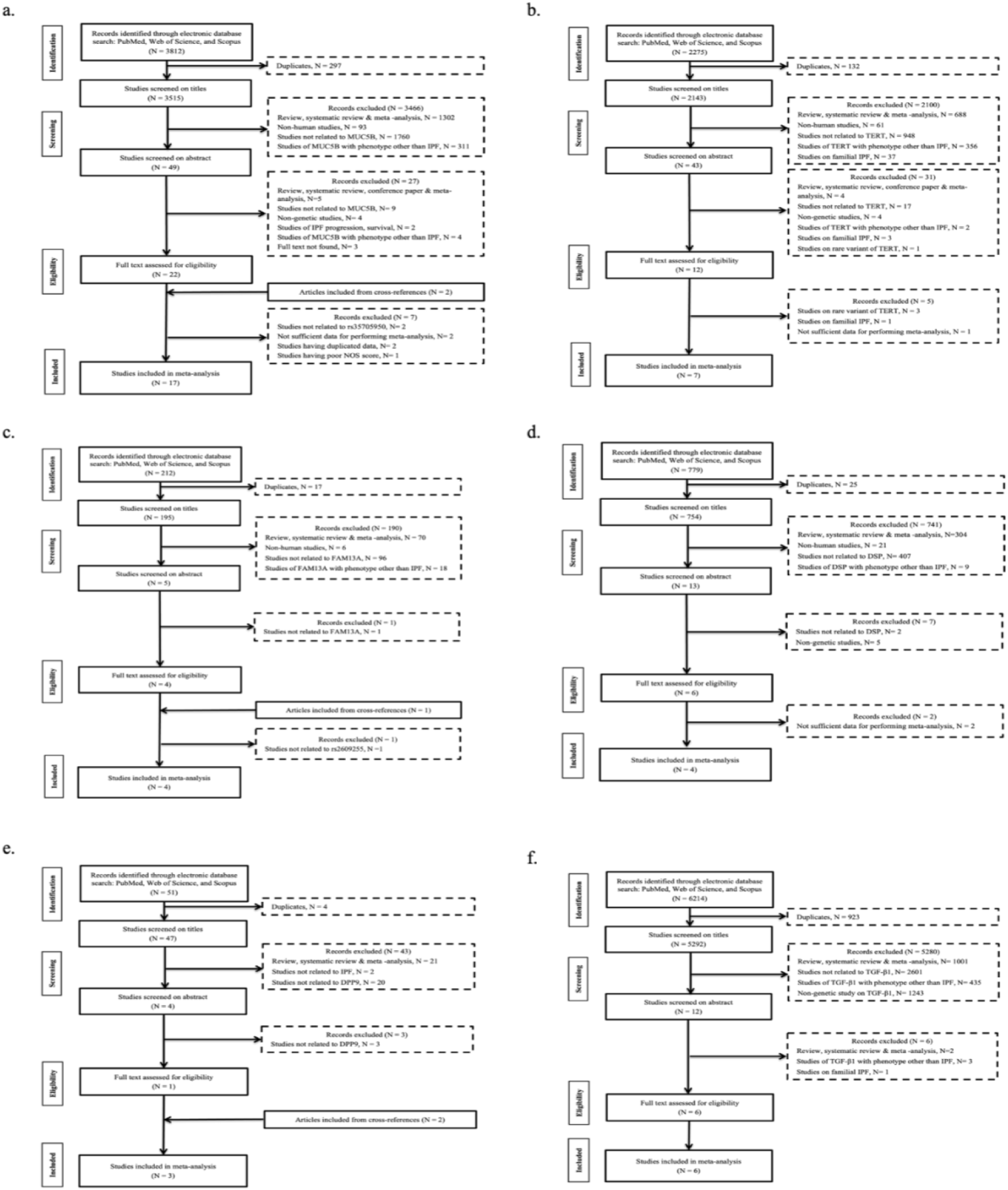
PRISMA flow diagram showing step-wise selection of studies for meta-analysis. Flow diagram for the inclusion and exclusion of studies exploring the role of genetic variants- a) rs35705950 (*MUC5B*), b) rs2736100 (*TERT*), c) rs2609255 (*FAM13A*), d) rs2076295 (*DSP*), e) rs12610495 (*DPP9*), f) rs1800470 (*TGF-β1*), in IPF risk. The number of studies excluded on each step is represented as N.

### 3.2. Characteristics of the studies

All the 57 included studies from the first search strategy adapted a common case-control study design, of which five were genome-wide association studies (GWAS)^11, 12, 15, 16, 27^ and 52 were candidate gene association studies. The characteristics of the included studies and genotypic/allelic distributions of the polymorphisms are shown in Supplementary Table S4. A total of 16483 patients with IPF and 45510 controls were included in our systematic review. The mean age of all the included subjects was 67.12 (range, 54.6-74) years for IPF cases and 55.64 (range, 34.5-71.4) years for controls. The ratio of male to female was 2.16 and 1.49 in cases and controls, respectively. The recruited cohort was of European (45.22%), Asian (44.08%), and American (10.70%) origin. In the included studies, cases were defined as patients diagnosed with sporadic IPF, i.e. no family history of IPF. Except in two studies, one included seven familial IPF cases in a total of 155 cases^28^ and the other included fibrotic idiopathic interstitial pneumonias as cases.^12^ The IPF patients were diagnosed based on American Thoracic Society/ European Respiratory Society guidelines (2000, 2001, 2002, 2011, 2013, 2018).^29–34^ Except in two studies, in the first study, the diagnostic criteria was BTS/Thoracic society of Australia/New Zealand and Irish Thoracic Society ILD guideline 2008^35, 36^ and in the second study, IPF patients were evaluated with a medical history, physical examination, chest radiograph, pulmonary function tests, and lung biopsy.^37^ On elaborate systematic analysis of published literature, 291 polymorphisms were found to be significantly associated with IPF susceptibility.

### 3.3. Quality assessment

Quality assessment scores were obtained by modified NOS (Supplementary Table S5) and only those studies which had high and moderate-quality were included in the meta-analysis. All the included studies in this meta-analysis were deemed to be of good quality except one study^38^ which is of poor quality and thus was excluded from the meta-analysis.

### 3.4. Meta-analyses results

Meta-analyses were done for six SNPs in six candidate genes (*MUC5B, TERT, FAM13A, DSP, DPP9, TGF-β1*). The average sample size analysed per polymorphism was 13308 (range, 1564-24907) combining cases and controls and were combined from 6.8 studies (range, 3-17). The most frequently analysed genes were *MUC5B*,^11, 12, 15, 39–52^ *TERT*, ^12, 16, 28, 41, 43, 49, 53^ and *TGF-β1*.^40, 54–58^

#### 3.4.1. rs35705950 (MUC5B) and IPF susceptibility

Seventeen studies^11, 12, 15, 39–52^ investigating the association of rs35705950 (*MUC5B*) in IPF risk were included (Fig. 1a, Table 1). Out of these seventeen studies, thirteen^11, 12, 15, 39, 41–44, 48–52^ included individuals of European origin, three^40, 46, 47^ of Asian population and one study^45^ was conducted in both European and Asian. Included IPF patients (N = 7101) and controls (N = 17806) had a mean age of 62.05 (range, 54.6-70) and 55.23 (range, 34.5-67.7) years, respectively. On performing meta-analysis based on random-effects model [I^2^ = 81.85%; p < 0.1], rs35705950 revealed strong association with risk of IPF [T vs C; OR = 3.85; 95% CI = 3.24-4.47; p < 0.05] (Table 2, Fig. 2a). Subgroup analysis by ethnicity indicated significant results among Europeans [OR = 3.97; 95% CI = 3.52-4.60; p < 0.05; I^2^ = 81.93%; p < 0.1 for heterogeneity]. Whereas among Asians, pooled OR for risk allele was lower as compared to overall pooled OR [OR = 2.54; 95% CI = 0.78-4.31; p < 0.05; I^2^ = 17.97%; p > 0.1 for heterogeneity] (Supplementary Table S6). Thus, Asian carriers of the risk allele (T) are at lower risk of developing IPF than Europeans.

**Fig. 2:**
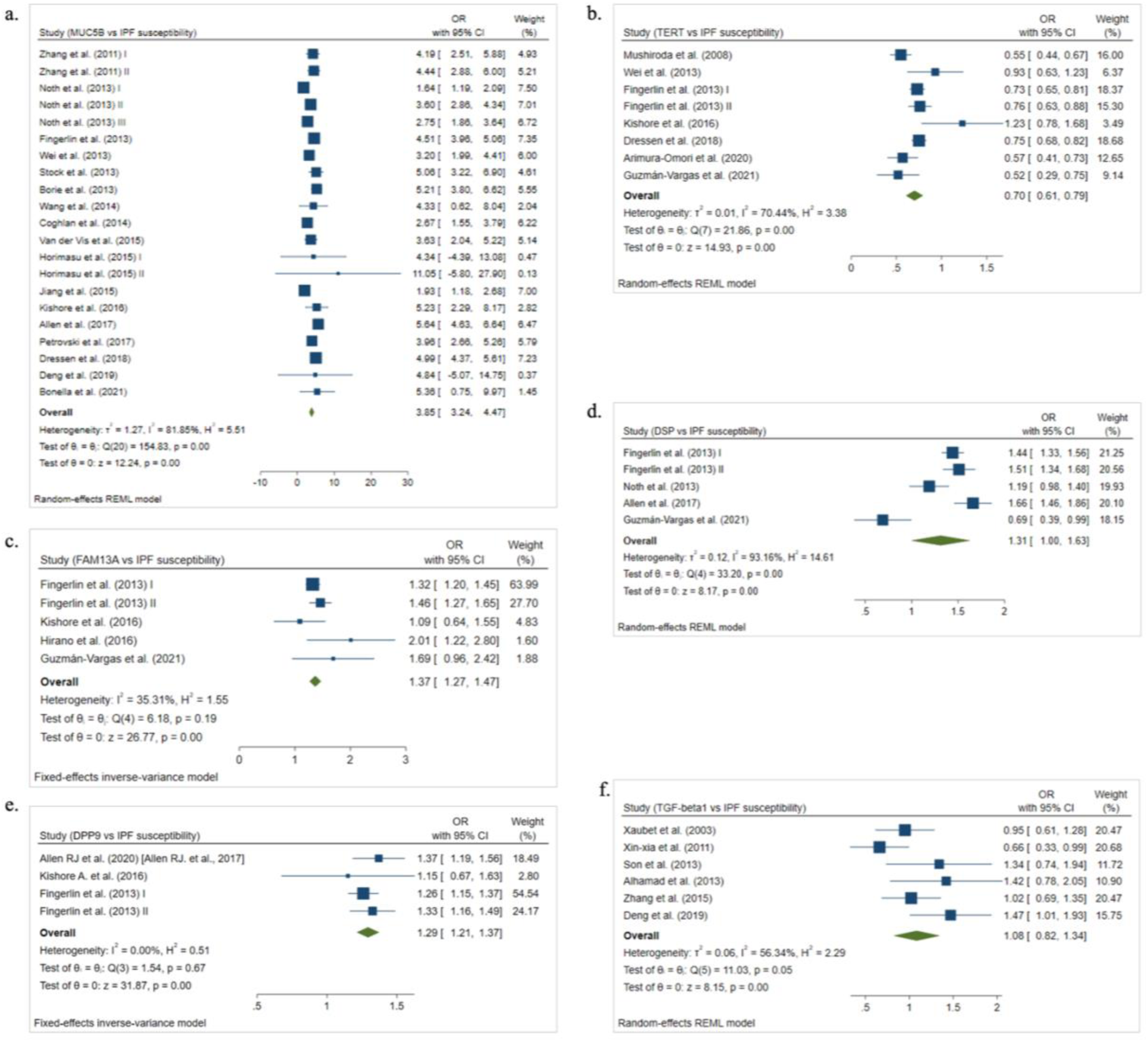
Forest plots showing OR with 95 % confidence intervals (CI) from individual studies and pooled data, for association between a) T allele of rs35705950 (*MUC5B*); b) C allele of rs2736100 (*TERT*); c) G allele of rs2609255 (*FAM13A*); d) G allele of rs2076295 (*DSP*); e) G allele of rs12610495 (*DPP9*); f) T allele of rs1800470 (*TGF-β1*), in the overall population. The square and horizontal lines correspond to the study-specific OR and 95% confidence interval (CI). The area of the square refers to the study specific weight (inverse of variance). The diamond represents the summary of OR and 95% CI.

**Table 1:**
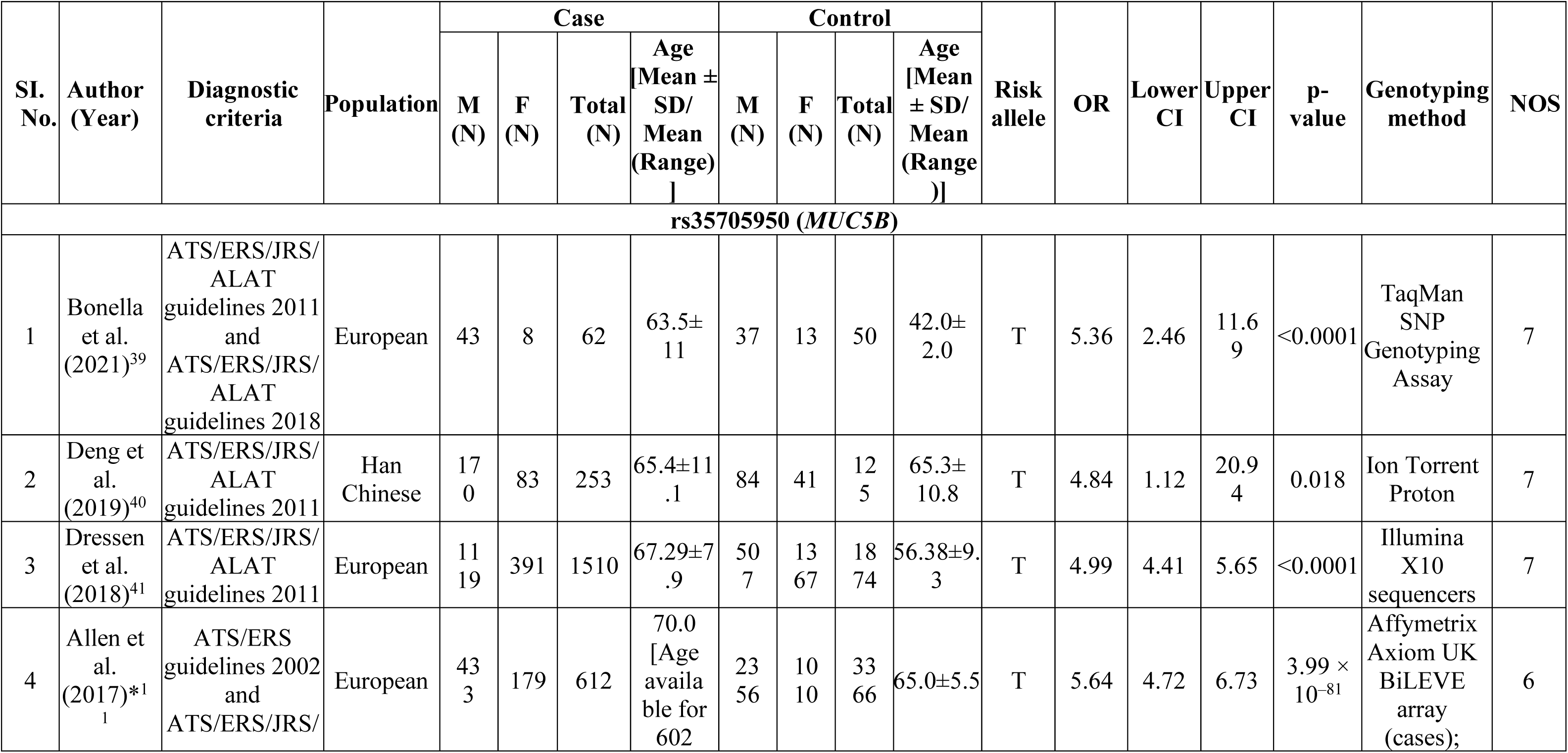
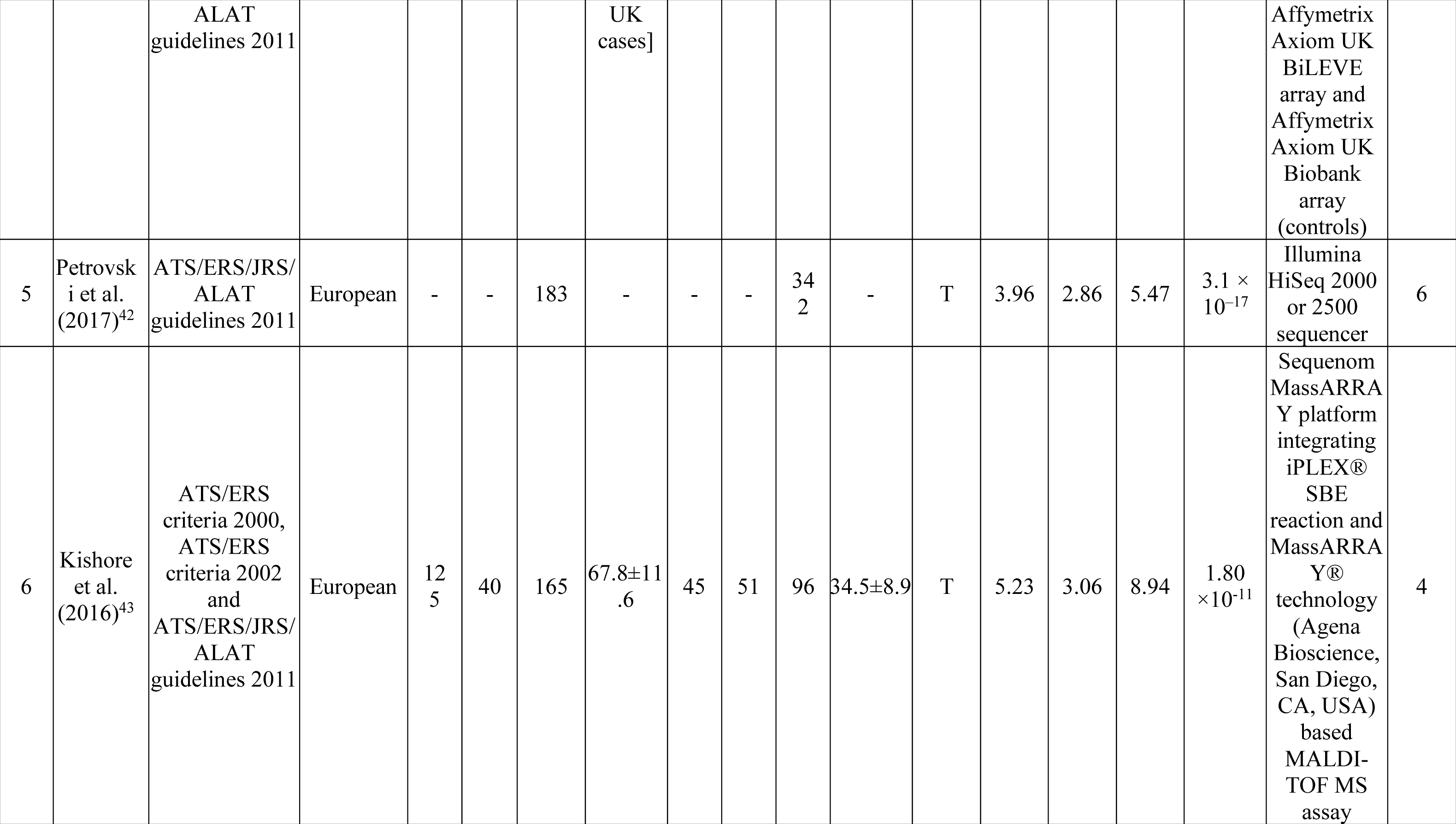
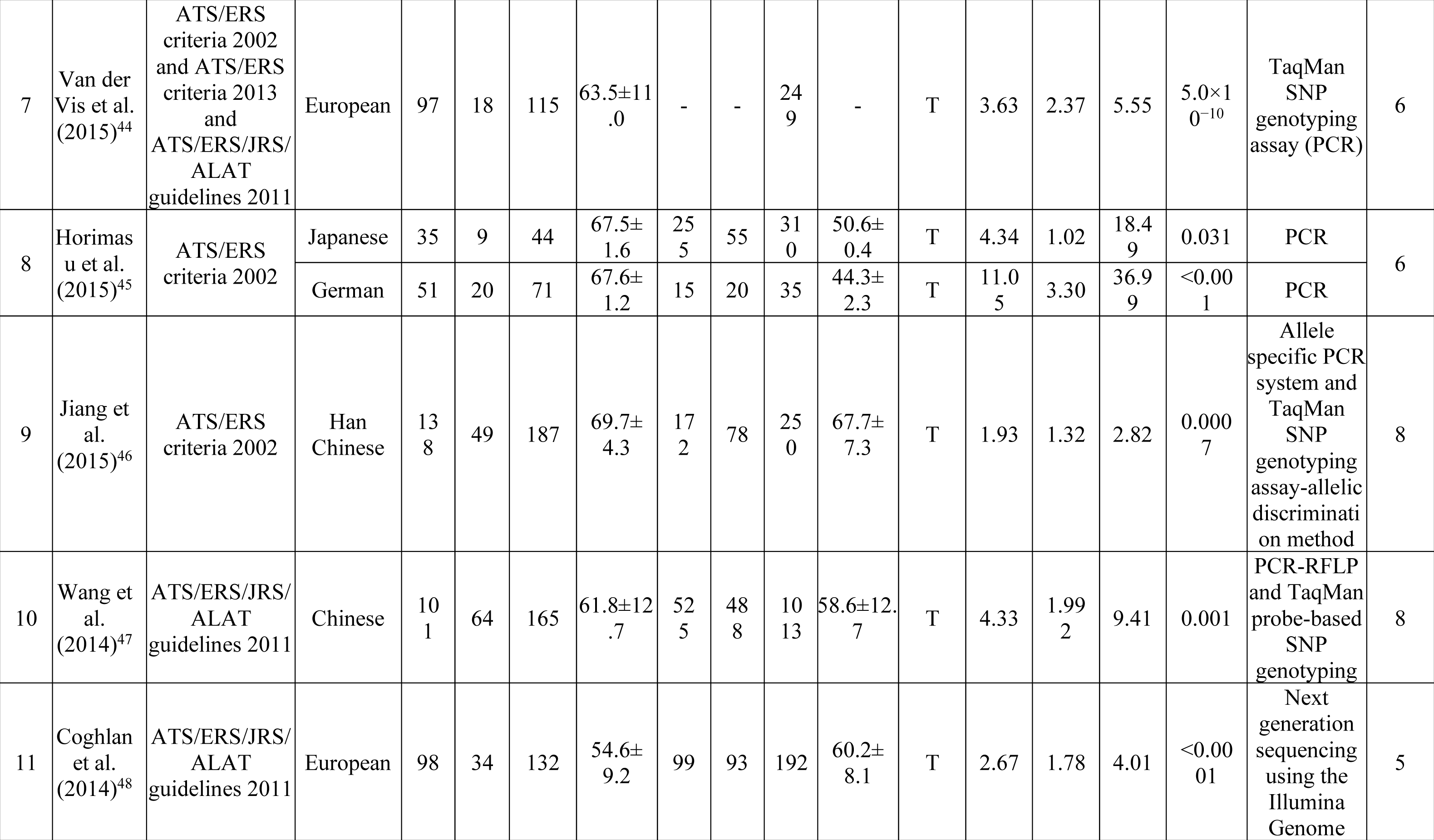
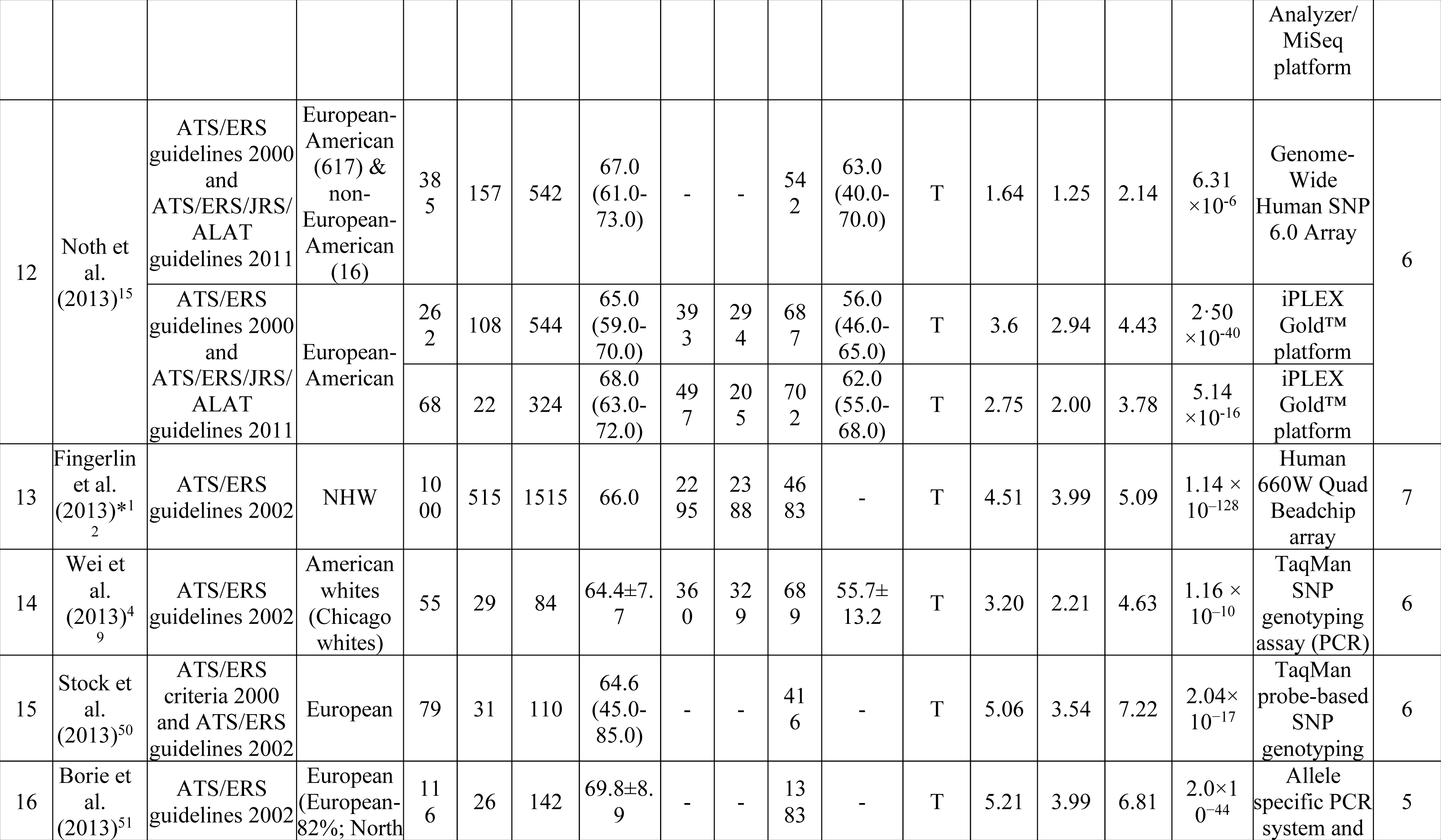
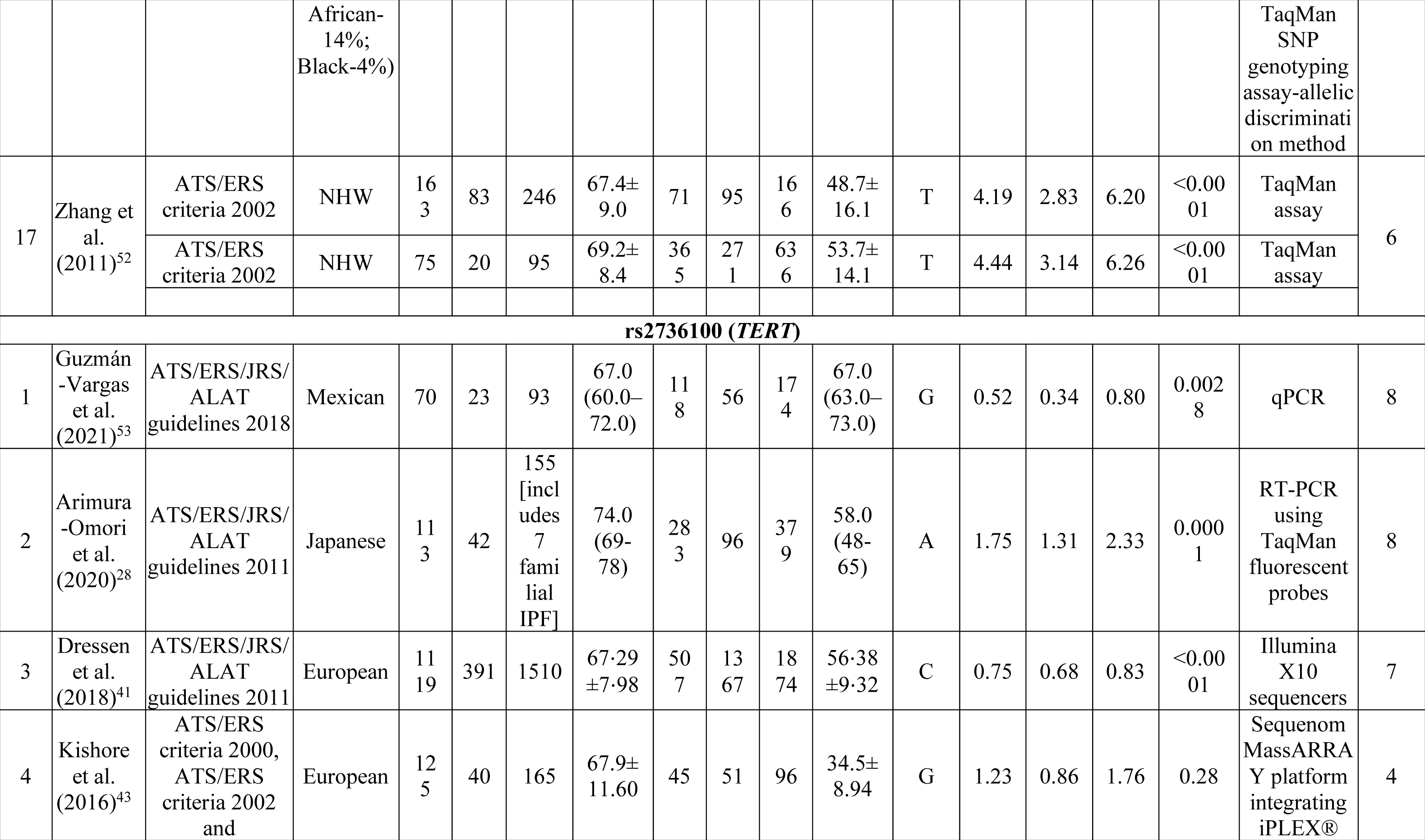
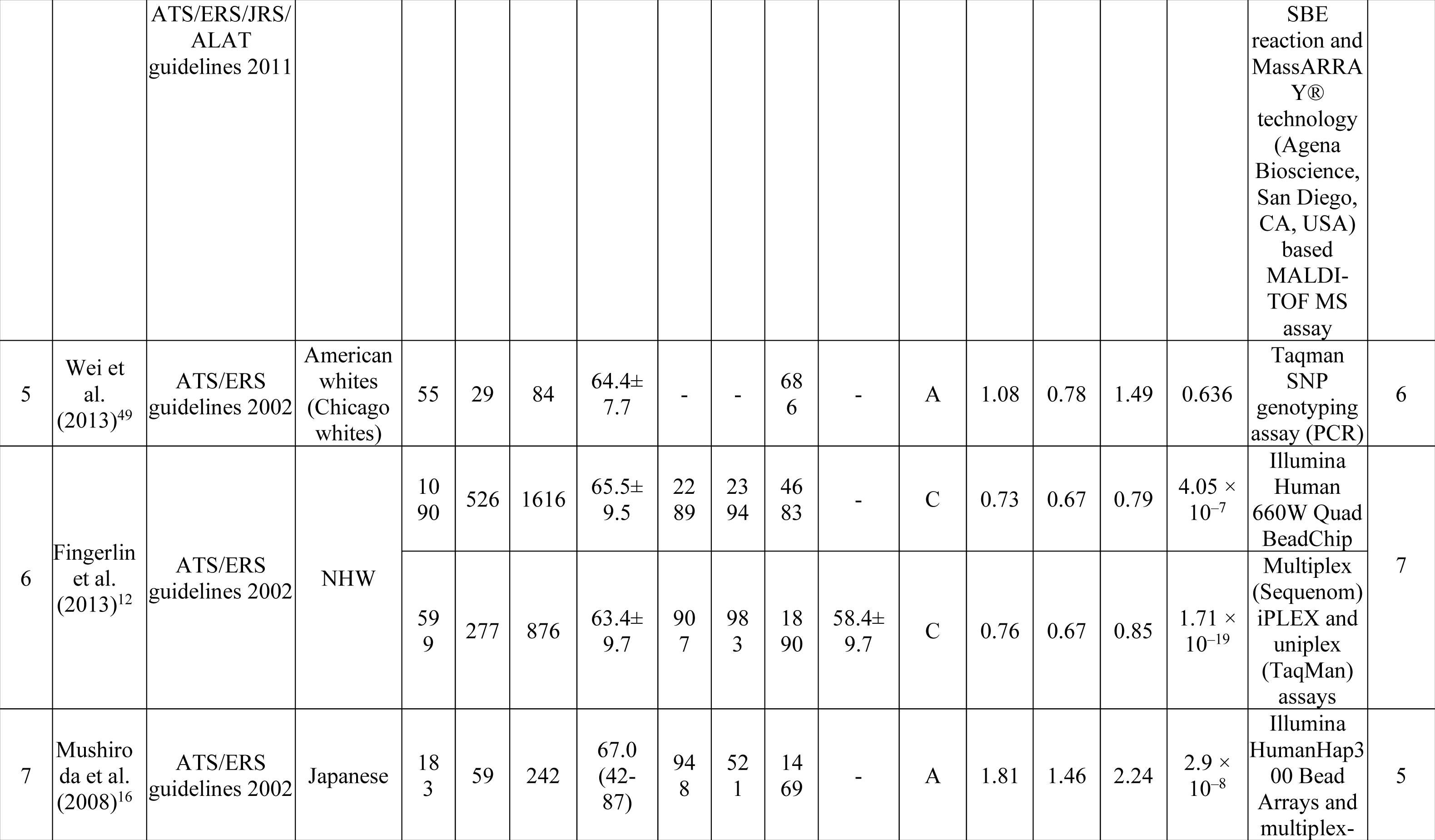
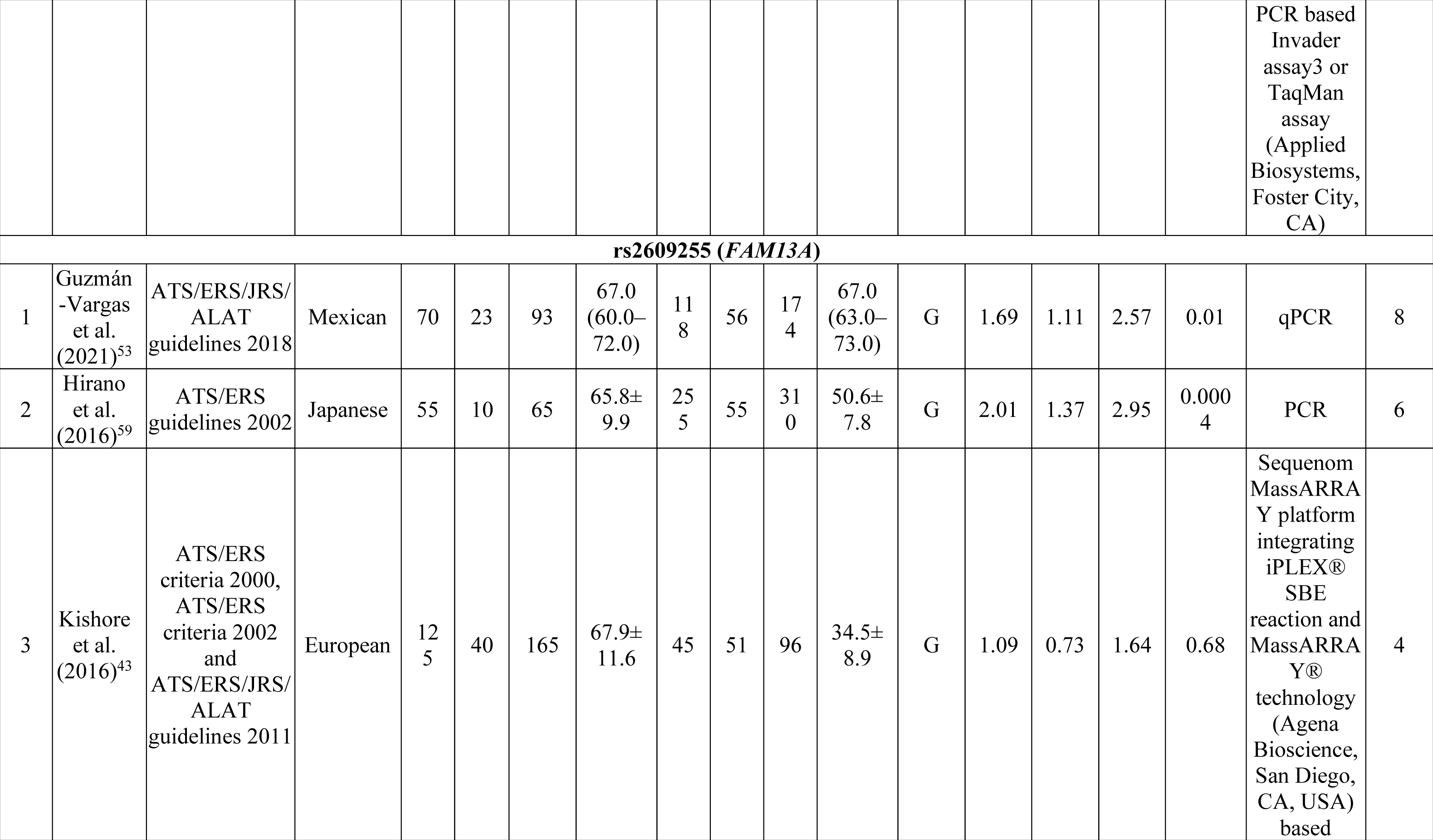
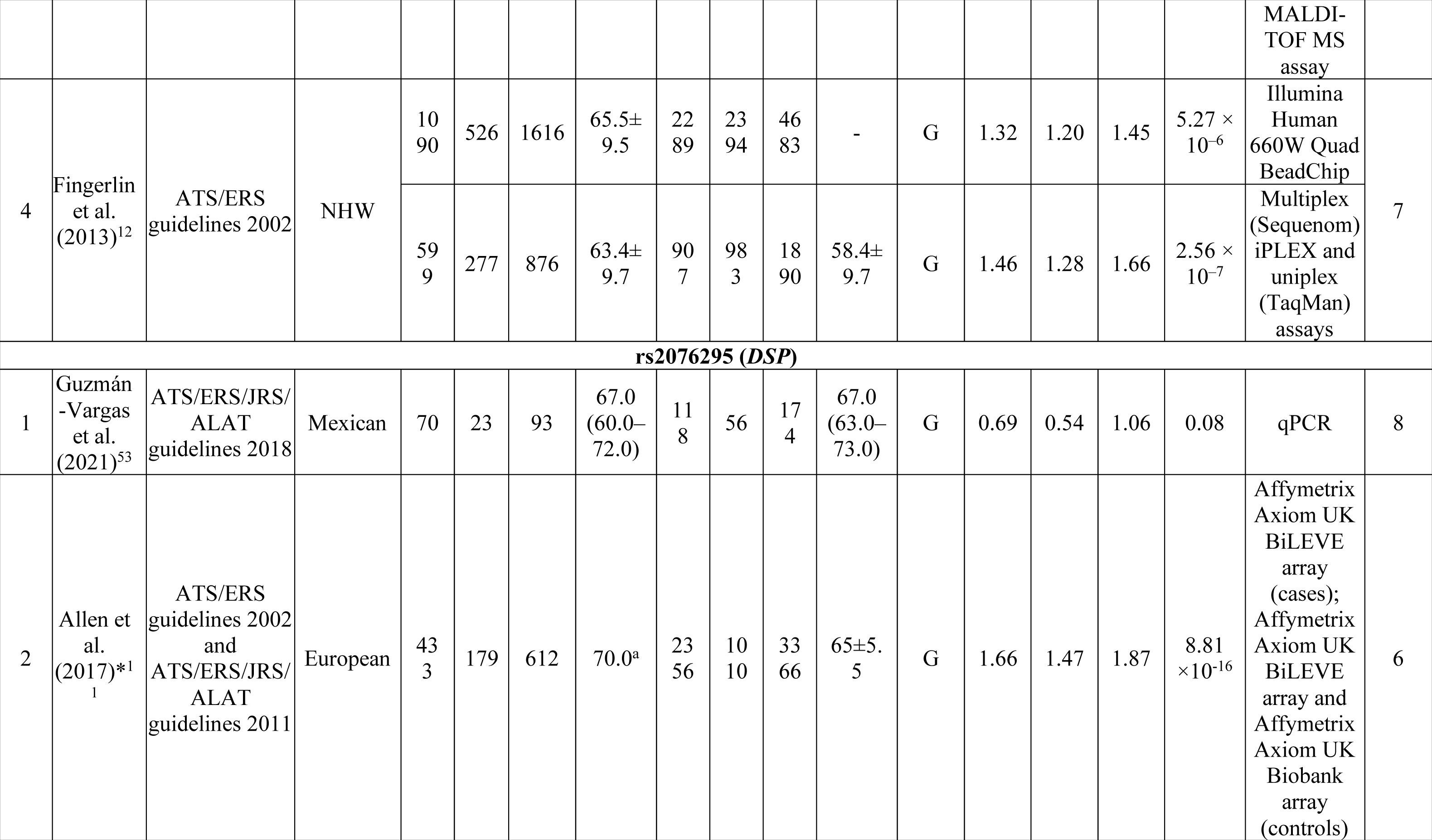
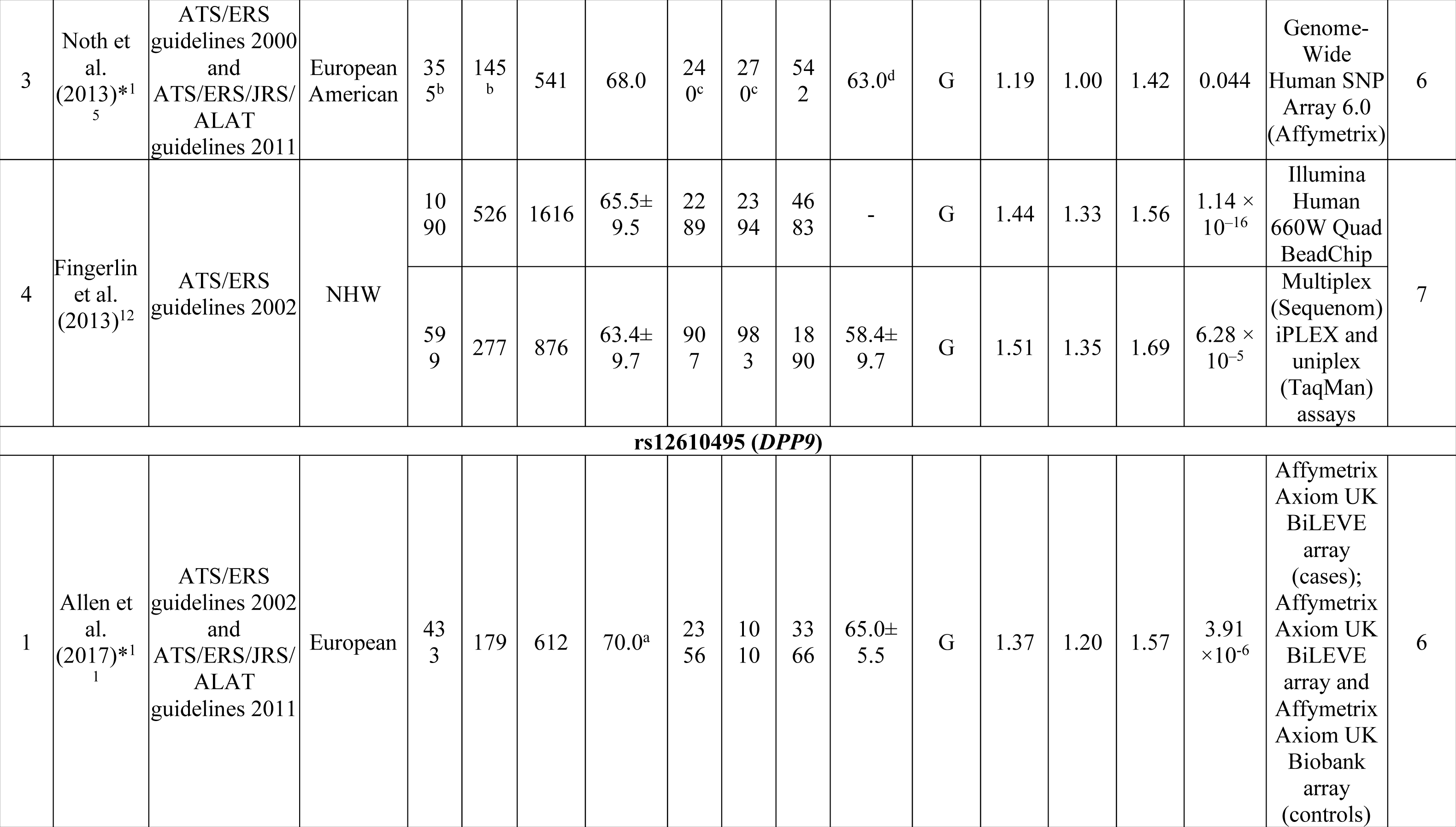
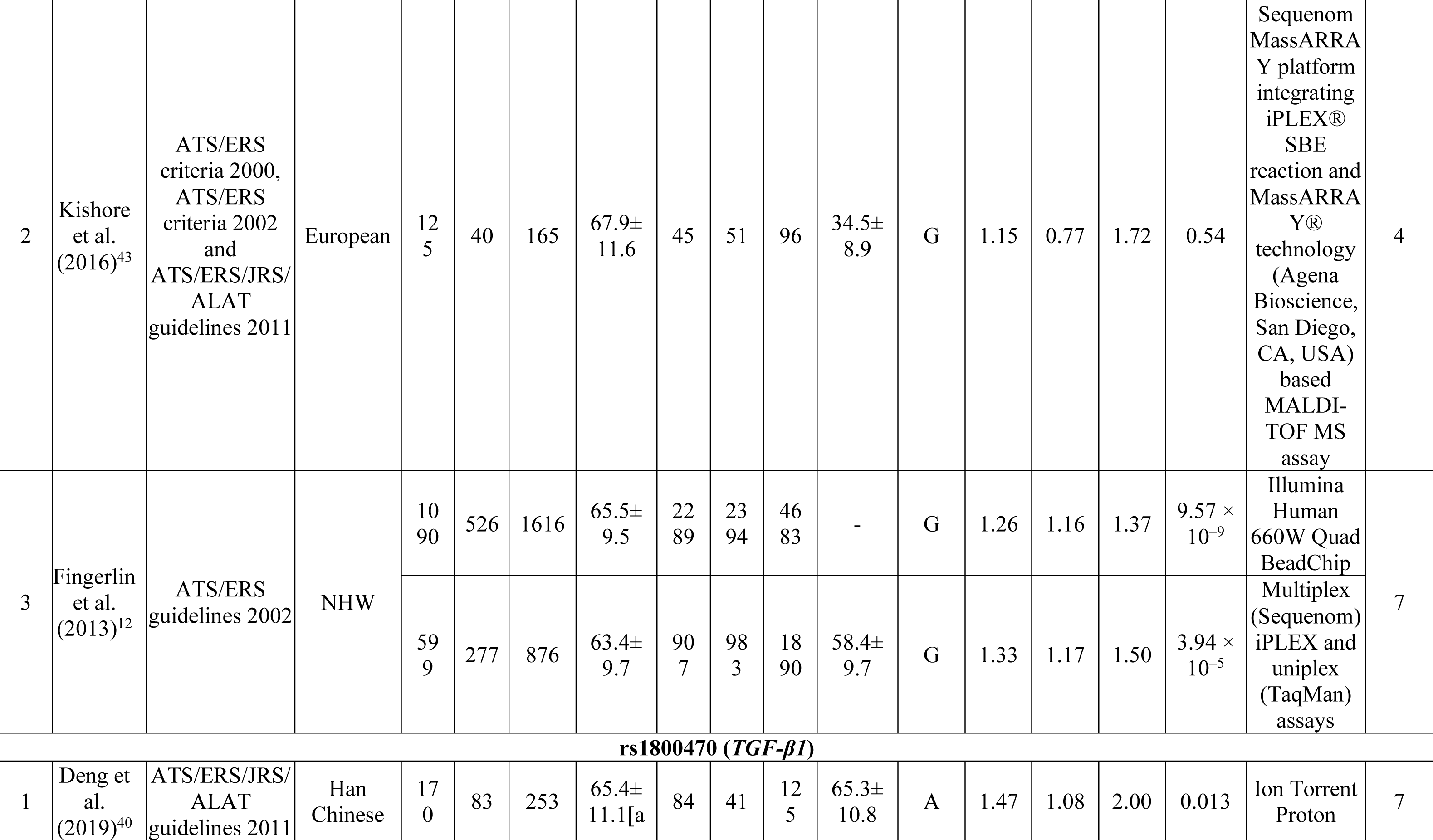
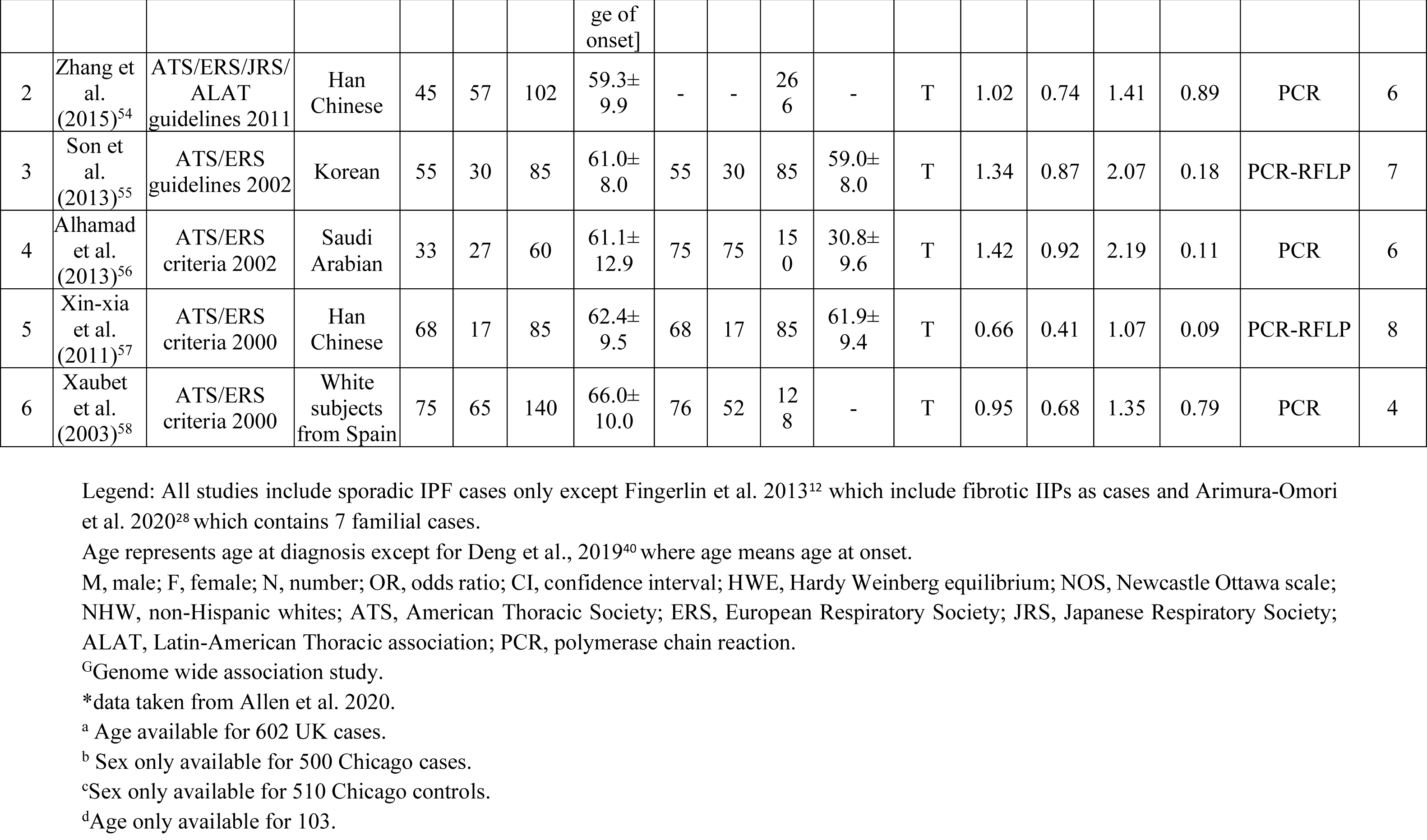
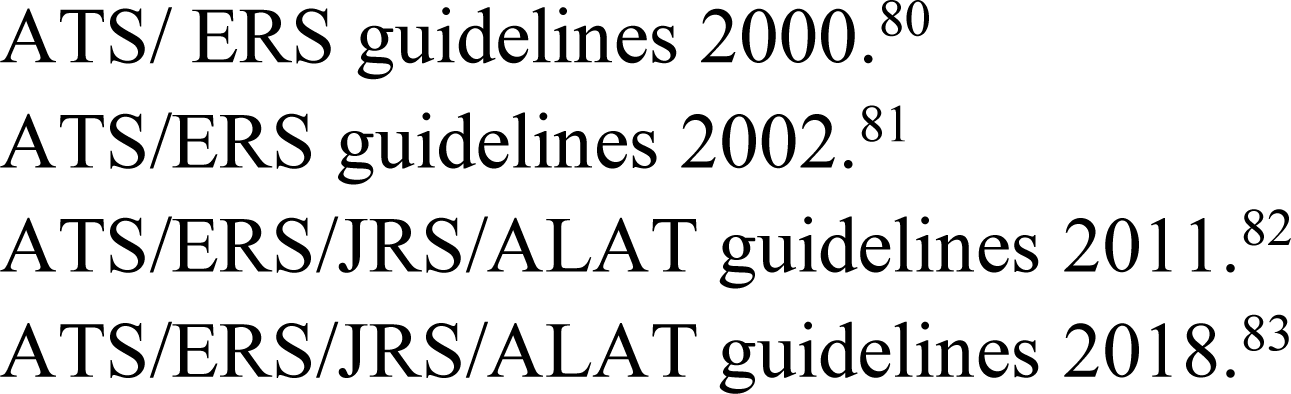
Characteristics of included studies for assessment of association between genetic variants- a) rs35705950 (*MUC5B*), b) rs2736100 (*TERT*), c) rs2609255 (*FAM13A*), d) rs2076295 (*DSP*), e) rs12610495 (*DPP9*), f) rs1800470 (*TGF-β1*) and IPF susceptibility.

**Table 2:** Pooled odds ratios (OR) for studies exploring association of genetic variants- rs35705950 (*MUC5B*), rs2736100 (*TERT*), rs2609255 (*FAM13A*), rs2076295 (*DSP*), rs12610495*9* (*DPP9*), rs1800470 (*TGF-β1*), rs7934606 (*MUC2*), rs1278769 (*ATP11A*), rs6793295 (*LRRC34*), rs111521887 (*TOLLIP*) with IPF susceptibility.

| Sl. No. | Gene | Variant | Risk allele | Number of studies (N) | Ethnicity | Number of participants |  |  | IPF risk |  | Heterogeneity |  | Model | Test for publication bias |  | Venice criteria grade | Cumulative evidence of associations |
| --- | --- | --- | --- | --- | --- | --- | --- | --- | --- | --- | --- | --- | --- | --- | --- | --- | --- |
|  |  |  |  |  |  | Cases (N) | Controls (N) | Total (N) | OR (95% CI) | p-value | I<sup>2</sup>(%) | p-value |  | Egger's test |  |  |  |
|  |  |  |  |  |  |  |  |  |  |  |  |  |  | Z-score | p-value |  |  |
| 1 | *MUC5B* | rs35705950 | T | 17<sup>11,12,15,39–52</sup> | European and Asian | 7101 | 17806 | 24907 | 3.85 (3.24-4.47) | **0.00** | 81.85 | 0.00 | R | 1.51 | 0.13 | AAA | +++ |
| 2 | *TERT* | rs2736100 | C | 7<sup>12,16,28,41,43,49,53</sup> | European and Asian | 4741 | 10968 | 15709 | 0.70 (0.61-0.79) | **0.00** | 70.44 | 0.00 | R | 1.48 | 0.14 | ABA | ++ |
| 3 | *FAM13A* | rs2609255 | G | 4<sup>12,43,53,59</sup> | European and Asian | 2815 | 7153 | 9968 | 1.37 (1.27-1.47) | **0.00** | 35.31 | 0.19 | F | 1.17 | 0.24 | ABA | ++ |
| 4 | *DSP* | rs2076295 | G | 4<sup>11,12,15,53</sup> | European | 3738 | 10655 | 14393 | 1.31 (1.00-1.63) | **0.00** | 93.16 | 0.00 | R | -2.04 | 0.04 | ABC | + |
| 5 | *DPP9* | rs12610495 | G | 3<sup>11,12,43</sup> | European | 3269 | 10035 | 13304 | 1.29 (1.21-1.37) | **0.00** | 0.00 | 0.66 | F | 0.02 | 0.98 | ABA | ++ |
| 6 | *TGF-β1* | rs1800470 | T | 6<sup>40,54–58</sup> | European, Asian and | 725 | 839 | 1564 | 1.08 (0.82-1.34) | **0.00** | 56.34 | 0.05 | R | 2.57 | 0.01 | BBC | + |
|  |  |  |  |  | Saudi Arabia |  |  |  |  |  |  |  |  |  |  |  |  |
| 7 | *MUC2* | rs7934606 | T | 2<sup>12,43</sup> | European | 2657 | 6669 | 9326 | 1.53<br>(1.43-1.63) | **0.00** | 21.55 | 0.27 | F | 1.60 | 0.11 | - | - |
| 8 | *ATP11A* | rs1278769 | A | 2<sup>12,43</sup> | European | 2657 | 6669 | 9326 | 0.79<br>(0.73-0.85) | **0.00** | 0.00 | 0.98 | F | -0.19 | 0.85 | - | - |
| 9 | *LRRC34* | rs6793295 | C | 2<sup>12,43</sup> | European | 2657 | 6669 | 9326 | 1.34<br>(1.24-1.43) | **0.00** | 39.01 | 0.19 | F | -1.34 | 0.18 | - | - |
| 10 | *TOLLIP* | rs111521887 | G | 2<sup>15,53</sup> | European | 1503 | 2105 | 3608 | 1.27<br>(0.89-1.65) | **0.00** | 84.92 | 0.00 | R | 0.78 | 0.43 | - | - |

Legend: Bold characters highlight significantly associated alleles with respective p-values.
OR, Odds ratio; CI, Confidence interval; R, Random effect model (maximum likelihood method); F, Fixed effect model (inverse-variance method); N, Number.
All calculations were done using Stata/MP 16.0.

#### 3.4.2. rs2736100 (TERT) and IPF susceptibility

Seven case-control studies^12, 16, 28, 41, 43, 49, 53^ (4741 cases and 10968 controls) investigating association between rs2736100 (*TERT*) and IPF susceptibility were included (Fig. 1b, Table 1). IPF cases with a mean age of 67.06 (range, 63.4-74) years and controls with a mean age of 54.86 (range, 34.5-67) years were included in the analysis. Out of these seven included studies, five studies^12, 41, 43, 49, 53^ investigated European subjects and two^16, 28^ were performed on Asian subjects. For calculating pooled OR, random-effects model was used as significant heterogeneity was observed [I^2^ = 70.44%; p < 0.1] among the studies (Table 2). rs2736100 revealed a strong association with IPF risk [C vs A; OR = 0.70; 95% CI = 0.61-0.79; p < 0.05] (Table 2, Fig. 2b). In the subgroup analysis by ethnicity, among Asians C allele is more protective [OR = 0.56; 95% CI = 0.46-0.65; p < 0.05; I^2^ = 0.00%; p > 0.1 for heterogeneity] as compared to Europeans [OR = 0.74; 95% CI = 0.70-0.79; p < 0.05; I^2^ = 0.00%; p > 0.1 for heterogeneity] (Supplementary Table S6). Thus, Asian carriers of the C allele may have increased protection against IPF.

#### 3.4.3. rs2609255 (FAM13A) and IPF susceptibility

Four studies^12, 43, 53, 59^ on the relationship between rs2609255 (*FAM13A*) and IPF susceptibility were included in the meta-analysis (Fig. 1c, Table 1). 2815 IPF cases with a mean age of 65.92 (range, 63.4-67.9) and 7153 controls with a mean age of 52.63 (range, 34.5-67) were included. Out of these four studies, three^12, 43, 53^ included patients of European origin and one^59^ of Asian population. Meta-analysis based on fixed-effect model [I^2^ = 35.31%; p > 0.1] revealed a significant association between rs2609255 and IPF susceptibility [G vs T; OR = 1.37; 95% CI = 1.27-1.47; p < 0.05] (Table 2, Fig. 2c). Stratification by ethnicity indicated significant associations among both, Europeans [OR = 1.36; 95% CI = 1.27-1.47; p < 0.05; I^2^ = 16.35%; p > 0.1 for heterogeneity] and Asians [OR = 2.01; 95% CI = 1.22-2.80; p < 0.05] (Supplementary Table S6).

#### 3.4.4. rs2076295 (DSP) and IPF susceptibility

Four studies^11, 12, 15, 53^ involving 3738 IPF cases with a mean age of 66.78 (range, 63.4-70) and 10655 controls with a mean age of 63.35 (range, 58.4-67) were included in the analysis (Fig. 1d, Table 1). All four studies included IPF patients and controls of European origin. The random-effects model was used for calculating pooled OR [G vs T; OR = 1.31; 95% CI = 1.00-1.63; p < 0.05] as significant heterogeneity was observed [I^2^ = 93.16%; p < 0.1] among the studies (Table 2, Fig. 2d).

#### 3.4.5. rs12610495 (DPP9) and IPF susceptibility

Three studies^11, 12, 43^ (3269 cases and 10035 controls) were included in the meta-analysis (Fig. 1e, Table 1). IPF cases and controls included in the analysis were Europeans and had a mean age of 66.72 (range, 63.4-70) and 52.62 (range, 34.4-65) years, respectively. Fixed-effect model was used for calculating pooled estimate as non-significant heterogeneity [I^2^ = 0.00%; p > 0.1] was present among studies (Table 2). Significant associations between rs12610495 and IPF risk were observed [G vs A; OR = 1.29; 95% CI = 1.21-1.37; p < 0.05] (Table 2, Fig. 2e).

#### 3.4.6. rs1800470 (TGF-β1) and IPF susceptibility

Six studies^40, 54–58^ involving 725 IPF cases with a mean age of 62.54 (range, 59.3-66) and 839 controls with a mean age of 54.25 (range, 30.8-65.3) were included in the analysis (Fig. 1f, Table 1). On performing meta-analysis on six included studies based on random-effects model [I^2^ = 56.34%; p < 0.1], rs1800470 revealed strong association with risk of IPF [T vs C; OR = 1.07; 95% CI = 0.84-1.31; p < 0.05] (Table 2, Fig. 2f). In the subgroup analysis pooled OR for risk allele did not change significantly from overall pooled estimate in both, Asians [OR = 1.07; 95% CI = 0.76-1.39; p < 0.05; I^2^ = 57.32%; p < 0.1 for heterogeneity] and Europeans [OR = 0.95; 95% CI = 0.62-1.29; p < 0.05] was observed (Supplementary Table S6).

### 3.5. Sensitivity analysis

Sensitivity analyses were done for six variants which are significantly associated with IPF risk (Supplementary Table S7). The sequential exclusion of each study from the sensitivity analysis did not significantly alter the pooled estimates for any of the variants.

### 3.6. Publication bias

Publication bias was assessed statistically by Egger’s test or visually by inspecting Begg’s funnel plot. Egger’s test indicated significant publication bias (p < 0.10) for two variants (rs2076295 and rs1800470) (Table 2). Supplementary Fig. S3(a-f), shows the funnel plot for all six variants under the allelic model.

### 3.7. Cumulative epidemiological evidence

Out of six SNPs having a significant association with IPF risk in meta-analyses, one variant (rs35705950) received A grades for all three criteria and can therefore be regarded as having strong cumulative epidemiological evidence of an association with IPF risk. Three variants (rs2736100, rs12610495 and rs2609255) received a grade of either A or B in all three criteria and were thus scored as having moderate evidence of an association. And two variants (rs2076295 and rs1800470) received a grade of C in protection from bias and were thus scored as having weak evidence of an association (Table 2).

## 4. DISCUSSION

This systematic meta-analysis is the most comprehensive assessment of available literature on the genetic architecture of IPF susceptibility conducted till date. Here we have performed an extensive two-stage literature search of genetic variants with IPF risk. In stage one search we performed a systematic review identifying 57 articles reporting association of 291 polymorphisms with IPF susceptibility. Subsequently retaining the genetic variants, obtained from stage one and performing a targeted search in stage two for each of six SNPs with IPF susceptibility. Meta-analyses were performed for six SNPs located in six different genes [rs35705950 (*MUC5B*), rs2736100 (*TERT*), rs2609255 (*FAM13A*), rs2076295 (*DSP*), rs12610495 (*DPP9*) and rs1800470 (*TGF-β1*)] showing significant association with IPF. Four variants (rs35705950, rs2736100, rs12610495 and rs2609255) showed moderate to strong cumulative epidemiological evidence for true association with IPF risk. Whereas, the remaining two variants (rs2076295 and rs1800470) provided weak evidence of association. To the best of our knowledge till now out of these six SNPs, the previous meta-analyses had been published only for *MUC5B*.^60, 61^ Also our meta-analysis for *MUC5B* involves about 2.5 times as much data as in previous meta-analyses. Thus, this systematic meta-analysis provides the most exhaustive and systematic assessment so far of the relevance of these six SNPs to IPF susceptibility and our results may provide a holistic view of the genetic architecture of IPF predisposition.

The most notable findings to emerge from this meta-analyses revealed genes involved in alveolar epithelial injuries (*MUC5B, TERT, FAM13A, DSP, DPP9*) and epithelial-mesenchymal transition (EMT) (*TGF-β1*) to play a role in IPF development (Fig. 3). This meta-analysis revealed the strongest association of *MUC5B* promoter variant rs35705950 (T) with IPF risk [OR = 3.85]. *MUC5B* gene encodes mucin 5B, a respiratory tract mucin glycoproteins playing role in mucociliary clearance (MCC) and host defense.^62^ In IPF lesions *MUC5B* was highly expressed and rs35705950 (T) is known to induce *MUC5B* expression in lung tissue of IPF patients. Studies in mice demonstrated that mucin overproduction leads to the accumulation of thick mucus layer thereby affecting MCC and mucosal defense.^63^ This thick mucus layer entraps inhaled substances and inflammatory particles which can cause repetitive micro-injury and increased inflammation in the distal airway and bronchoalveolar junction. Such changes embark abnormal activation of alveolar epithelial cells (AEC), followed by the development of pro-fibrotic phenotypes and finally fibrosis.^63^ Second, *MUC5B* hypersecretion might elevate endoplasmic reticulum stress signalling in the distal lung, which may ultimately cause apoptosis, chronic inflammation and fibrosis.^64, 65^ Thus, dysregulated *MUC5B* expression might contribute to IPF development. In line with previous findings,^60, 61^ our meta-analysis had also shown that when studies were stratified by ethnicity, the strength of association between rs35705950 and IPF risk in Europeans was stronger than that in Asians.

**Fig. 3:**
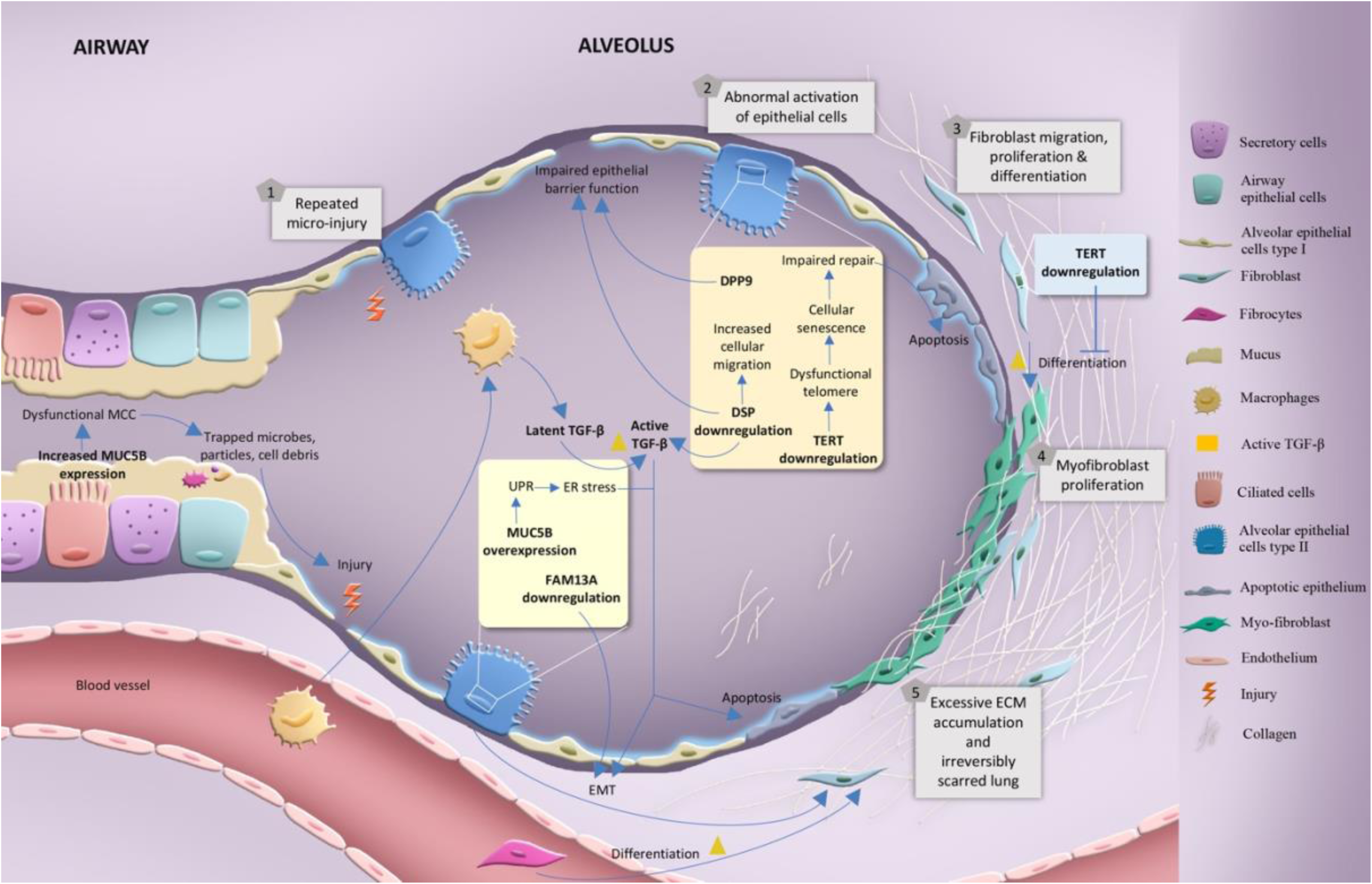
Proposed patho-genetic mechanism of IPF highlighting the role of the genes (or proteins) in the disease pathogenesis. Repetitive micro-injury of epithelial cells in genetically susceptible individuals causes abnormal activation of alveolar epithelial cells (AEC), releasing inflammatory mediators. This leads to increased migration of inflammatory cells in the lungs and enhanced production of pro-fibrotic mediators. This induces fibroblasts migration, proliferation and differentiation into myofibroblasts, resulting in aberrant extracellular matrix (ECM) deposition, lung tissue remodelling and scarring. ***MUC5B***: Mucus accumulation due to the overproduction of mucins (*MUC5B*) will result in altered mucociliary clearance (MCC) and entrapment of inhaled particles. Micro-injury caused by these trapped irritants may cause abnormal activation of AEC. Additionally, overexpression of *MUC5B* in AEC can activate unfolded protein response, elevating endoplasmic reticulum (ER) stress. These can lead to AEC apoptosis, chronic inflammation and fibrosis. ***DSP***: Loss of *DSP*, a cell adhesion molecule, can lead to altered AEC integrity and epithelial barrier dysfunction. Its loss can cause increased production of pro-fibrotic mediators (*TGF-β1*), altered epithelial-mesenchymal transition (EMT), increased cellular migration and fibrotic gene expression. ***DPP9***: altered expression of DPP9 can lead to impaired epithelial barrier function as it is involved in cell adhesion ***TERT***: Deficiency of *TERT* can lead to short/dysfunctional telomere, which results in exacerbation of fibrosis by impairing the repair process and enhanced apoptosis specifically in AEC. ***FAM13A***: Loss of *FAM13A* can accelerate the EMT process through activating the β-catenin pathway. ***TGF-β***: a profibrotic mediator, can induce EMT and increased ECM production. Also, it can promote pulmonary fibrosis through AEC apoptosis and by protecting myofibroblasts against apoptosis. *MUC5B*, mucin5b; *FAM13A*, family with sequence similarity member 13A; *TGF-β*, transforming growth factor-beta; *DSP*, desmoplakin; *DPP9*, dipeptidyl peptidase 9; *TERT*, telomerase reverse transcriptase; MCC, mucociliary clearance; EMT, epithelial-mesenchymal transition; ER, endoplasmic reticulum; UPR, unfolded protein response; ECM, extracellular matrix.

Telomerase gene polymorphisms have been reported to be associated with IPF in recent studies. Our finding shows a negative association of the “C” allele of *TERT* intronic polymorphism (rs2736100) (OR = 0.70) with IPF risk and is found to be more protective in Asians than Europeans. Studies have shown that IPF patients have short telomere versus healthy individuals^66^ and rs2736100 (C) is associated with longer telomere length.^67, 68^ Deficiency of *TERT* which encodes telomerase reverse transcriptase, can lead to short telomere which results in fibrosis exacerbation by impairing the repair process and enhanced apoptosis specifically in AEC.^69, 70^ Whereas it is shown that fibroblast cells of both IPF patients and mice model of bleomycin-induced pulmonary fibrosis (PF) have enhanced telomerase activity which can promote fibrosis by enhancing fibroblast proliferation and resisting apoptosis of fibroblast and differentiated myofibroblast.^71^ These pieces of evidence point to the cell-specific role of telomerase genes in IPF development and suggest the importance of telomerase, telomere length and early cell senescence in IPF.

Our results implicated significant association of variants in cell adhesion molecules (*DSP* and *DPP9*) with IPF susceptibility. *DSP* intronic variant rs2076295 (G) (OR = 1.31) and *DPP9* intronic variant rs12610495 (G) (OR = 1.29) shows strong association. *DSP* gene encodes desmoplakin, a component of desmosomes (a cell-cell adhesion complex) enabling tissues to resist mechanical stress. *DSP* is known to regulate cell-cell adhesion, epithelial barrier function, wound repair and structural integrity of epithelial cells. Prior evidence that loss of AEC integrity and epithelial barrier dysfunction leads to dysregulated repair process and altered epithelial-mesenchymal communication with subsequent progressive fibrosis, has pointed to the role of cell adhesion molecules in IPF pathogenesis.^72^ DSP is shown to be localized in various lung epithelial cell types relevant for IPF like bronchial epithelial cells and AEC. *DSP* intronic variant was associated with reduced *DSP* expression in both IPF and control lung tissue.^12, 73^ *In vitro* studies have shown that loss of *DSP* can lead to decreased cellular integrity, enhanced EMT and expression of ECM genes.^73^ Another cell adhesion-related gene, *DPP9* encodes dipeptidyl peptidase 9 which is expressed in epithelia and has a role in cell adhesion, migration and apoptosis.^74^ Studies have shown that there was nominal *DPP9* overexpression in IPF lung versus healthy lung,^12^ but rs12610495 was not found to be associated with *DPP9* expression.^12^ Thus, dysregulated expression of cell adhesion molecules could contribute to IPF, but the molecular mechanism remains poorly understood.

The risk allele ‘G’ of *FAM13A* intronic variant rs2609255 (OR = 1.37) shows a 1.37-fold increased risk of IPF. *FAM13A* is reported to be expressed in the lung, especially in mucosal cells, club cells, airway epithelial cells and AEC.^75, 76^ Studies have shown that *FAM13A* was downregulated in bleomycin-induced PF in mouse lungs and in *FAM13A* deficient mice, PF was exacerbated by enhancing EMT.^77^ *FAM13A* was known to increase β-catenin degradation by enhancing its phosphorylation.^76^ Thus, loss of *FAM13A* can exaggerate bleomycin-induced PF potentially through activating the β-catenin pathway which is known to accelerate EMT.^77^

In the present meta-analysis, the coding variant rs1800470 in *TGF-β1* (transforming growth factor-beta1), a gene involved in EMT (OR = 1.07) is nominally associated with IPF. *TGF-β1* is a multifunctional cytokine that might play an important role in IPF as its overexpression was reported in both fibrotic human lung and animal models of PF. Cellular sources of *TGF-β1* during PF development include alveolar macrophages, bronchial epithelium and AEC type II. Various studies had reported that *TGF-β1* can promote PF by promoting EMT, AEC apoptosis, protecting myofibroblasts against apoptosis and increasing ECM production.^78, 79^ In addition to this, it also reduces the breakdown of ECM by inhibiting the generation of matrix metalloproteinases and plasminogen activators, as well as by enhancing the expression of tissue inhibitors of metalloproteases and plasminogen activator inhibitor-1. Thus, overexpression of *TGF-β1* might induce PF development.

Several limitations should be considered while interpreting our results. Firstly, our literature search was limited to English articles and articles whose full text is available, it is possible that some studies might have been overlooked. However, non-English articles represent a small proportion of studies. Thus, their inclusion might not have a significant influence on the main results. Second, in this meta-analysis heterogeneity across studies was common. Although subgroup analysis based on ethnicity was done to possibly remove some variability, other sources of heterogeneity were not examined due to limited data. Third, the calculation of pooled estimate in meta-analyses is based on allelic contrasts only. As for most complex disease genes, the underlying mode of inheritance is unknown. Allele counts and unadjusted estimates of effect were used, rather than adjusted estimates of the association. We consider our approach to be a reasonable compromise between loss in power and practicality.

In summary, our study represents the first and only comprehensive systematic meta-analyses of the current literature on the genetic architecture of IPF susceptibility. Our findings provide useful data for designing future studies and may serve as an ideal resource for selecting the most prominent genetic risk variants to include in future risk prediction models in an effort to improve risk stratification and the cost-effectiveness of screening campaigns.

## Data Availability

All data produced in the present work are contained in the manuscript

## ACKNOWLEDGEMENT

We thank the Director, CSIR-Institute of Genomics and Integrative Biology (IGIB), for his scientific vision, motivation and support. This work was supported by the Council of Scientific and Industrial Research (CSIR) [grant numbers OLP 1154]. PS acknowledge CSIR, Government of India for providing their fellowships. DG acknowledge ICMR, Govt. of India. We thank the anonymous reviewers for their helpful suggestions in improving the manuscript.

## AUTHOR CONTRIBUTIONS

RK and PS devised the concept of the review. RK reviewed the manuscript and supervised the overall study till the final manuscript preparation. PS and DG did the literature search, reviewed articles, and extracted the data. PS prepared the original draft and performed the analysis. DG and BP reviewed the manuscript. PS and DG edited the manuscript. PS and DG visualized, designed, and generated the Figures. PS and DG prepared the tables and supplementary material. RK provided critical insights to improve the analysis.

## ETHICAL STATEMENT

Ethical review and approval is not applicable to this study, due to it being a meta-analysis of existing studies. PROSPERO registration: (registration number: CRD42022297970) at crd.york.ac.uk/prospero/

## DATA AVAILABILITY STATEMENT

The data that supports the findings of this study are available in the supplementary material of this article.

## ABBREVIATIONS

IPF: idiopathic pulmonary fibrosis
ILD: interstitial lung diseases
ECM: extracellular matrix genes
GWAS: genome-wide association studies
SNP: single nucleotide polymorphism
PRISMA: Preferred Reporting Items for Systematic Reviews and Meta-Analyses
MeSH: medical subject heading
NOS: Newcastle-Ottawa scale
OR: odds ratio
CI: confidence interval
EMT: epithelial-mesenchymal transition
MCC: mucociliary clearance
AEC: alveolar epithelial cells
PF: pulmonary fibrosis.

## REFERENCES

1. Martinez FJ, Collard HR, Pardo A, Raghu G, Richeldi L, Selman M, et al. Idiopathic pulmonary fibrosis. Nat. Rev. Dis. Primers. 2017; 3(1): 1–9.

2. Raghu G, Rochwerg B, Zhang Y, Garcia CAC, Azuma A, Behr J, et al. An official ATS/ERS/JRS/ALAT clinical practice guideline: Treatment of idiopathic pulmonary fibrosis: An update of the 2011 clinical practice guideline. Am J Respir Crit Care Med. 2015; 192(2): e3–19.

3. Khor YH, Ng Y, Barnes H, Goh NSL, McDonald CF, Holland AE. Prognosis of idiopathic pulmonary fibrosis without anti-fibrotic therapy: A systematic review. Eur Respir Rev. European Respiratory Society; 2020; 29(157): 1–16.

4. Nathan SD, Shlobin OA, Weir N, Ahmad S, Kaldjob JM, Battle E, et al. Long-term course and prognosis of idiopathic pulmonary fibrosis in the new millennium. Chest. 2011; 140(1): 221–9.

5. Dempsey TM, Sangaralingham LR, Yao X, Sanghavi D, Shah ND, Limper AH. Clinical Effectiveness of Antifibrotic Medications for Idiopathic Pulmonary Fibrosis. Am J Respir Crit Care Med. 2019; 200(2): 168–74.

6. Spagnolo P, Grunewald J, du Bois RM. Genetic determinants of pulmonary fibrosis: Evolving concepts. Lancet Respir. Med. 2014; 2: 416–28.

7. Fukuhara A, Tanino Y, Ishii T, Inokoshi Y, Saito K, Fukuhara N, et al. Pulmonary fibrosis in dyskeratosis congenita with TINF2 gene mutation. Eur. Respir. J. 2013; 42; 1757–9.

8. DePinho RA, Kaplan KL. The Hermansky-Pudlak syndrome. Report of three cases and review of pathophysiology and management considerations. Medicine. 1985; 64(3): 192– 202.

9. Javaheri S, Lederer DH, Pella JA, Mark GJ, Levine BW. Idiopathic pulmonary fibrosis in monozygotic twins. The importance of genetic predisposition. Chest. 1980; 78(4): 591–4.

10. Bonanni PP, Frymoyer JW, Jacox RF. A family study of idiopathic pulmonary fibrosis: a possible dysproteinemic and genetically determined disease. Am. J. Med. 1965; 39(3): 411–21.

11. Allen RJ, Porte J, Braybrooke R, Flores C, Fingerlin TE, Oldham JM, et al. Genetic variants associated with susceptibility to idiopathic pulmonary fibrosis in people of European ancestry: a genome-wide association study. Lancet Respir. Med. 2017; 5(11): 869–80.

12. Fingerlin TE, Murphy E, Zhang W, Peljto AL, Brown KK, Steele MP, et al. Genome-wide association study identifies multiple susceptibility loci for pulmonary fibrosis. Nat. Genet. 2013; 45(6): 613–20.

13. Moore C, Blumhagen RZ, Yang I v., Walts A, Powers J, Walker T, et al. Resequencing study confirms that host defense and cell senescence gene variants contribute to the risk of idiopathic pulmonary fibrosis. Am J Respir Crit Care Med. 2019; 200(2): 199–208.

14. Mathai SK, Pedersen BS, Smith K, Russell P, Schwarz MI, Brown KK, et al. Desmoplakin Variants Are Associated with Idiopathic Pulmonary Fibrosis. Am J Respir Crit Care Med. 2016; 193(10): 1151–60.

15. Noth I, Zhang Y, Ma SF, Flores C, Barber M, Huang Y, et al. Genetic variants associated with idiopathic pulmonary fibrosis susceptibility and mortality: A genome-wide association study. Lancet Respir. Med. 2013; 1(4): 309–17.

16. Mushiroda T, Wattanapokayakit S, Takahashi A, Nukiwa T, Kudoh S, Ogura T, et al. A genome-wide association study identifies an association of a common variant in TERT with susceptibility to idiopathic pulmonary fibrosis. J. Med. Genet. 2008; 45(10): 654–6.

17. Korthagen NM, van Moorsel CHM, Barlo NP, Kazemier KM, Ruven HJT, Grutters JC. Association between variations in cell cycle genes and idiopathic pulmonary fibrosis. PLoS One. 2012; 7(1).

18. Hutton B, Salanti G, Caldwell DM, Chaimani A, Schmid CH, Cameron C, et al. The PRISMA extension statement for reporting of systematic reviews incorporating network meta-analyses of health care interventions: checklist and explanations. Ann Intern Med. 2015; 162(11): 777–84.

19. Sagoo GS, Little J, Higgins JPT. Systematic reviews of genetic association studies. PLoS Med. 2009; 6: 0239–45.

20. Wells G, Shea B, O’Connell D, Robertson J, Peterson J, Welch V, et al. The Newcastle-Ottawa Scale (NOS) for assessing the quality of nonrandomised studies in meta-analyses.

21. Stang A. Critical evaluation of the Newcastle-Ottawa scale for the assessment of the quality of nonrandomized studies in meta-analyses. Eur. J. Epidemiol. 2010; 25: 603–5.

22. Higgins JPT, Thompson SG. Quantifying heterogeneity in a meta-analysis. Stat. Med. 2002; 21(11): 1539–58.

23. Higgins JP, Thompson SG, Deeks JJ, Altman DG. Measuring inconsistency in meta-analyses. BMJ. 2003; 327(7414): 557–60.

24. Kelley GA KK. Statistical models for meta-analysis: A brief tutorial. World J Methodol. 2012; 2(4): 27–32.

25. Egger M, Smith GD, Schneider M, Minder C. Bias in meta-analysis detected by a simple, graphical test. Bmj. 1997; 315(7109): 629–34.

26. Langevin SM, Ioannidis JPA, Vineis P, Taioli E. Assessment of cumulative evidence for the association between glutathione S-transferase polymorphisms and lung cancer: Application of the Venice interim guidelines. Pharmacogenet. Genomics. 2010; 20(10): 586–97.

27. Allen RJ, Guillen-Guio B, Oldham JM, Ma SF, Dressen A, Paynton ML, et al. Genome-wide association study of susceptibility to idiopathic pulmonary fibrosis. Am J Respir Crit Care Med. 2020; 201(5): 564–74.

28. Arimura-Omori M, Kiyohara C, Yanagihara T, Yamamoto Y, Ogata-Suetsugu S, Harada E, et al. Association between telomere-related polymorphisms and the risk of IPF and COPD as a precursor lesion of lung cancer: Findings from the fukuoka tobacco-related lung disease (fold) registry. Asian Pac. J. Cancer Prev. 2020; 21(3): 667–73.

29. American Thoracic Society, European Respiratory Society. Idiopathic pulmonary fibrosis: diagnosis and treatment: international consensus statement. Am J Respir Crit Care Med. 2000; 161: 646–664.

30. American Thoracic Society, European Respiratory Society. American Thoracic Society/European Respiratory Society international multidisciplinary consensus classification of the idiopathic interstitial pneumonias. Am J Respir Crit Care Med. 2002; 165: 277–304.

31. Demedts M, Costabel U. ATS/ERS international multidisciplinary consensus classification of the idiopathic interstitial pneumonias. Eur. Respir. J. 2002; 19(5): 794–796

32. Raghu G, Collard HR, Egan JJ, Martinez FJ, Behr J, Brown KK, et al. An official ATS/ERS/JRS/ALAT statement: idiopathic pulmonary fibrosis: evidence-based guidelines for diagnosis and management. Am J Respir Crit Care Med. 2011; 183: 788– 824

33. Travis WD, Costabel U, Hansell DM, King TE Jr, Lynch DA, Nicholson AG, et al. An official American Thoracic Society/European Respiratory Society statement: update of the international multidisciplinary classification of the idiopathic interstitial pneumonias. Am J Respir Crit Care Med. 2013; 188: 733–48.

34. Raghu G, Remy-Jardin M, Myers JL, Richeldi L, Ryerson CJ, Lederer DJ, et al. Diagnosis of idiopathic pulmonary fibrosis. An official ATS/ERS/JRS/ALAT clinical practice guideline. Am J Respir Crit Care Med. 2018; 198(5): e44–68.

35. Bournazos S, Bournazou I, Murchison JT, Wallace WA, McFarlane P, Hirani N, et al. Copy number variation of FCGR3B is associated with susceptibility to idiopathic pulmonary fibrosis. Respiration. 2011; 81(2): 142–9.

36. Bradley B, Branley HM, Egan JJ, Greaves MS, Hansell DM, Harrison NK, et al. Interstitial lung disease guideline: the British Thoracic Society in collaboration with the Thoracic Society of Australia and New Zealand and the Irish Thoracic Society. Thorax. 2008; 63: v1–v58.

37. Libby DM, Gibofsky A, Fotino M, Waters SJ, Smith JP. Immunogenetic and clinical findings in idiopathic pulmonary fibrosis: association with the B-cell alloantigen HLA-DR2. Am. Rev. Respir. Dis. 1983; 127(5): 618–22.

38. Dabar G, Aoun Z, Riachi M, Boutros J, Khayat G. Effect of MUC5B gene polymorphism on survival in a Lebanese population of patients with idiopathic pulmonary fibrosis. 2008

39. Bonella F, Campo I, Zorzetto M, Boerner E, Ohshimo S, Theegarten D, et al. Potential clinical utility of MUC5B und TOLLIP single nucleotide polymorphisms (SNPs) in the management of patients with IPF. Orphanet J. Rare Dis. 2021; 16(1).

40. Deng Y, Li Z, Liu J, Wang Z, Cao Y, Mou Y, et al. Targeted resequencing reveals genetic risks in patients with sporadic idiopathic pulmonary fibrosis. Hum. Mutat. 2018; 39(9): 1238–45.

41. Dressen A, Abbas AR, Cabanski C, Reeder J, Ramalingam TR, Neighbors M, et al. Analysis of protein-altering variants in telomerase genes and their association with MUC5B common variant status in patients with idiopathic pulmonary fibrosis: a candidate gene sequencing study. Lancet Respir. Med. 2018; 6(8): 603–14.

42. Petrovski S, Todd JL, Durheim MT, Wang Q, Chien JW, Kelly FL, et al. An exome sequencing study to assess the role of rare genetic variation in pulmonary fibrosis. Am J Respir Crit Care Med. 2017; 196(1): 82–93.

43. Kishore A, Žižková V, Kocourková L, Petrkova J, Bouros E, Nunes H, et al. Association study for 26 candidate loci in idiopathic pulmonary fibrosis patients from four European populations. Front. immunol. 2016; 7: 274.

44. Van der Vis JJ, Snetselaar R, Kazemier KM, ten Klooster L, Grutters JC, van Moorsel CH. Effect of Muc5b promoter polymorphism on disease predisposition and survival in idiopathic interstitial pneumonias. Respirology. 2016; 21(4): 712–7.

45. Horimasu Y, Ohshimo S, Bonella F, Tanaka S, Ishikawa N, Hattori N, et al. MUC5B promoter polymorphism in Japanese patients with idiopathic pulmonary fibrosis. Respirology. 2015; 20(3): 439–44.

46. Jiang H, Hu Y, Shang L, Li Y, Yang L, Chen Y. Association between MUC5B polymorphism and susceptibility and severity of idiopathic pulmonary fibrosis. Int. J. Clin. Exp. Pathol. 2015; 8(11): 14953.

47. Wang C, Zhuang Y, Guo W, Cao L, Zhang H, Xu L, et al. Mucin 5B promoter polymorphism is associated with susceptibility to interstitial lung diseases in Chinese males. PloS One. 2014; 9(8): e104919.

48. Coghlan MA, Shifren A, Huang HJ, Russell TD, Mitra RD, Zhang Q, et al. Sequencing of idiopathic pulmonary fibrosis-related genes reveals independent single gene associations. BMJ Open Respir. Res. 2014; 1(1).

49. Wei R, Li C, Zhang M, Jones-Hall YL, Myers JL, Noth I, et al. Association between MUC5B and TERT polymorphisms and different interstitial lung disease phenotypes. Transl Res. 2014; 163(5): 494–502.

50. Stock CJ, Sato H, Fonseca C, Banya WAS, Molyneaux PL, Adamali H, et al. Mucin 5B promoter polymorphism is associated with idiopathic pulmonary fibrosis but not with development of lung fibrosis in systemic sclerosis or sarcoidosis. Thorax. 2013; 68(5): 436–41.

51. Borie R, Crestani B, Dieude P, Nunes H, Allanore Y, Kannengiesser C, et al. The MUC5B Variant Is Associated with Idiopathic Pulmonary Fibrosis but Not with Systemic Sclerosis Interstitial Lung Disease in the European Caucasian Population. PLoS One. 2013; 8(8).

52. Zhang Y, Noth I, Garcia JGN, Kaminski N. A Variant in the Promoter of MUC5B and Idiopathic Pulmonary Fibrosis. N. Engl. J. Med. 2011; 364(16): 1576–7.

53. Guzmán-Vargas J, Ambrocio-Ortiz E, Pérez-Rubio G, Ponce-Gallegos MA, Hernández-Zenteno R de J, Mejía M, et al. Differential Genomic Profile in TERT, DSP, and FAM13A Between COPD Patients With Emphysema, IPF, and CPFE Syndrome. Front. Med. 2021; 8.

54. Zhang HP, Zou J, Xie P, Gao F, Mu HJ. Association of HLA and cytokine gene polymorphisms with idiopathic pulmonary fibrosis. Kaohsiung J. Med. Sci. 2015; 31(12): 613–20.

55. Son JY, Kim SY, Cho SH, Shim HS, Jung JY, Kim EY, et al. TGF-β1 T869C polymorphism may affect susceptibility to idiopathic pulmonary fibrosis and disease severity. Lung. 2013; 191(2): 199–205.

56. Alhamad EH, Cal JG, Shakoor Z, Almogren A, AlBoukai AA. Cytokine gene polymorphisms and serum cytokine levels in patients with idiopathic pulmonary fibrosis. BMC Med. Genet. 2013; 14(1).

57. Li XX, Li N, Ban CJ, Zhu M, Xiao B, Dai HP. Idiopathic pulmonary fibrosis in relation to gene polymorphisms of transforming growth factor-β1 and plasminogen activator inhibitor 1. Chin. Med. J. 2011; 124(13): 1923–7.

58. Xaubet A, Marin-Arguedas A, Lario S, Ancochea J, Morell F, Ruiz-Manzano J, et al. Transforming growth factor-β1 gene polymorphisms are associated with disease progression in idiopathic pulmonary fibrosis. Am J Respir Crit Care Med. 2003; 168(4): 431–5.

59. Hirano C, Ohshimo S, Horimasu Y, Iwamoto H, Fujitaka K, Hamada H, et al. FAM13A polymorphism as a prognostic factor in patients with idiopathic pulmonary fibrosis. Respir. Med. 2017; 123: 105–9.

60. Lee MG, Lee YH. A meta-analysis examining the association between the MUC5B rs35705950 T/G polymorphism and susceptibility to idiopathic pulmonary fibrosis. Inflamm. Res. 2015; 64(6): 463–70.

61. Zhu QQ, Zhang XL, Zhang SM, Tang SW, Min HY, Yi L, et al. Association between the MUC5B promoter polymorphism rs35705950 and idiopathic pulmonary fibrosis: A meta-analysis and trial sequential analysis in Caucasian and Asian populations. Medicine. 2015; 94(43).

62. Evans CM, Fingerlin TE, Schwarz MI, Lynch D, Kurche J, Warg L, et al. Idiopathic pulmonary fibrosis: a genetic disease that involves mucociliary dysfunction of the peripheral airways. Physiol. Rev. 2016; 96(4): 1567–91.

63. Hancock LA, Hennessy CE, Solomon GM, Dobrinskikh E, Estrella A, Hara N, et al. Muc5b overexpression causes mucociliary dysfunction and enhances lung fibrosis in mice. Nat. Commun. 2018; 9(1): 1–10.

64. Dobrinskikh E, Walts AD, Schwartz DA. MUC5B Expression in IPF Is Associated with ER Stress. *In: D27 redox and beyond: mitochondria, er stress, and proteasomes*. American Thoracic Society. 2018; A6354–A6354.

65. Ma X, Dobrinskikh E, Kurche JS, Stancil IT, Kim E, Yang IV, et al. Endoplasmic Reticulum Stress in MUC5B-Driven Lung Fibrosis. In TP107 pulmonary fibrosis: molecular and cellular mechanisms. American Thoracic Society. 2021; A4216–A4216.

66. Dai J, Cai H, Zhuang Y, Wu Y, Min H, Li J, et al. Telomerase gene mutations and telomere length shortening in patients with idiopathic pulmonary fibrosis in a Chinese population. Respirology. 2015; 20(1): 122–8.

67. Codd V, Nelson CP, Albrecht E, Mangino M, Deelen J, Buxton JL, et al. Identification of seven loci affecting mean telomere length and their association with disease. Nat. Genet. 2013; 45(4): 422–7.

68. Iles MM, Bishop DT, Taylor JC, Hayward NK, Brossard M, Cust AE, et al. The effect on melanoma risk of genes previously associated with telomere length. J Natl Cancer Inst. 2014; 106(10).

69. Waisberg DR, Barbas-Filho JV, Parra ER, Fernezlian S, de Carvalho CR, Kairalla RA, et al. Abnormal expression of telomerase/apoptosis limits type II alveolar epithelial cell replication in the early remodeling of usual interstitial pneumonia/idiopathic pulmonary fibrosis. Hum. Pathol. 2010; 41(3): 385–91.

70. Liu T, Myoung JC, Ullenbruch M, Yu H, Jin H, Hu B, et al. Telomerase activity is required for bleomycin-induced pulmonary fibrosis in mice. J. Clin. Investig. 2007; 117(12): 3800– 9.

71. Liu T, Yu H, Ding L, Wu Z, de Los Santos FG, Liu J, et al. Conditional knockout of telomerase reverse transcriptase in mesenchymal cells impairs mouse pulmonary fibrosis. PLoS One. 2015; 10(11).

72. Camelo A, Dunmore R, Sleeman MA, Clarke DL. The epithelium in idiopathic pulmonary fibrosis: breaking the barrier. Front. pharmacol. 2014; 4: 173.

73. Hao Y, Bates S, Mou H, Yun JH, Pham B, Liu J, et al. Genome-wide association study: Functional variant rs2076295 regulates desmoplakin expression in airway epithelial cells. Am J Respir Crit Care Med. 2020; 202(9): 1225–36.

74. Yu DM, Wang XM, Ajami K, McCaughan GW, Gorrell MD. DP8 and DP9 have extra-enzymatic roles in cell adhesion, migration and apoptosis. *Dipeptidyl Aminopeptidases*. Springer, Boston, MA. 2006; 63–72.

75. Corvol H, Rousselet N, Thompson KE, Berdah L, Cottin G, Foussigniere T, et al. FAM13A is a modifier gene of cystic fibrosis lung phenotype regulating rhoa activity, actin cytoskeleton dynamics and epithelial-mesenchymal transition. J. Cyst. Fibros. 2018; 17(2): 190–203.

76. Jiang Z, Lao T, Qiu W, Polverino F, Gupta K, Guo F, et al. A chronic obstructive pulmonary disease susceptibility gene, FAM13A, regulates protein stability of β-catenin. Am J Respir Crit Care Med. 2016; 194(2): 185–97.

77. Rinastiti P, Ikeda K, Rahardini EP, Miyagawa K, Tamada N, Kuribayashi Y, et al. Loss of family with sequence similarity 13, member A exacerbates pulmonary hypertension through accelerating endothelial-to-mesenchymal transition. PloS One. 2020; 15(2): e0226049.

78. Aschner Y, Downey GP. Transforming growth factor-β: master regulator of the respiratory system in health and disease. Am. J. Respir. Cell Mol. Biol. 2016; 54(5): 647–55.

79. Kasai H, Allen JT, Mason RM, Kamimura T, Zhang Z. TGF-β1 induces human alveolar epithelial to mesenchymal cell transition (EMT). Respir. Res. 2005; 6.

