## Supplementary material for "Genetic landscape of idiopathic pulmonary fibrosis: A systematic review, meta-analysis and epidemiological evidence of case-control studies"

Ritushree Kukreti**,**

Genomics and Molecular Medicine Unit,

CSIR- Institute of Genomics and Integrative Biology,

Mall Road, Delhi 110 007, India.

**Supplementary files:**

1. **Supplementary Table 1:** PRISMA checklist
2. **Supplementary Fig. 1:** Two-stage search strategy to identify and select IPF gene association studies.
3. **Supplementary Table 2:** Literature search: MeSH terms used, individually and in combination, with AND/OR Boolean operators in I$^{st}$ and II$^{nd}$ literature search strategy.
4. **Supplementary file 1:** Inclusion and exclusion criteria.
5. **Supplementary file 2:** Data extraction.
6. **Supplementary file 3:** Literature search and study selection.
7. **Supplementary Table 3:** Criteria for evaluation of epidemiological credibility.
8. **Supplementary Fig. 2:** PRISMA flow diagram for the inclusion and exclusion of studies exploring the role of genetic variants in IPF risk. The number of studies excluded on each step is represented as N.
9. **Supplementary Table 4:** Characteristics of included studies for assessment of the association between genetic variants and IPF susceptibility.
10. **Supplementary Table 5:** Details of quality assessment scoring for studies included in the meta-analysis based on modified Newcastle Ottawa Scale (NOS).
11. **Supplementary Table 6:** Summary of subgroup analysis findings based on ethnicity between IPF patients and healthy controls for genetic variants- a) rs35705950 (*MUC5B*), b) rs2736100 (*TERT*), c) rs2609255 (*FAM13A*), d) rs1800470 (*TGF-β1*).
12. **Supplementary Table 7:** Sensitivity analysis with pooled odds ratios (OR) for studies exploring the association of genetic variants with IPF susceptibility by removing one study each time.
13. **Supplementary Fig. 3:** Funnel plots showing publication bias among studies exploring IPF risk associated with genetic variants- a) rs35705950 (*MUC5B*), b) rs2736100 (*TERT* ), c) rs2609255 (*FAM13A*), d) rs2076295 (*DSP*), e) rs12610495 (*DPP9*), f) rs1800470 (*TGF-β1*). Each point – represents a separate study for the indicated association or natural logarithm (log) of OR. A horizontal line means effect size.

**Supplementary Table 1**: PRISMA checklist

| Section and Topic | Item # | Checklist item | Page No. |
|---|---|---|---|
| **TITLE** | | | |
| Title | 1 | Identify the report as a systematic review. | 1 |
| **ABSTRACT** | | | |
| Abstract | 2 | See the PRISMA 2020 for Abstracts checklist. | 2 |
| **INTRODUCTION** | | | |
| Rationale | 3 | Describe the rationale for the review in the context of existing knowledge. | 3 |
| Objectives | 4 | Provide an explicit statement of the objective(s) or question(s) the review addresses. | 3 |
| **METHODS** | | | |
| Eligibility criteria | 5 | Specify the inclusion and exclusion criteria for the review and how studies were grouped for the syntheses. | 9(Supplementary file 1 of Supporting Information) |
| Information sources | 6 | Specify all databases, registers, websites, organisations, reference lists and other sources searched or consulted to identify studies. Specify the date when each source was last searched or consulted. | 4 |
| Search strategy | 7 | Present the full search strategies for all databases, registers and websites, including any filters and limits used. | 4, 7-8 (Table 2 of Supporting Information |
| Selection process | 8 | Specify the methods used to decide whether a study met the inclusion criteria of the review, including how many reviewers screened each record and each report retrieved, whether they worked independently, and if applicable, details of automation tools used in the process. | 4 |
| Data collection process | 9 | Specify the methods used to collect data from reports, including how many reviewers collected data from each report, whether they worked independently, any processes for obtaining or confirming data from study investigators, and if applicable, details of automation tools used in the process. | 5, 9 (Supplementary file 2 of Supporting Information) |
| Data items | 10a | List and define all outcomes for which data were sought. Specify whether all results that were | 9 |

| Section and Topic | Item # | Checklist item | Page No. |
|---|---|---|---|
| | | compatible with each outcome domain in each study were sought (e.g. for all measures, time points, analyses), and if not, the methods used to decide which results to collect. | (Supplementary file 2 of Supporting Information) |
| | 10b | List and define all other variables for which data were sought (e.g. participant and intervention characteristics, funding sources). Describe any assumptions made about any missing or unclear information. | |
| Study risk of bias assessment | 11 | Specify the methods used to assess risk of bias in the included studies, including details of the tool(s) used, how many reviewers assessed each study and whether they worked independently, and if applicable, details of automation tools used in the process. | 5 |
| Effect measures | 12 | Specify for each outcome the effect measure(s) (e.g. risk ratio, mean difference) used in the synthesis or presentation of results. | 5 |
| Synthesis methods | 13a | Describe the processes used to decide which studies were eligible for each synthesis (e.g. tabulating the study intervention characteristics and comparing against the planned groups for each synthesis (item #5)). | 4,5 |
| | 13b | Describe any methods required to prepare the data for presentation or synthesis, such as handling of missing summary statistics, or data conversions. | |
| | 13c | Describe any methods used to tabulate or visually display results of individual studies and syntheses. | |
| | 13d | Describe any methods used to synthesize results and provide a rationale for the choice(s). If meta-analysis was performed, describe the model(s), method(s) to identify the presence and extent of statistical heterogeneity, and software package(s) used. | |
| | 13e | Describe any methods used to explore possible causes of heterogeneity among study results (e.g. subgroup analysis, meta-regression). | |
| | 13f | Describe any sensitivity analyses conducted to assess robustness of the synthesized results. | |
| Reporting bias assessment | 14 | Describe any methods used to assess risk of bias due to missing results in a synthesis (arising from reporting biases). | 6 |
| Certainty assessment | 15 | Describe any methods used to assess certainty (or confidence) in the body of evidence for an outcome. | 5 |
| **RESULTS** | | | |
| Study selection | 16a | Describe the results of the search and selection process, from the number of records identified in the search to the number of studies included in the review, ideally using a flow diagram. | 6-7, Fig. 1, 9 (Supplementary file 3 of Supporting |

| Section and Topic | Item # | Checklist item | Page No. |
|---|---|---|---|
| | | | Information), 11 (Fig. 2 of Supporting Information) |
| | 16b | Cite studies that might appear to meet the inclusion criteria, but which were excluded, and explain why they were excluded. | 8 |
| Study characteristics | 17 | Cite each included study and present its characteristics. | 7, 12-36 (Table 4 of Supporting Information), 24-35 (Table 1) |
| Risk of bias in studies | 18 | Present assessments of risk of bias for each included study. | 10, 42-43 (Fig. S2), 10, 40-41 (Table 7 of supporting information) |
| Results of individual studies | 19 | For all outcomes, present, for each study: (a) summary statistics for each group (where appropriate) and (b) an effect estimate and its precision (e.g. confidence/credible interval), ideally using structured tables or plots. | 12-36 (Table 4 of Supporting Information) |
| Results of syntheses | 20a | For each synthesis, briefly summarise the characteristics and risk of bias among contributing studies. | 23-34 (Table 1) |
| | 20b | Present results of all statistical syntheses conducted. If meta-analysis was done, present for each the summary estimate and its precision (e.g. confidence/credible interval) and measures of statistical heterogeneity. If comparing groups, describe the direction of the effect. | 8-10, 35 (Table 2), 39 (Table 6 of Supporting Information) |
| | 20c | Present results of all investigations of possible causes of heterogeneity among study results. | 8-10, 35 (Table 2) |
| | 20d | Present results of all sensitivity analyses conducted to assess the robustness of the synthesized results. | 10, 39-41 (Table 7 of Supporting Information) |
| Reporting biases | 21 | Present assessments of risk of bias due to missing results (arising from reporting biases) for each synthesis assessed. | 10, 42-43 (Fig. 2) |
| Certainty of evidence | 22 | Present assessments of certainty (or confidence) in the body of evidence for each outcome assessed. | 10, 40-41 (Table 7 of supporting |

| Section and Topic | Item # | Checklist item | Page No. |
|---|---|---|---|
| | | | information) |
| **DISCUSSION** | | | |
| Discussion | 23a | Provide a general interpretation of the results in the context of other evidence. | 11-13 |
| | 23b | Discuss any limitations of the evidence included in the review. | 13-14 |
| | 23c | Discuss any limitations of the review processes used. | 13-14 |
| | 23d | Discuss implications of the results for practice, policy, and future research. | 14 |
| **OTHER INFORMATION** | | | |
| Registration and protocol | 24a | Provide registration information for the review, including register name and registration number, or state that the review was not registered. | 4 |
| | 24b | Indicate where the review protocol can be accessed, or state that a protocol was not prepared. | 4 |
| | 24c | Describe and explain any amendments to information provided at registration or in the protocol. | NA |
| Support | 25 | Describe sources of financial or non-financial support for the review, and the role of the funders or sponsors in the review. | 14 |
| Competing interests | 26 | Declare any competing interests of review authors. | 14 |
| Availability of data, code and other materials | 27 | Report which of the following are publicly available and where they can be found: template data collection forms; data extracted from included studies; data used for all analyses; analytic code; any other materials used in the review. | 15 |

**Supplementary Fig. 1:** Two-stage search strategy to identify and select IPF gene association studies.

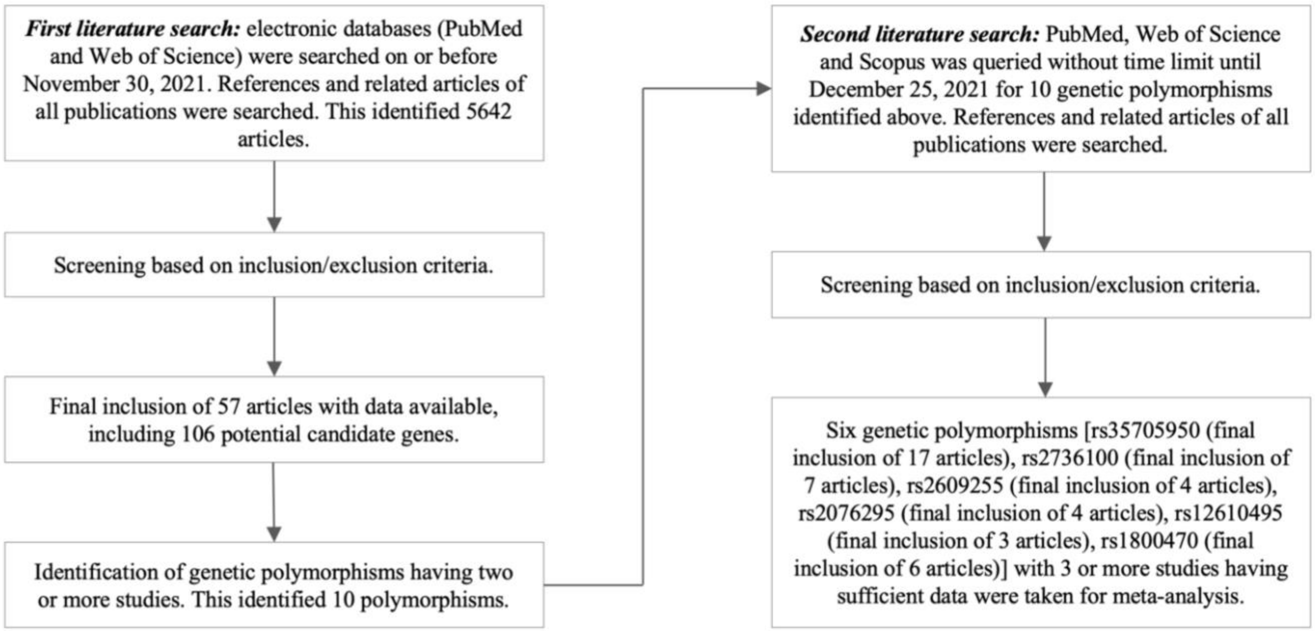


**Supplementary Table 2**: Literature search: MeSH terms used, individually and in combination, with AND/OR Boolean operators in I$^{st}$ and II$^{nd}$ literature search strategy.

| MeSH terms used for I$^{st}$ search strategy | |
|---|---|
| "idiopathic pulmonary fibrosis", "idiopathic interstitial pneumonia", "IPF", "genetics", "genetic", "gene", "genetic variation", "genetic variant", "genetic factors", "genetic association", "SNP", "single nucleotide polymorphism", "polymorphism" | |
| MeSH terms used for II$^{nd}$ search strategy | |
| rs35705950 (*MUC5B*) | "mucin5b", "MUC5B", "rs35705950", "mucin", "genetics", "genetic", "gene", "genetic variation", "genetic variant", "genetic factors", "genetic association", "SNP", "single nucleotide polymorphism", "polymorphism", "idiopathic pulmonary fibrosis", "idiopathic interstitial pneumonia", "IPF", "pulmonary fibrosis" |

| | |
|---|---|
| rs2736100 (*TERT*) | “telomerase reverse transcriptase”, “TERT”, “rs2736100”, “telomerase related gene”, “genetics”, “genetic”, “gene”, “genetic variation”, “genetic variant”, “genetic factors”, “genetic association”, “SNP”, “single nucleotide polymorphism”, “polymorphism”, “idiopathic pulmonary fibrosis”, “idiopathic interstitial pneumonia”, “IPF”, “pulmonary fibrosis” |
| rs2609255 (*FAM13A*) | “family with sequence similarity 13 member A”, “family with Sequence Similarity 13, Member A”, “FAM13A”, “rs2609255”, “genetics”, “genetic”, “gene”, “genetic variation”, “genetic variant”, “genetic factors”, “genetic association”, “SNP”, “single nucleotide polymorphism”, “polymorphism”, “idiopathic pulmonary fibrosis”, “idiopathic interstitial pneumonia”, “IPF”, “pulmonary fibrosis” |
| rs2076295 (*DSP*) | “desmoplakin”, “desmoplakins”, “DSP”, “rs2076295”, “cell-cell adhesion”, “genetics”, “genetic”, “gene”, “genetic variation”, “genetic variant”, “genetic factors”, “genetic association”, “SNP”, “single nucleotide polymorphism”, “polymorphism”, “idiopathic pulmonary fibrosis”, “idiopathic interstitial pneumonia”, “IPF”, “pulmonary fibrosis” |
| rs12610495 (*DPP9*) | “Dipeptidyl Peptidase 9”, “DPP9”, “rs12610495”, “DP9”, “genetics”, “genetic”, “gene”, “genetic variation”, “genetic variant”, “genetic factors”, “genetic association”, “SNP”, “single nucleotide polymorphism”, “polymorphism”, “idiopathic pulmonary fibrosis”, “idiopathic interstitial pneumonia”, “IPF”, “pulmonary fibrosis” |
| rs1800470 (*TGF-β1*) | “TGF-beta1”, “TGF-beta-1”, “TGFbeta”, “transforming growth factor beta1”, “TGFB1”, “rs1800470”, “genetics”, “genetic”, “gene”, “genetic variation”, “genetic variant”, “genetic factors”, “genetic association”, “SNP”, “single nucleotide polymorphism”, “polymorphism”, “idiopathic pulmonary fibrosis”, “idiopathic interstitial pneumonia”, “IPF”, “pulmonary fibrosis” |
| rs7934606 (*MUC2*) | “MUC2”, “mucin2”, “mucin-2”, “rs7934606”, “mucin”, “genetics”, “genetic”, “gene”, “genetic variation”, “genetic variant”, “genetic factors”, “genetic association”, “SNP”, “single nucleotide polymorphism”, “polymorphism”, “idiopathic pulmonary fibrosis”, “idiopathic interstitial pneumonia”, “IPF”, “pulmonary fibrosis” |
| rs1278769 (*ATP11A*) | “ATP11A”, “ATPase Phospholipid Transporting 11A”, “rs1278769”, “genetics”, “genetic”, “gene”, “genetic variation”, “genetic variant”, “genetic factors”, “genetic association”, “SNP”, “single nucleotide polymorphism”, “polymorphism”, “idiopathic pulmonary fibrosis”, “idiopathic interstitial pneumonia”, “IPF”, “pulmonary fibrosis” |
| rs5793295 (*LRRC34*) | “Leucine Rich Repeat Containing 34”, “Leucine-Rich Repeat-Containing 34”, “LRRC34”, “rs5793295”, “genetics”, “genetic”, “gene”, “genetic variation”, “genetic variant”, “genetic factors”, “genetic association”, “SNP”, “single nucleotide polymorphism”, “polymorphism”, “idiopathic pulmonary fibrosis”, “idiopathic interstitial pneumonia”, “IPF”, “pulmonary fibrosis” |
| rs111521887 (*TOLLIP*) | “TOLLIP”, “Toll Interacting Protein”, “rs111521887”, “Toll-Interacting Protein”, “genetics”, “genetic”, “gene”, “genetic variation”, “genetic variant”, “genetic factors”, “genetic association”, “SNP”, “single nucleotide polymorphism”, “polymorphism”, “idiopathic pulmonary fibrosis”, “idiopathic interstitial pneumonia”, “IPF”, “pulmonary fibrosis” |

**Supplementary file 1**: Inclusion and exclusion criteria

Articles meeting the following criteria were eligible to be included in the study: (i) studies evaluating the association of genetic variants in IPF susceptibility, (ii) the study design must be a case-control design based on unrelated individuals, (iii) IPF patients must have been diagnosed by radiological, histopathological examination or based on American Thoracic Society (ATS)/ European Respiratory Society (ERS)/ Japanese Respiratory Society (JRS)/ Latin-American Thoracic Association (ALAT) diagnostic criteria. Articles were excluded based on the following criteria (i) review articles, meta-analysis, editorials, commentary, errata, and news articles, (ii) study design based on family or sibling pairs, (iii) non-human studies, (iv) non-English articles, (v) recruited IPF patients with comorbid conditions. Additionally, for meta-analyses, articles were excluded if (vi) insufficient data was provided for estimating odds ratios (OR) with 95% confidence interval (CI), (vii) the genotype distribution in controls was inconsistent with Hardy-Weinberg equilibrium (HWE).

**Supplementary file 2**: Data extraction

The following data were extracted: first author, year of publication, ethnicity of the studied population (where not conveyed, the country in which the study was carried out was considered), diagnostic criteria for IPF patients, number of participants (total and male/female), the mean age of cases and controls, genes, genetic variants, risk allele or genotype, statistical parameters [p-value, OR and 95% CI] of allelic model (for insufficient data for calculation, genotypic data is represented), genotyping methods used. Global allele frequency of risk allele was determined according to the dbSNP database (54). Authors were contacted where data required for conducting meta-analysis was not provided. Publications were excluded if responses were not received after two weeks. In the case of repeated or overlapping studies, the one with the most subjects and sufficient data was included.

**Supplementary file 3**: Literature search and study selection

Out of 4032 excluded studies, 908 studies were grey literature (Review, systematic review and meta-analysis), one study is in a language other than English, twenty-nine studies discussed IPF with comorbid condition, thirty studies were on familial IPF/pulmonary fibrosis (PF)/ idiopathic interstitial pneumonias (IIP) (as the current study focuses on sporadic IPF), 109 studies were conducted on animal or *in vitro* models for IPF, 1637 studies were not related to IPF, 1112 studies were related to IPF but do not talk about genetics, 104 non-genetic studies, nine studies talked about the genetics of IPF progression and survival, seventy-eight studies were related to therapeutics and drugs used in IPF, in twelve studies abstract was not found and three studies had non-significant results for the association of genetic variants and IPF susceptibility.

**Supplementary Table 3:** Criteria for evaluation of epidemiological credibility.

| **Criteria** | **Categories** |
|---|---|
| Amount of evidence | A: Sample size* for the rarer allele in the meta-analysis is greater than 1000. |
| | B: Sample size* for the rarer allele in the meta-analysis is between 100-1000. |
| | C: Sample size* for the rarer allele in the meta-analysis is less than 100. |
| Replication | A: Extensive replication including at least one well-conducted meta-analysis, and similarity of phenotyping, genotyping and analytical models across studies. |
| | B: Well replicated without any conducted meta-analysis, and similarity of phenotyping, genotyping and analytical models across studies. |
| | C: No association, failed replication, and no independent replication. |
| Protection from bias | A: The summary effect size is greater than 1.15 or less than 0.87, and the following four situations do not occur: 1) nominal statistical significance is lost with the exclusion of the first published study; 2) nominal statistical significance is lost with the exclusion of studies where HWE is violated; 3) small study effects. |
| | B: There is no evidence of bias that would invalidate an association, but important information is missing |
| | C: The summary effect size is 0.87–1.15, or at least one of the following four situations occurs: 1) nominal statistical significance is lost with the exclusion of the first published study; 2) nominal statistical significance is lost with the exclusion of studies where HWE is violated; 3) small study effects. |

*calculated by multiplying population specific minor allele frequency with total number of cases and controls included in meta-analysis. HWE, Hardy Weinberg equilibrium.

**Supplementary Fig. 2:** PRISMA flow diagram for the inclusion and exclusion of studies exploring the role of genetic variants in IPF risk. The number of studies excluded on each step is represented as N.

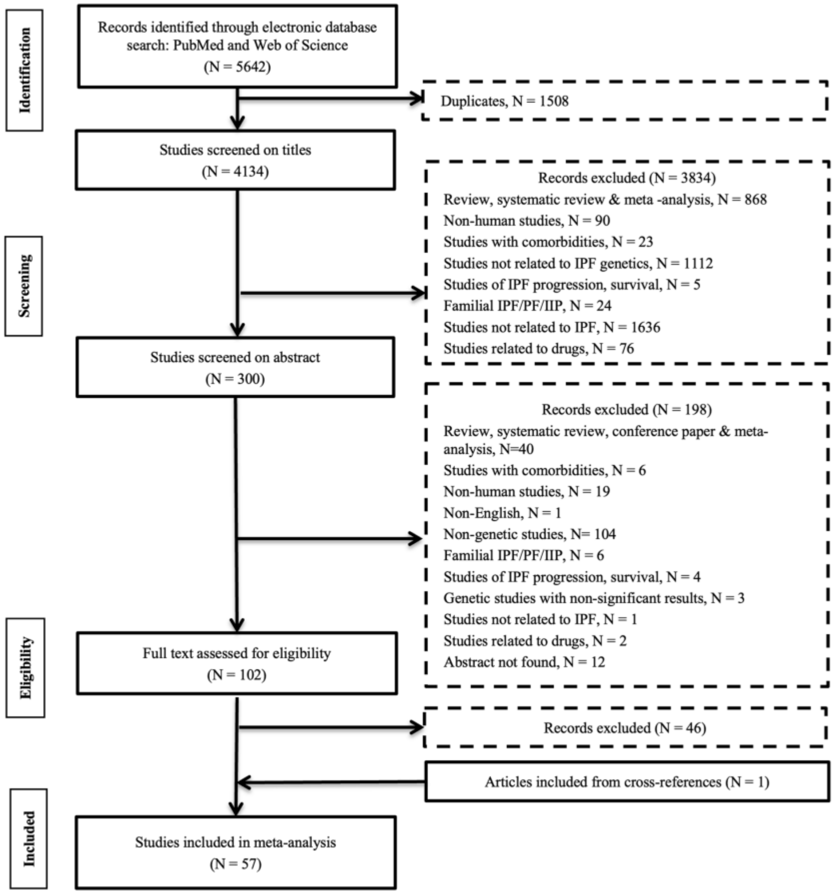

**Supplementary Table 4:** Characteristics of included studies for assessment of the association between genetic variants and IPF susceptibility.

| Sl. No | Study Name (year) | Population/ Ethnicity | Diagnostic criteria | Sample characteristics | | | | | | | Gene | SNP | Risk allele/ genotype | OR (95% CI) | p-value | Method | Global allele frequency (risk allele) |
|---|---|---|---|---|---|---|---|---|---|---|---|---|---|---|---|---|---|
| | | | | Total (N) | Case | | | Control | | | | | | | | | |
| | | | | | N | M/F | Age in years [mean±SD/ mean (range)] | N | M/F | Age in years [mean±SD /mean (range)] | | | | | | | |
| 1 | Dhindsa et al. (2021)[1] | European | ATS/ERS guidelines 2013 | 119,807 | 752 | - | 71.0 | 119,055 | - | - | *MUC2* | rs7944723 | C | 1.86 (1.66-2.07) | $1.71 \times 10^{-26}$ | Whole exome sequencing | 0.122 “ |
| | | | | | | | | | | | *MUC5AC* | rs28403537 | T | 2.78 (2.35-3.29) | $6.96 \times 10^{-26}$ | | 0.046 “ |
| | | | | | | | | | | | *MUC2* | rs6421972 | T | 1.41 (1.27-1.55) | $4.30 \times 10^{-11}$ | | 0.390 “ |
| | | | | | | | | | | | *TOLLIP* | rs3750920 | T | 1.37 (1.24-1.51) | $9.63 \times 10^{-10}$ | | 0.424 “ |
| | | | | | | | | | | | *MUC5B* | rs908229 | C | 0.75 (0.67-0.82) | $1.90 \times 10^{-8}$ | | 0.443 “ |
| | | | | | | | | | | | *SPDL1* | rs116483731 | A | 2.87 (2.03-4.07) | $2.40 \times 10^{-7}$ | | 0.006 “ |
| | | | | 198,014 | 1028 | - | - | 196,986 | - | - | *SPDL1* | rs116483731 | A | 3.13 (2.37-4.14) | $1.00 \times 10^{-15}$ | | 0.006 “ |
| 2 | Bonella et al. (2021)[2] | Caucasian | ATS/ERS/JRS/ALAT guidelines 2011 and ATS/ERS/JRS/ALAT guidelines 2018 | 112 | 62 | 43/8 | 63.5±11 | 50 | 37/13 | 42.0±2 | *MUC5B* | rs35705950 | T | 5.36 (2.46-11.69) | <0.0001 | RT-PCR using TaqMan | 0.046 |
| 3 | Allen et al. (2020)[G3] | European | (ATS/ERS guidelines 2000 and ATS/ERS/JRS/ALAT | 11259[D] | 2668 [541[α], 1,515[β], 612[γ]] | 1818/809[(355/145)[α], (1030/48 | (68.0±3.0/ 63.0±7.5)[α], 66.0±9.5[β], (70/65.0)[γ] | 8591 [542[α], 4,683[β], 3,366[γ]] | 4891/3,668 [(240/270)[α], (2295/23 | - | *KIF15* | rs78238620 | A | 1.58 (1.37-1.83) | $5.12 \times 10^{-10}$ | Affymetrix 6.0 SNP array[α]; Illumina Human 660W Quad BeadChip[β]; Affymetrix UK BiLEVE | 0.054 |
| | | | | | | | | | | | *MAD1L1* | rs12699415 | A | 1.28 (1.19-1.37) | $7.15 \times 10^{-13}$ | | 0.409 |
| | | | | | | | | | | | *DEPTOR* | rs28513081 | G | 0.82 (0.76-0.87) | $1{\cdot}20 \times 10^{-9}$ | | 0.454 |
| | | | | | | | | | | | *RTEL1* | rs41308092 | A | 2.12 (1.67-2.69) | $7.65 \times 10^{-10}$ | | 0.007 |

| | | | | | | | | | | | | | | | | | |
|---|---|---|---|---|---|---|---|---|---|---|---|---|---|---|---|---|---|
| | | | guidelines 2011)[α]; ATS/ERS criteria 2002[β]; (ATS/ERS guidelines 2002 and ATS/ERS/JRS/ALAT guidelines 2011)[γ] | | | 5)[β], (433/179)[γ]] | | | 88)[β], (2356/1010)[γ]] | | *HECTD2* | rs537322302 | G | 7.82 (3.77-16.2) | 3.43 $\times 10^{-8}$ | array(cases)[γ]/ Affymetrix UK BiLEVE and UK Biobank arrays(control)[γ] | 0.0002 |
| | | | | | | | | | | | *RTEL1* | rs41308092 | A | 2.12 (1.67-2.69) | 7.65 $\times 10^{-1}$ | | 0.008 |
| | | | | | | | | | | | *LRRC34/ TERC* | rs12696304 | G | 1.31 (1.21-1.40) | $7.09\times10^{-13}$ | | 0.470 |
| | | | | | | | | | | | *FAM13A* | rs2013701 | T | 0.78 (0.74-0.84) | $3.30\times10^{-13}$ | | 0.499 |
| | | | | | | | | | | | *TERT* | rs7725218 | A | 0.72 (0.67-0.77) | $1.54\times10^{-20}$ | | 0.408 |
| | | | | | | | | | | | *DSP* | rs2076295 | G | 1.46 (1.37-1.56) | $2.79\times10^{-30}$ | | 0.432 |
| | | | | | | | | | | | - | rs2897075 | T | 1.30 (1.21-1.38) | $3.10\times10^{-14}$ | | 0.310 |
| | | | | | | | | | | | *MUC5B* | rs35705950 | T | 4.84 (4.37-5.36) | $1.18\times10^{-203}$ | | 0.046 |
| | | | | | | | | | | | *ATP11A* | rs9577395 | G | 0.77 (0.71-0.83) | $1.34\times10^{-10}$ | | 0.166 |
| | | | | | | | | | | | *IVD* | rs59424629 | G | 0.77 (0.71-0.82) | $7.30\times10^{-16}$ | | 0.480 |
| | | | | | | | | | | | *AKAP13* | rs62023891 | A | 1.27 (1.18-1.36) | $1.27\times10^{-10}$ | | - |
| | | | | | | | | | | | *MAPT* | rs2077551 | C | 0.71 (0.65-0.77) | $2.83\times10^{-16}$ | | - |
| | | | | | | | | | | | *DPP9* | rs12610495 | G | 1.31 (1.22-1.42) | $2.92\times10^{-12}$ | | 0.183 |
| | | | ATS/ERS/JRS/ALAT guidelines 2011^ | 13330[R] | 1456 [792[@], 664^] | 1,070/386 [(582/210)[@], (488/176)^] | (69/58)[@], 68^ | 11874 [10,000[@], 1,874^] | 7,718/4,156 [(7210/2790)[@], (508/1366)^] | - | *MAD1L1* | rs12699415 | A | 1.29 (1.18-1.41) | 2.27 $\times 10^{-8}$ | Affymetrix UK Biobank and Spain Biobank arrays(cases)[@]; Affymetrix UK BiLEVE and UK Biobank arrays(control)[@]; HiSeq X Ten platform (Illumina)^ | 0.409 |
| | | | | | | | | | | | *KIF15* | rs78238620 | A | 1.48 (1.24-1.77) | $1.43\times10^{-5}$ | | 0.054 |
| | | | | | | | | | | | *DEPTOR* | rs28513081 | G | 0.87 (0.80-0.95) | 0.002 | | 0.454 |

| | | | | | | | | | | | | | | | | | |
|---|---|---|---|---|---|---|---|---|---|---|---|---|---|---|---|---|---|
| | | | (ATS/ERS guidelines 2000 and ATS/ERS/JRS/ALAT guidelines 2011)$^{\alpha}$; ATS/ERS guideline 2002$^{\beta}$; (ATS/ERS guidelines 2002 and ATS/ERS/JRS/ALAT guidelines 2011)$^{\gamma}$; ATS/ERS/JRS/ALAT guidelines 2011^ | 24589$^{M}$ | 4124 [541$^{\alpha}$, 1,515$^{\beta}$, 612$^{\gamma}$, 792$^{@}$, 664^] | 2888/1195 [(355/145)$^{\alpha}$, (1030/485)$^{\beta}$, (433/179)$^{\gamma}$, (582/210)$^{@}$, (488/176)^] | (68.0±3.0/63.0±7.5)$^{\alpha}$, 66.0±9.5$^{\beta}$, (70/65)$^{\gamma}$, (69/58)$^{@}$, 68^ | 20465 [542$^{\alpha}$, 4,683$^{\beta}$, 3,366$^{\gamma}$, 10,000$^{@}$, 1,874^] | 12,609/7,824 [(240/270)$^{\alpha}$, (2295/2388)$^{\beta}$, (2356/1010)$^{\gamma}$, (7210/2790)$^{@}$, (508/1366)^] | - | *KIF15* | rs78238620 | A | 1.54 (1.38-1.73) | 4.05 ×10$^{-14}$ | Affymetrix 6.0 SNP array$^{\alpha}$; Illumina Human 660W Quad BeadChip$^{\beta}$; Affymetrix UK BiLEVE array(cases)$^{\#\#\#}$/Affymetrix UK BiLEVE and UK Biobank arrays (control)$^{\gamma}$; Affymetrix UK Biobank and Spain Biobank arrays (cases)$^{@}$, Affymetrix UK BiLEVE and UK Biobank arrays (control)$^{@}$; HiSeq X Ten platform (Illumina)^ | 0.054 |
| | | | | | | | | | | | *MAD1L1* | rs12699415 | A | 1.28 (1.21-1.35) | 9.38 ×10$^{-20}$ | | 0.409 |
| | | | | | | | | | | | *DEPTOR* | rs28513081 | G | 0.83 (0.79-0.88) | 1.93 ×10$^{-11}$ | | 0.454 |
| | | | | | | | | | | | *RTEL1* | rs41308092 | A | 1.82 (1.51-2.19) | 2.24 ×10$^{-10}$ | | 0.007 |
| 4 | Arimura-Omori et al. (2020)[4] | Japanese | ATS/ERS/JRS/ALAT guidelines 2011 | 534 | 155 [include 7 familial IPF] | 113/42 | 74.0 (69-78) | 379 | 283/96 | 58 (48-65) | *TERT* | rs2736100 | T | 1.75 (1.31-2.33)~ | 0.0001~ | RT-PCR using TaqMan fluorescent probes | 0.515 |
| 5 | Moore et al. (2019)[5] | non-Hispanic whites | ATS/ERS/JRS/ALAT guidelines 2018 | 7588 | 3146 | NA | NA | 4442 | - | - | *TERC* | rs2293607 | CT vs TT; CC vs TT | 1.27 (1.15-1.41); 1.82 (1.50-1.41) | - | Deep sequencing (HiSeq 4000 sequencer) | 0.269 |
| | | | | | | | | | | | *FAM13A* | rs2609260 | CT vs TT; CC vs TT | 1.38 (1.25-1.54); 2.09 (1.65-2.64) | - | | 0.239 |
| | | | | | | | | | | | *TERT* | rs4449583 | CT vs CC; TT vs CC | 0.66 (0.59-0.73); 0.47 (0.39-0.56) | - | | 0.323 |

| | | | | | | | | | | | | | | | | | |
|---|---|---|---|---|---|---|---|---|---|---|---|---|---|---|---|---|---|
| | | | | | | | | | | | *DSP* | rs2076295 | GT vs TT; GG vs TT | 1.27 (1.13-1.43); 2.03 (1.78–2.33) | - | | 0.432 |
| | | | | | | | | | | | *ZKSCAN1* | rs6963345 | AG vs GG; AA vs GG | 1.35 (1.21–1.50); 1.69 (1.47–1.96) | - | | 0.301 |
| | | | | | | | | | | | *OBFC1/STN1* | rs2488000 | T | 0.70 (0.62–0.80) | - | | 0.108 |
| | | | | | | | | | | | *MUC5B* | rs35705950 | TG vs GG; TT vs GG | 5.31 (4.76-5.92); 19.01 (13.49-26.78) | - | | 0.046 |
| | | | | | | | | | | | *ATP11A* | rs1278769 | AG vs GG; AA vs GG | 0.79 (0.71–0.88); 0.65 (0.57–0.90) | - | | 0.233 |
| | | | | | | | | | | | *IVD* | rs35700143 | CT vs TT; CC vs TT | 0.76 (0.68–0.85); 0.65 (0.56–0.74) | - | | 0.491 |
| | | | | | | | | | | | *DPP9* | rs12610495 | GA vs AA; GG vs AA | 1.25 (1.13–1.38); 1.64 (1.39–1.93) | - | | 0.183 |
| 6 | Liu et al. (2020)[6] | Chinese | ATS/ERS/JRS/ALAT guidelines 2011 | 378 | 253 | 169/84 | 65.4±11.1 | 125 | 84/41 | 65.3±10.8 | *HYAL1* | rs117179004 | A | 2.27 (1.27–4.07) | 0.006 | Ion Torrent platform (Thermo Fisher) | 0.013 |
| | | | | 387 | 162 | 99/63 | 64.2±10.9 | 225 | 109/73 | 60.9±10.5 | *HYAL1* | rs117179004 | A | 2.18 (1.32–3.61) | 0.002 | Big Dye v.1.1 terminator cycle sequencing kit and Applied Biosystems 3500xl capillary sequencer (Applied Biosystems) | 0.013 |
| 7 | Deng et al. (2019)[7] | Han Chinese | ATS/ERS/JRS/ALAT guidelines 2011 | 378 | 253 | 170/83 | 65.4±11.1 [age means onset age] | 125 | 84/41 | 65.3±10.8 | *CR1* | rs3737002 | T | 1.77 (1.28-2.45) | 0.001 | Ion Torrent Proton | 0.248 |
| | | | | | | | | | | | *CR1* | rs2296160 | G | 1.54 (1.13-2.09) | 0.006 | | 0.171 |
| | | | | | | | | | | | *TGF-β1* | rs1800470 | A | 1.47 (1.08-2.00) | 0.013 | | 0.454 |
| | | | | | | | | | | | *MUC5B* | rs35705950 | T | 4.84 (1.12-20.94) | 0.018 | | 0.046 |

| | | | | | | | | | | | | | | | | | |
|---|---|---|---|---|---|---|---|---|---|---|---|---|---|---|---|---|---|
| 8 | Wang et al. (2019)[8] | Chinese | ATS/ERS/JRS/ALAT guidelines 2018 | 279 | 79 | 68/11 | 64.2±8.86 | 200 | NA | NA | *MUC5B* | rs868903 | C | 1.46 (1.01-2.13)~ | 0.042 | PCR | 0.489 |
| | | | | | | | | | | | *TERT* | rs2853676 | A | 0.57 (0.33-0.98)~ | 0.041 | | 0.251 |
| 9 | Lorenzo-Salazar et al. (2019)[9] | European-American | ATS/ERS/JRS/ALAT guidelines 2011 | 652 (60.8% samples were from Noth I. et al., 2013) | 181 | 138/43 | 67.0 | 501 | - | - | *MUC5AC* | rs371630624 | C | 1942(245.6-15360) | $7.18 \times 10^{-13}$ | HiSeq 2500 (Illumina, Inc.) | 0.000 |
| | | | | | | | | | | | | rs34474233 | A | 4.08 (2.56-6.49) | $2.99 \times 10^{-9}$ | | 0.022 |
| | | | | | | | | | | | | rs34815853 | A | 4.01 (2.52-6.38) | $4.15 \times 10^{-9}$ | | 0.021 |
| | | | | | | | | | | | *MUC5B* | rs12802931 | G | 3.76 (2.73-5.16) | $3.72 \times 10^{-16}$ | | 0.073 |
| | | | | | | | | | | | | rs35705950 | T | 6.18 (4.28-8.93) | $2.69 \times 10^{-22}$ | | 0.046 |
| | | | | | | | | | | | *MUC5B/RP11-532E4.2* | rs200243273 | C | 0.27 (0.17-0.43) | $3.55 \times 10^{-8}$ | | 0.294 |
| | | | | | | | | | | | *CTD-224506.1* | rs4963073 | G | 3.23 (2.12-4.921) | $4.91 \times 10^{-8}$ | | 0.386 |
| | | | | | | | | | | | *CTD-224506.2* | rs4963072 | G | 3.34 (2.19-5.11) | $2.63 \times 10^{-8}$ | | 0.387 |
| | | | | | | | | | | | - | rs71469892 | G | 0.22 (0.15-0.31) | $2.15 \times 10^{-16}$ | | 0.232 |
| | | | | | | | | | | | - | rs145898170 | G | 0.47 (0.36-0.62) | $4.71 \times 10^{-8}$ | | - |
| | | | | | | | | | | | - | rs199838022 | C | 0.45 (0.34-0.57) | $7.14 \times 10^{-9}$ | | - |
| | | | | | | | | | | | - | rs12586854 | T | 0.18 (0.12-0.26) | $6.81 \times 10^{-19}$ | | 0.455 |
| | | | | | | | | | | | - | rs11157543 | C | 0.13 (0.07-0.22) | $5.97 \times 10^{-14}$ | | 0.344 |
| | | | | | | | | | | | - | rs11157544 | C | 0.30 (0.21-0.41) | $1.37 \times 10^{-13}$ | | 0.436 |
| | | | | | | | | | | | - | rs12586856 | G | 0.18 (0.11-0.29) | $3.77 \times 10^{-13}$ | | 0.417 |
| | | | | | | | | | | | - | rs11157545 | T | 0.44 (0.34-0.59) | $7.81 \times 10^{-9}$ | | 0.522 |
| | | | | | | | | | | | - | rs183643415 | A | 0.04 (0.01-0.12) | $2.77 \times 10^{-8}$ | | 0.108 |
| | | | | | | | | | | | - | rs150322840 | T | 0.13 (0.07-0.24) | $3.90 \times 10^{-10}$ | | 0.151 |
| | | | | | | | | | | | - | rs8005465 | A | 0.37 (0.27-0.51) | $4.41 \times 10^{-10}$ | | 0.472 |

| | | | | | | | | | | | | | | | | | |
|---|---|---|---|---|---|---|---|---|---|---|---|---|---|---|---|---|---|
| | | | | | | | | | | | - | rs543453148 | A | 25.22 (8.29-76.73) | 1.30 ×$10^{-8}$ | | 0.001 |
| | | | | | | | | | | | - | rs12890180 | G | 0.39 (0.28-0.53) | 2.91 ×$10^{-9}$ | | 0.456 |
| | | | | | | | | | | | - | rs73251857 | G | 0.06 (0.02-0.14) | 9.59 ×$10^{-10}$ | | 0.082 |
| | | | | | | | | | | | - | rs7141653 | C | 0.25 (0.17-0.37) | 1.65 ×$10^{-11}$ | | 0.317 |
| | | | | | | | | | | | - | rs7145329 | T | 0.34 (0.24-0.49) | 2.59 ×$10^{-9}$ | | 0.332 |
| | | | | | | | | | | | - | rs4900770 | A | 0.38 (0.28-0.53) | 9.10 ×$10^{-9}$ | | 0.452 |
| | | | | | | | | | | | - | rs58731325 | G | 0.34 0.25-0.47) | 7.35 ×$10^{-11}$ | | 0.495 |
| | | | | | | | | | | | *RP11-7-7O23.1* | rs115811519 | C | 4.93 (2.83-8.58) | 1.68 ×$10^{-8}$ | | 0.191 |
| | | | | | | | | | | | *CTC-501O10.1* | rs56383763 | C | 0.07 (0.03-0.16) | 1.75 ×$10^{-9}$ | | 0.086 |
| | | | | | | | | | | | *CRHR1* | rs373417 | T | 0.10 (0.05-0.20) | 1.24 ×$10^{-10}$ | | 0.086 |
| | | | | | | | | | | | | rs7221124 | A | 0.04 (0.02-0.09) | 6.70 ×$10^{-14}$ | | 0.359 |
| | | | | | | | | | | | | rs55938136 | A | 151.90 (62.14-371.50) | 3.37 ×$10^{-28}$ | | 0.309 |
| | | | | | | | | | | | *KANSL1* | rs11870844 | A | 3.98 (2.82-5.62) | 3.85 ×$10^{-15}$ | | 0.333 |
| | | | | | | | | | | | | rs371996525 | A | 0.04 (0.02-0.11) | 2.17 ×$10^{-10}$ | | 0.135 |
| | | | | | | | | | | | | rs142920272 | C | 0.10 (0.05-0.21) | 7.45 ×$10^{-11}$ | | 0.185 |
| | | | | | | | | | | | *KANSL1/ LRRC37A* | rs2668637 | G | 5.32 (3.09-9.13) | 1.43 ×$10^{-9}$ | | 0.037 |
| | | | | | | | | | | | | rs2696618 | C | 6.74 (4.02-11.31) | 5.09 ×$10^{-13}$ | | 0.173 |
| | | European | ATS/ERS criteria 2002 and ATS/ERS/JRS/ALAT guidelines 2011 | 3968 (samples from Allen et al., 2017) | 602 | 426/176 | 70.0±8.4 | 3366 | 2356.0/1010.0 | 65±5.5 | *MUC5AC* | rs34474233 | A | 3.15 (2.37-4.21) | 4.10 ×$10^{-14}$ | Affymetrix Axiom UK BiLEVE array (Affymetrix) and Affymetrix Axiom UK Biobank array (Affymetrix) | 0.022 |
| | | | | | | | | | | | | rs34815853 | A | 3.16 (2.37-4.20) | 4.13 ×$10^{-14}$ | | 0.021 |
| | | | | | | | | | | | *MUC5B* | rs12802931 | G | 2.42 (2.02-2.90) | 6.07 ×$10^{-22}$ | | 0.073 |
| | | | | | | | | | | | | rs35705950 | T | 4.11 (3.31-5.11) | 1.86 ×$10^{-37}$ | | 0.046 |
| | | | | | | | | | | | *CTD-224506.1* | rs4963072 | G | 1.29 (1.08-1.54) | 5.30 ×$10^{-3}$ | | 0.387 |

| | | | | | | | | | | | | | | | | | |
|---|---|---|---|---|---|---|---|---|---|---|---|---|---|---|---|---|---|
| | | | | | | | | | | | *CTC-501O10.1* | rs56383763 | C | 0.82 (0.68-0.98) | $2.42 \times 10^{-2}$ | | 0.086 |
| | | | | | | | | | | | *CRHR1* | rs373417 | T | 0.82 (0.69-0.98) | $2.72 \times 10^{-2}$ | | 0.086 |
| | | | | | | | | | | | *KANSL1* | rs371996525 | A | 0.80 (0.67-0.95) | $1.26 \times 10^{-2}$ | | 0.135 |
| | | | | | | | | | | | | rs142920272 | C | 0.83 (0.69-0.98) | $3.07 \times 10^{-2}$ | | 0.185 |
| | | | | | | | | | | | *KANSL1/LRRC37A* | rs2696618 | C | 1.25 (1.05-1.49) | $1.05 \times 10^{-2}$ | | 0.173 |
| 10 | Dressen et al. (2018)[10] | Caucasian | ATS/ERS/JRS/ALAT guidelines 2011 | 3384 | 1510 | 1119/391 | 67.3±7.9 | 1874 | 507/1367 | 56.38±9.32 | *MUC5B* | rs35705953 | T | 4.98 (4.40-5.65) | <0.0001 | Illumina X10 sequence | 0.046 |
| | | | | | | | | | | | *TERT* | rs2736100 | C | 0.75 (0.68-0.83) | <0.0001 | | 0.515 |
| 11 | Daniil et al. (2018)[11] | European | ATS/ERS/JRS/ALAT guidelines 2018 | 244 | 40 | 34/6 | 68.8±8.3 | 204 | - | - | *tRNA* | rs28358279* | C | 18.96 (5.66-63.52)~ | <0.001 | PCR | 0.102 “ |
| | | | | | | | | | | | | rs41456348* | C | 14.51 (4.21-50.01)~ | <0.001 | | 0.022 “ |
| | | | | | | | | | | | | rs527236198* | A | 106.96 (6.02-1898.22)~ | <0.001 | | 0.078 “ |
| | | | | | | | | | | | | rs1556423293*/A7472T | T | 14.43 (2.69-77.31)~ | 0.002 | | - |
| | | | | | | | | | | | *ND3* | rs2853826* | G | 3.20 (1.26-8.16)~ | <0.001 | | 0.169 “ |
| | | | | | | | | | | | | rs28358278* | T | 50.75 (6.14-419.45)~ | <0.001 | | 0.028 “ |
| | | | | | | | | | | | *16S rRNA* | C3157T | T | 91.56 (5.10-1640.98)~ | <0.001 | | - |
| | | | | | | | | | | | *ND2* | rs3020602* | C | 63.36 (3.42-1171.32)~ | <0.001 | | 0.007 “ |
| | | | | | | | | | | | *CYB* | G15884C | C | 11.22 (1.98-63.55)~ | 0.007 | | - |
| | | | | | | | | | | | *COII* | G8251A | A | 5.68 (1.56-20.66)~ | 0.012 | | - |
| | | | | | | | | | | | *16S rRNA* | A3136T | T | 38.17 (1.93-754.28)~ | 0.004 | | - |
| | | | | | | | | | | | *CYB* | rs1603225535* | T | 26.55 (1.25-564.09)~ | 0.026 | | 0.0002 “ |
| 12 | Allen et al. (2017)[G12] | European | ATS/ERS guidelines 2002 and ATS/ERS | 3968[D] | 602 | 426/176 | 70.0±8.4 | 3,366 | 2356/1010 | 65.0±5.5 | *CD1C* | rs147562345 | T | 4.45 (2.47-8.05) | $2.90 \times 10^{-6}$ | Affymetrix Axiom UK BiLEVE array (cases); Affymetrix | 0.003 |
| | | | | | | | | | | | *CFH* | rs560105094*/chr1:196625450 | G | 3.27 (2.12-5.04) | $5.09 \times 10^{-7}$ | | 0.0005(gnomAD) |

| | | | | | | | | | | | | | | | | | |
|---|---|---|---|---|---|---|---|---|---|---|---|---|---|---|---|---|---|
| | | | /JRS/ALAT guidelines 2011 | | | | | | | | *SRD5A2* | rs191031841 | T | 3.07 (2.21-4.27) | $1.80 \times 10^{-10}$ | Axiom UK BiLEVE array and Affymetrix Axiom UK Biobank array (controls) | 0.007 |
| | | | | | | | | | | | *TTC27* | rs6726541 | G | 3.78 (2.62-5.46) | $1.42 \times 10^{-12}$ | | 0.389 |
| | | | | | | | | | | | - | rs74266324 | A | 0.45 (0.34-0.59) | $4.56 \times 10^{-9}$ | | 0.284 |
| | | | | | | | | | | | - | rs33938858 | C | 0.20 (0.10-0.39) | $3.61 \times 10^{-9}$ | | 0.326 |
| | | | | | | | | | | | - | rs555288190*/chr2:138817685 | G | 47.8 (6.90-332) | $1.65 \times 10^{-6}$ | | 0.0005 (gnomAD) |
| | | | | | | | | | | | *FAM47E* | rs34369701 | A | 0.29 (0.16-0.54) | $3.73 \times 10^{-6}$ | | 0.199 |
| | | | | | | | | | | | - | rs28673968 | C | 0.47 (0.38-0.58) | $2.04 \times 10^{-12}$ | | 0.472 |
| | | | | | | | | | | | - | rs112863361 | A | 0.022 (0.001-0.36) | $1.60 \times 10^{-9}$ | | 0.031 |
| | | | | | | | | | | | *NDUFAF2* | rs140507420 | G | 2.98 (1.98-4.48) | $5.64 \times 10^{-7}$ | | 0.007 |
| | | | | | | | | | | | - | rs150626307 | A | 154 (21.0-1136) | $2.81 \times 10^{-7}$ | | 0.000 |
| | | | | | | | | | | | *DSP* | rs2076295 | G | 1.67 (1.44-1.92) | $4.14 \times 10^{-12}$ | | 0.432 |
| | | | | | | | | | | | *C6orf10* | rs9268159 | T | 0.43 (0.32-0.58) | $2.19 \times 10^{-8}$ | | 0.185 |
| | | | | | | | | | | | - | rs208501 | T | 1.83 (1.51-2.22) | $8.45 \times 10^{-10}$ | | 0.183 |
| | | | | | | | | | | | *FYN* | rs1409839 | C | 0.53 (0.42-0.67) | $1.10 \times 10^{-7}$ | | 0.365 |
| | | | | | | | | | | | *MRPL13* | rs562453723*/chr8:121446432 | T | 18.10 (6.07-53.9) | $6.69 \times 10^{-7}$ | | 0.000 |
| | | | | | | | | | | | - | rs10117639 | C | 25.30 (6.47-98.8) | $4.01 \times 10^{-6}$ | | 0.112 |
| | | | | | | | | | | | *MOB3B* | rs118048878 | A | 0.03 (0.002-0.45) | $1.50 \times 10^{-7}$ | | 0.007 |
| | | | | | | | | | | | *MOB3B* | rs1975503 | C | 0.15 (0.05-0.44) | $1.27 \times 10^{-6}$ | | 0.117 |
| | | | | | | | | | | | *REEP3* | chr10:65359362 | T | 36.40 (7.75-172) | $1.16 \times 10^{-6}$ | | - |
| | | | | | | | | | | | - | rs776241108*/chr10:77228877 | A | 21.10 (6.25-70.9) | $7.53 \times 10^{-7}$ | | 0.001 (gnomAD) |

| | | | | | | | | | | | | | | | | | |
|---|---|---|---|---|---|---|---|---|---|---|---|---|---|---|---|---|---|
| | | | | | | | | | | | - | rs746131658*/chr10:107751048 | C | 27.90 (7.21-108) | 6.44 $\times 10^{-7}$ | | 0.000 (ALSPAC) |
| | | | | | | | | | | | *MUC5B* | rs35705950 | T | 4.11 (3.31-5.11) | 1.86 $\times 10^{-37}$ | | 0.046 |
| | | | | | | | | | | | *HSD17B12* | rs1305768509*/chr11:43823527 | A | 1.47 (1.25-1.72) | 2.53 $\times 10^{-6}$ | | 0.000 (TOPMED) |
| | | | | | | | | | | | - | rs181297970 | T | 6.18 (3.17-12.0) | 3.27 $\times 10^{-7}$ | | 0.004 |
| | | | | | | | | | | | - | rs538043452*/chr12:38945784 | T | 3.84 (2.44-6.03) | 2.08 $\times 10^{-8}$ | | 0.002 |
| | | | | | | | | | | | *LINC00327* | rs186638373 | C | 131 (14.4-1192) | 5.52 $\times 10^{-7}$ | | 0.001 |
| | | | | | | | | | | | - | rs193268061 | C | 49.30 (7.76-313) | 3.46 $\times 10^{-6}$ | | 0.000 |
| | | | | | | | | | | | - | rs76271340 | T | 0.48 (0.37-0.61) | 1.15 $\times 10^{-8}$ | | 0.012 |
| | | | | | | | | | | | *AKAP13/ KLHL25* | rs62025270 | A | 1.49 (1.26-1.76) | 3.11 $\times 10^{-6}$ | | 0.113 |
| | | | | | | | | | | | *IL9RP3* | rs367849850 | G | 2.38 (1.68-3.35) | 1.82 $\times 10^{-6}$ | | 0.014 |
| | | | | | | | | | | | *GOSR2* | rs117791180 | T | 2.81 (1.96-4.01) | 7.87 $\times 10^{-8}$ | | 0.005 |
| | | | | | | | | | | | *APOC1P1* | rs60049679 | C | 0.35 (0.21-0.58) | 2.47 $\times 10^{-6}$ | | 0.135 |
| | | | | | | | | | | | *TTYH1* | rs73606754 | G | 0.46 (0.38-0.57) | 2.24 $\times 10^{-13}$ | | 0.070 |
| | | | | | | | | | | | *LILRP2* | rs141247056 | A | 3.17 (2.35-4.28) | 4.49 $\times 10^{-14}$ | | 0.033 |
| | | | | | | | | | | | *RDH13* | rs552589281*/chr19:55557609 | A | 63.00 (15.4-257) | 2.25 $\times 10^{-11}$ | | 0.003 |
| | | | | | | | | | | | *RDH13* | rs28591280 | C | 0.47 (0.36-0.60) | 2.68 $\times 10^{-9}$ | | 0.199 |
| | | | | | | | | | | | *SULT4A1* | rs552369499*/chr22:44250669 | T | 55.60 (9.50-326) | 1.28 $\times 10^{-6}$ | | 0.001 |
| | | American-European $^{\alpha}$ and non- | (ATS/ERS guidelines 2000 and | 7353 $^{R}$ | 2158 [542 $^{\alpha}$, | 1475/683 [(385/15 | (68.0±3.0) $^{\alpha}$, (65.5±9.5) $^{\beta}$ | 5195 [512$^{\alpha}$, | 2,531/2,664 [(242/270) | (63.0±7.5) $^{\alpha}$ | *DSP* | rs2076295 | G | 1.39 (1.29-1.50) | 2.47 $\times 10^{-18}$ | Affymetrix Genome-Wide Human SNP 6.0 Array$^{\alpha}$; | 0.432 |

| | | | | | | | | | | | | | | | | | |
|---|---|---|---|---|---|---|---|---|---|---|---|---|---|---|---|---|---|
| | | Hispanic white[β] | ATS/ERS/JRS/ALAT guidelines 2011)[α] and ATS/ERS guidelines 2002[β] | | 1616[β]] | 7)[α], (1090/526)[β]] | | 4683[β]] | [α], (2289/2394)[β]] | | *MUC5B* | rs35705950 | T | 2.46 (2.13-2.85) | $3.13 \times 10^{-34}$ | Illumina 660 Quad beadchip[β] | 0.046 |
| | | | | | | | | | | | *AKAP13/KLHL25* | rs62025270 | A | 1.22 (1.11-1.33) | $9.96 \times 10^{-6}$ | | 0.113 |
| | | European, American-European[α] and non-Hispanic white[β] | ATS/ERS guidelines 2000, ATS/ERS guidelines 2002 and ATS/ERS/JRS/ALAT guidelines 2011 | 11321[M] | 2760 [602, 542[α], 1616[β]] | 1,901/859 [(426/176), (385/157)[α], (1090/526)[β]] | 70.0±8.4, 68.0±3.0[α], 65.5±9.5[β] | 8,561 [3366, 512[α], 4683[β]] | 4,887/3674 [(2356/1010), (242/270)[α], (2289/2394)[β]] | 65.0±5.5, 63.0±7.5[α] | *DSP* | rs2076295 | G | 1.44 (1.35-1.54) | $7.81 \times 10^{-28}$ | Affymetrix Axiom UK BiLEVE array (discovery cases); Affymetrix Axiom UK BiLEVE array and Affymetrix Axiom UK Biobank array (discovery controls), Affymetrix Genome-Wide Human SNP 6.0 Array[α]; Illumina 660 Quad beadchip[β] | 0.432 |
| | | | | | | | | | | | *MUC5B* | rs35705950 | T | 2.89 (2.56-3.26) | $1.12 \times 10^{-66}$ | | 0.046 |
| | | | | | | | | | | | *AKAP13/KLHL25* | rs62025270 | A | 1.27 (1.18-1.37) | $1.32 \times 10^{-9}$ | | 0.113 |
| 13 | Petrovski et al. (2017)[13] | European | ATS/ERS/JRS/ALAT guidelines 2011 | 525 | 183 | - | - | 342 | - | - | *MUC5B* | rs35705950 | T | 4.0 (2.8–5.6) | $3.1 \times 10^{-17}$ | Illumina HiSeq 2000 or 2500 sequencer | 0.046 |
| 14 | Hirano et al. (2016)[14] | Japanese | ATS/ERS criteria 2002 | 375 | 65 | 55/10 | 65.8±9.9 | 310 | 255/55 | 50.6±7.8 | *FAM13A* | rs2609255 | G | 1.78 (1.29-2.44) | <0.001 | PCR | 0.348 |
| 15 | Yamaguchi et al. (2016)[15] | Japanese | ATS/ERS/JRS/ALAT guideline s 2011 | 390 | 87 | 76/11 | 67.5±9.9 | 303 | 248/55 | 50.5±7.8 | *AGER* | rs2070600 | CT+TT vs CC | 1.84 (1.08-3.1) | 0.021 | PCR | 0.072 |

| | | | | | | | | | | | | | | | | | |
|---|---|---|---|---|---|---|---|---|---|---|---|---|---|---|---|---|---|
| 16 | Kishore et al. (2016) [16] | European | ATS/ERS criteria 2000, ATS/ERS criteria 2001 and ATS/ERS/JRS/ALAT guidelines 2011 | 261 | 165 | 125/40 | 67.9±11.60 | 96 | 45/51 | 34.5±8.94 | *MUC5B* | rs35705950 | T | 5.23 (3.06-8.94) | $1.80 \times 10^{-11}$ | Sequenom MassARRAY platform integrating iPLEX® SBE reaction and MassARRAY® technology (Agena Bioscience, San Diego, CA, USA) based MALDI-TOF MS assay | 0.046 |
| | | | | | | | | | | | *IL-1α* | rs1800587 | T | 0.68 (0.99-0.46) | $4.94 \times 10^{-2}$ | | - |
| | | | | | | | | | | | *TF* | rs1799899 | A | 2.06 (1.12-3.78) | $1.87 \times 10^{-2}$ | | - |
| | | | | | | | | | | | *MUC2* | rs7934606 | A | 2.18 (1.50-3.16) | $3.73 \times 10^{-5}$ | | 0.380 “ |
| 17 | Van der Vis et al. (2015) [17] | Caucasian | ATS/ERS criteria 2002 and ATS/ERS criteria 2013 and ATS/ERS/JRS/ALAT guidelines 2011 | 364 | 115 | 97/18 | 63.5±11.0 | 249 | - | - | *MUC5B* | rs35705950 | T | 3.60 (2.37-5.55)~ | $5.0 \times 10^{-10}$ | TaqMan SNP genotyping assay (PCR) | 0.046 |
| 18 | Aquino-Gálvez et al. (2015) [18] | Mexican | ATS/ERS/JRS/ALAT guidelines 2011 | 373 | 168 | 103/65 | 64.5±11.0 | 205 | 36/169 | 47.0±5.4 | *HSPA1B* | rs1061581 | AA vs AG+GG | 0.27 (0.13-0.57) | 0.0003 | 5’ nuclease SNP genotyping assays (PCR) | - |
| | | | | | | | | | | | *HSPA1A* | rs1043618 | GG vs GC+CC | 0.63 (0.41–0.97) | 0.039 | | 0.481 |
| | | | | | | | | | | | *HSPA1L* | rs2227956 | G | 0.27 (0.10-0.75) | 0.01 | | 0.122 |
| | | | | | | | | | | | | rs2075800 | CC vs TT | 2.52 (1.32-4.81) | 0.005 | | 0.255 |
| 19 | Peljto et al. (2015) [19] | Mexican | ATS/ERS/JRS/ALAT guidelines 2011 | 194 | 83 | 59/24 | 66.0±7.7 | 111 | - | - | *LRRC34* | rs6793295 | C | 2.24 | 0.01 | - | 0.421 |
| | | | | | | | | | | | *FAM13A* | rs2609255 | G | 2.09 | 0.02 | | 0.348 |
| | | | | | | | | | | | *TERT* | rs2736100 | C | 0.50 | 0.05 | | 0.484 |
| | | | | | | | | | | | *MUC5B* | rs35705950 | T | 7.36 | $1.1 \times 10^{-4}$ | | 0.046 |

| | | | | | | | | | | | | | | | | | |
|---|---|---|---|---|---|---|---|---|---|---|---|---|---|---|---|---|---|
| | | | | | | | | | | | *TOLLIP* | rs111521887 | G | 3.79 | 0.01 | | - |
| | | | | | | | | | | | *IVD* | rs2034650 | G | 0.40 | 0.01 | | 0.491 |
| | | Korean | | 326 | 239 | 181/60 | 65.1±7.7 | 87 | - | - | *IVD* | rs2034650 | G | 0.13 | 8.1×10$^{-4}$ | | 0.491 |
| 20 | Zhang et al. (2015)[20] | Han Chinese | ATS/ERS/JRS/ALAT guidelines 2011 | 368 | 102 | 45/57 | 59.3±9.87 | 266 | 137/129 | 56.7±12.80 | *HLA-DRB1* | DRB1*08 | DRB1*08 | 0.43 (0.19-0.96) | 0.035 | High resolution melting (HRM) method | - |
| | | | | | | | | | | | *IL-10* | rs1800896 | G | 1.80 (0.99-3.26) | 0.05 | | 0.272 |
| 21 | Zhang et al. (2015)[21] | Chinese | ATS/ERS/JRS/ALAT guidelines 2013 | 215 | 115 | 67/48 | 58.1±11.4 | 100 | 55/45 | 57.2±5.5 | *MMP-9* | rs3918242 | T | 1.62 (1.040-2.536) | 0.034 | PCR-RFLP | 0.155 |
| 22 | Horimasu et al. (2015)[22] | Japanese | ATS/ERS criteria 2002 | 354 | 44 | 35/9 | 67.5±1.6 | 310 | 255/55 | 50.6±0.4 | *MUC5B* | rs35705950 | T | 4.34 (1.02-18.49) | 0.031 | PCR | 0.046 |
| | | German | | 106 | 71 | 51/20 | 67.6±1.2 | 35 | 15/20 | 44.3±2.3 | *MUC5B* | rs35705950 | T | 11.05 (3.30-36.99) | <0.001 | | 0.046 |
| 23 | Mathai et al. (2015)[23] | American`` | ATS/ERS/JRS/ALAT guidelines 2011 | 458 | 230 | 159/71 | 65.4 (64.2-66.5) | 228 | 151/79 | 64.5 (63.7-65.4) | *DSP* | rs72825047 | G | 0.4 (0.19-0.85)~ | 0.0007 | Illumina HiSeq platform | 0.037 |
| | | | | | | | | | | | | rs72825054 | C | 0.4 (0.19-0.85)~ | 0.0003 | | 0.037 |
| | | | | | | | | | | | | rs72825058 | T | 0.40 (0.17-0.78)~ | 0.0005 | | 0.038 |
| | | | | | | | | | | | | rs874646 | G | 0.60 (0.34-0.89)~ | 0.002 | | 0.256 |
| | | | | | | | | | | | | rs2744362 | C | 0.60 (0.34-0.89)~ | 0.002 | | 0.256 |
| | | | | | | | | | | | | rs2008748 | T | 0.60 (0.34-0.89)~ | 0.001 | | 0.745 |
| | | | | | | | | | | | | rs2744365 | G | 0.60 (0.34-0.89)~ | 0.002 | | 0.230 |
| | | | | | | | | | | | | rs2237102 | C | 0.60 (0.34-0.89)~ | 0.002 | | 0.245 |
| | | | | | | | | | | | | rs2237103 | G | 0.60 (0.34-0.89)~ | 0.002 | | 0.250 |
| | | | | | | | | | | | | rs2237104 | T | 0.60 (0.34-0.89)~ | 0.002 | | 0.246 |

| | | | | | | | | | | | | | | | | | |
|---|---|---|---|---|---|---|---|---|---|---|---|---|---|---|---|---|---|
| | | | | | | | | | | | | rs2078438 | A | 0.60 (0.34-0.89)~ | 0.002 | | 0.230 |
| | | | | | | | | | | | | rs2744368 | T | 0.60 (0.34-0.89)~ | 0.002 | | 0.233 |
| | | | | | | | | | | | | rs2744369 | T | 0.60 (0.34-0.89)~ | 0.001 | | 0.234 |
| | | | | | | | | | | | | rs2744370 | C | 0.60 (0.34-0.89)~ | 0.002 | | 0.236 |
| | | | | | | | | | | | | rs2744371 | C | 0.60 (0.34-0.89)~ | 0.002 | | 0.234 |
| | | | | | | | | | | | | rs2744372 | G | 0.60 (0.34-0.89)~ | 0.002 | | 0.249 |
| | | | | | | | | | | | | rs2744374 | T | 0.60 (0.34-0.89)~ | 0.002 | | 0.250 |
| | | | | | | | | | | | | rs2744373 | A | 0.50 (0.34-1.04)~ | 0.004 | | 0.108 |
| | | | | | | | | | | | | rs2744375 | T | 0.50 (0.34-1.04)~ | 0.004 | | 0.146 |
| | | | | | | | | | | | | rs17133512 | A (insertion ) | 1.70 (1.05-2.96)~ | 0.004 | | 0.199 |
| | | | | | | | | | | | | rs145125286 | C (deletion) | 0.50 (0.34-1.04)~ | 0.003 | | 0.116 |
| | | | | | | | | | | | | rs2008756 | G | 0.60 (0.34-0.89)~ | 0.0009 | | 0.246 |
| | | | | | | | | | | | | rs2237105 | A | 0.60 (0.34-0.89)~ | 0.002 | | 0.230 |
| | | | | 1872 | 936 | 647/289 | 64.9 (64.34-65.60) | 936 | 653/283 | 62.7 (62.23-63.24) | *DSP* | rs2744371 | C | 0.77 (0.66-0.91) | 0.002 | TaqMan genotyping assays | 0.234 |
| | | | | | | | | | | | | rs2744373 | A | 0.80 (0.66-0.98) | 0.03 | | 0.108 |
| | | | | | | | | | | | | rs2076295/6p24 | G | 1.36 (1.19-1.56) | <0.001 | | 0.432 |
| 24 | Jiang et al. (2015) [24] | Han Chinese | ATS/ERS criteria 2002 | 437 | 187 | 138/49 | 69.7±4.3 | 250 | 172/78 | 67.7±7.3 | *MUC5B* | rs35705950 | T | 1.93 (1.32-2.82) | 0.0007 | TaqMan probe-based SNP genotyping | 0.046 |
| 25 | Wang et al. (2014) [25] | Chinese | ATS/ERS/JRS/ALAT guidelines 2011 | 1178 | 165 | 101/64 | 61.8±12.72 | 1013 | 525/488 | 58.6±12.72 | *MUC5B* | rs35705950 | T | 4.33 (1.99-9.42) | 0.001 | PCR-RFLP and TaqMan probe-based SNP genotyping | 0.046 |
| | | | | 485 | 80 (male and | - | - | 405 (male and | - | - | *MUC5B* | rs35705950 | T | 10.48 (2.60-42.36) | 0.001 | | 0.046 |

| | | | | | | | | | | | | | | | | | |
|---|---|---|---|---|---|---|---|---|---|---|---|---|---|---|---|---|---|
| | | | | | >=55 y) | | | >=55 y) | | | | | | | | | |
| 26 | Uh et al. (2014)[26] | Korean | ATS/ERS guidelines 2002 | 420 | 237 | 163/74 | 62.0(31-83) | 183 | 55/128 | 61.0(50-81) | *ADAM33* | rs628977 | AA vs AG+GG | 0.50 (0.27-0.94) | 0.03 | Single-Base Extension (SBE) | 0.341 |
| | | | | 345 | 162 surgical IPF | 112/50 | 59.0(31-83) | 183 | 55/128 | 61.0(50-81) | *ADAM33* | rs628977 | AA vs AG+GG | 0.40 (0.19-0.84) | 0.02 | | 0.341 |
| 27 | Sangiuolo et al. (2014)[27] | Italian | ATS/ERS/JRS/ALAT guidelines 2011 | 196 | 89 | 67/22 | 68.8±7.6 | 107 | 105/2 | 54.3±4.3 | *HFE* | rs1799945* | GC | 2.60 (1.35-5.02) | 0.004** | reverse dot blot | 0.073 |
| 28 | Coghlan et al.(2014)[28] | Mixed, European (90% Caucasian, 5% African-American, 3% Asian and 2% Hispanic) | ATS/ERS/JRS/ALAT guidelines 2011 | 324 | 132 | 98/34 | 54.6±9.2 | 192 | 99/93 | 60.2±8.1 | *MUC5B* | rs35705950 | T | 2.67 (1.78-4.01) | <0.0001 | NGS | 0.046 |
| 29 | Ohshimo et al. (2014)[29] | German | - | 127 | 42 | - | - | 85 | - | - | *ANGPT2* | rs2515475 | T | - | 0.048 | RT-PCR | 0.0132 |
| 30 | Fingerlin et al. (2013)[G 30] | non-hispanic white | ATS/ERS guidelines 2002 | 6299[D] | 1616 | 1090/526 | 65.5±9.5 | 4683 | 2289/2394 | - | *TERT* | rs2736100 | C | 0.73 (0.67–0.79) | $7.60 \times 10^{-14}$ | Illumina Human 660W Quad BeadChip | 0.484 |
| | | | | | | | | | | | *DSP* | rs2076295 | G | 1.43 (1.32–1.55) | $1.14 \times 10^{-16}$ | | 0.432 |
| | | | | | | | | | | | - | rs4727443 | A | 1.30 (1.20–1.41) | $6.72 \times 10^{-9}$ | | 0.381 |
| | | | | | | | | | | | *MUC2* | rs7934606 | C | 1.52 (1.40–1.65) | $5.46 \times 10^{-22}$ | | 0.380 “ |
| | | | | | | | | | | | - | rs2034650 | G | 0.77 (0.71–0.84) | $1.86 \times 10^{-9}$ | | 0.491 |
| | | | | | | | | | | | *MAPT* | rs1981997 | A | 0.71 (0.64-0.78) | $2.52 \times 10^{-8}$ | | 0.087 |
| | | | | | | | | | | | *DPP9* | rs12610495 | G | 1.29 (1.18–1.41) | $9.57 \times 10^{-9}$ | | 0.183 |
| | | | | | | | | | | | - | rs3778337 | A | 1.31(1.20-1.43) | $6.41 \times 10^{-9}$ | | 0.303 |
| | | | | | | | | | | | - | rs1379326 | C | 1.78 (1.45-2.19) | $5.74 \times 10^{-9}$ | | 0.329 |

| | | | | | | | | | | | | | | | | | |
|---|---|---|---|---|---|---|---|---|---|---|---|---|---|---|---|---|---|
| | | | | | | | | | | | - | rs7942850 | C | 1.38 (1.27-1.50) | $9.29 \times 10^{-14}$ | | 0.315 |
| | | | | | | | | | | | - | rs6421972 | T | 1.51 (1.39-1.64) | $1.62 \times 10^{-21}$ | | 0.390 |
| | | | | | | | | | | | - | rs7480563 | C | 0.69 (0.64-0.75) | $4.17 \times 10^{-18}$ | | 0.697 |
| | | | | | | | | | | | - | rs4077759 | C | 0.74 (0.67-0.80) | $8.47 \times 10^{-13}$ | | 0.484 |
| | | | | | | | | | | | - | rs868903 | T | 0.64 (0.59-0.70) | $1.26 \times 10^{-22}$ | | 0.510 |
| | | | | | | | | | | | - | rs2334659 | A | 0.71 (0.63-0.80) | $4.71 \times 10^{-9}$ | | 0.061 |
| | | | | | | | | | | | - | rs7122936 | C | 0.76 (0.69-0.82) | $3.69 \times 10^{-8}$ | | 0.346 |
| | | | | | | | | | | | - | rs1992272 | T | 0.78 (0.71-0.85) | $3.49 \times 10^{-8}$ | | 0.271 |
| | | | | | | | | | | | - | rs17563986 | G | 0.71 (0.64-0.78) | $3.39 \times 10^{-8}$ | | 0.086 |
| | | | | | | | | | | | - | rs807023 | G | 0.71 (0.64-0.79) | $1.22 \times 10^{-8}$ | | 0.773 |
| | | | | | | | | | | | - | rs2109069 | A | 1.28 (1.18-1.40) | $3.87 \times 10^{-8}$ | | 0.211 |
| | | | | 2766 R | 876 | 599/277 | 63.4±9.7 | 1890 | 907/983 | 58.4±9.7 | *TERT* | rs2736100 | C | 0.74 (0.65–0.83) | $4.05 \times 10^{-7}$ | Multiplex (Sequenom) iPLEX and uniplex (TaqMan) assays | 0.484 |
| | | | | | | | | | | | *DSP* | rs2076295 | T | 1.26 (1.13–1.42) | $6.28 \times 10^{-5}$ | | 0.432 |
| | | | | | | | | | | | *MUC2* | rs7934606 | C | 1.56 (1.39–1.76) | $1.49 \times 10^{-13}$ | | 0.380 “ |
| | | | | | | | | | | | - | rs2034650 | G | 0.82 (0.74–0.93) | 0.00098 | | 0.491 |
| | | | | | | | | | | | *MAPT* | rs1981997 | A | 0.67 (0.58-0.79) | $4.74 \times 10^{-7}$ | | 0.087 |
| | | | | | | | | | | | *DPP9* | rs12610495 | G | 1.30 (1.15–1.47) | $3.94 \times 10^{-5}$ | | 0.183 |
| | | | | 9065 M | 2492 | 1689/803 | - | 6573 | 3196/3377 | - | *TERT* | rs2736100 | C | - | $1.71 \times 10^{-19}$ | | 0.503 |
| | | | | | | | | | | | *DSP* | rs2076295 | T | - | $1.08 \times 10^{-19}$ | | 0.432 |
| | | | | | | | | | | | - | rs4727443 | A | - | $1.17 \times 10^{-8}$ | | 0.381 |
| | | | | | | | | | | | *MUC2* | rs7934606 | C | - | $6.87 \times 10^{-34}$ | | 0.38 “ |
| | | | | | | | | | | | - | rs2034650 | G | - | $9.76 \times 10^{-12}$ | | 0.491 |
| | | | | | | | | | | | *MAPT* | rs1981997 | A | - | $8.87 \times 10^{-14}$ | | 0.087 |
| | | | | | | | | | | | *DPP9* | rs12610495 | G | - | $1.68 \times 10^{-12}$ | | 0.183 |

| | | | | | | | | | | | | | | | | | |
|---|---|---|---|---|---|---|---|---|---|---|---|---|---|---|---|---|---|
| | | | | | | | | | | | *LRRC34* | rs6793295 | C | - | $8.33 \times 10^{-13}$ | | 0.421 |
| | | | | | | | | | | | *FAM13A* | rs2609255 | G | - | $2.20 \times 10^{-11}$ | | 0.348 |
| | | | | | | | | | | | *OBFC1* | rs11191865 | A | - | $2.44 \times 10^{-8}$ | | 0.397 |
| | | | | | | | | | | | *ATP11A* | rs1278769 | A | - | $6.72 \times 10^{-9}$ | | 0.233 |
| | | | | 3625 | 1735 | - | - | 1890 | - | - | *MUC5B* | rs35705950 | T | 4.51 (3.91-5.21) | $7.21 \times 10^{-95}$ | | 0.046 |
| 31 | Noth et al. (2013)[G31] | European-American (617) and non-European-American (16) | ATS/ERS guidelines 2000 and ATS/ERS/JRS/ALAT guidelines 2011 | 1084[D] | 542 | 385/157 | 67.0(61-73) | 542 | - | 63.0(40-70) | *SLC35F3* | rs11810452 | G | - | $6.31 \times 10^{-6}$ | Genome-Wide Human SNP 6.0 Array | 0.230 |
| | | | | | | | | | | | | rs12750467 | A | 1.55 (1.25-1.93) | $6.31 \times 10^{-5}$ | | 0.071 |
| | | | | | | | | | | | *ARHGEF4* | rs13034562 | G | - | $6.31 \times 10^{-5}$ | | 0.555 |
| | | | | | | | | | | | | rs66525751 | A | 1.79 (1.36-2.35) | $7.94 \times 10^{-6}$ | | 0.064 |
| | | | | | | | | | | | *ABCG2* | chr4:89147344 | - | 7.36 (2.37-22.9) | $1.58 \times 10^{-6}$ | | - |
| | | | | | | | | | | | - | rs10065111 | T | 1.57 (1.28-1.93) | $1.58 \times 10^{-5}$ | | 0.196 |
| | | | | | | | | | | | - | rs7449198 | A | 1.65 (1.33-2.04) | $1.26 \times 10^{-6}$ | | 0.132 |
| | | | | | | | | | | | - | rs58608739 | A | 0.32 (0.18-0.56) | $6.31 \times 10^{-6}$ | | 0.144 |
| | | | | | | | | | | | - | rs6862667 | G | 0.32 (0.17-0.59) | $7.94 \times 10^{-5}$ | | 0.208 |
| | | | | | | | | | | | *MAD1L1* | rs55948146 | G | 0.66 (0.79-0.55) | $2.00 \times 10^{-6}$ | | 0.437 |
| | | | | | | | | | | | | rs6977733 | A | 0.67 (0.56-0.80) | $1.00 \times 10^{-5}$ | | 0.315 |
| | | | | | | | | | | | | rs3800917 | A | 0.67 (0.56-0.08) | $7.94 \times 10^{-6}$ | | 0.302 |
| | | | | | | | | | | | *K03193* | rs76795398 | G | 2.36 (1.54-3.63) | $1.00 \times 10^{-5}$ | | 0.014 |
| | | | | | | | | | | | | rs79842896 | C | 2.49 (1.61-3.85) | $7.94 \times 10^{-6}$ | | 0.014 |
| | | | | | | | | | | | - | rs113172295 | T | - | $7.94 \times 10^{-6}$ | | 0.054 |
| | | | | | | | | | | | *COLIA2* | rs2293739 | A | 0.63 (0.48-0.82) | $3.98 \times 10^{-4}$ | | 0.063 |
| | | | | | | | | | | | *MYOM2* | rs17751471 | A | 1.56 (1.24-1.97) | $2.51 \times 10^{-5}$ | | 0.105 |
| | | | | | | | | | | | | rs6558599 | G | 1.87 (2.48-1.41) | $7.94 \times 10^{-6}$ | | 0.112 “ |

| | | | | | | | | | | | | | | | | | |
|---|---|---|---|---|---|---|---|---|---|---|---|---|---|---|---|---|---|
| | | | | | | | | | | | *PPP2R2A* | rs1425735 | A | 1.45 (1.76-1.20) | $1.00 \times 10^{-4}$ | | 0.470 |
| | | | | | | | | | | | *LRP12* | rs4291236 | C | 1.78 (2.45-1.30) | $1.58 \times 10^{-4}$ | | 0.029 |
| | | | | | | | | | | | | rs4537272 | C | - | $2.00 \times 10^{-6}$ | | 0.017 |
| | | | | | | | | | | | - | rs138020443*/ chr9:25356216 | T | 10.27 (2.24-46.6) | $7.94 \times 10^{-6}$ | | 0.003 |
| | | | | | | | | | | | - | rs151045778*/ chr9:25370576 | T | - | $1.26 \times 10^{-5}$ | | 0.004 |
| | | | | | | | | | | | *MUC5B* | rs35705950 | T | 1.57 (1.20-2.05) | $6.31 \times 10^{-6}$ | | 0.046 |
| | | | | | | | | | | | - | rs11041133 | G | 0.81 (0.96-0.68) | $1.00 \times 10^{-2}$ | | 0.524 |
| | | | | | | | | | | | - | rs7481967 | C | 0.98 (1.25-0.77) | $7.94 \times 10^{-1}$ | | 0.324 |
| | | | | | | | | | | | *TOLLIP* | rs5744033 | G | - | $1.26 \times 10^{-6}$ | | 0.927 |
| | | | | | | | | | | | | rs3168046 | C | 1.24 (1.05-1.47) | $1.00 \times 10^{-2}$ | | 0.563 |
| | | | | | | | | | | | | rs3829223 | C | 0.83 (0.99-0.70) | $2.51 \times 10^{-2}$ | | 0.501 |
| | | | | | | | | | | | | rs3750920 | T | 1.25 (1.06-1.48) | $5.01 \times 10^{-3}$ | | 0.360 |
| | | | | | | | | | | | | rs5743961 | A | 0.82 (0.55-1.21) | $2.51 \times 10^{-1}$ | | 0.048 |
| | | | | | | | | | | | | rs111521887 | G | 1.54 (1.26-1.87) | $5.01 \times 10^{-7}$ | | 0.072 |
| | | | | | | | | | | | | rs908225 | T | 0.75 (0.91-0.62) | $2.51 \times 10^{-3}$ | | 0.376 |
| | | | | | | | | | | | | rs5743944 | A | 0.69 (0.56-0.85) | $1.58 \times 10^{-4}$ | | 0.148 |
| | | | | | | | | | | | | rs5743942 | C | 0.88 (0.75-1.05) | $1.00 \times 10^{-1}$ | | 0.338 |
| | | | | | | | | | | | | rs5743905 | C | 0.82 (0.55-1.21) | $2.51 \times 10^{-1}$ | | 0.047 |
| | | | | | | | | | | | | rs5743900 | G | 0.69 (0.55-0.86) | $3.16 \times 10^{-4}$ | | 0.930 |
| | | | | | | | | | | | | rs5743899 | G | 0.90 (1.12-0.72) | $3.16 \times 10^{-1}$ | | 0.339 |
| | | | | | | | | | | | | rs5743894 | G | 1.58 (1.30-1.93) | $1.26 \times 10^{-7}$ | | 0.074 |

| | | | | | | | | | | | | | | | | | |
|---|---|---|---|---|---|---|---|---|---|---|---|---|---|---|---|---|---|
| | | | | | | | | | | | | rs117572864 | T | 1.11 (0.73-1.69) | $6.31 \times 10^{-1}$ | | 0.024 |
| | | | | | | | | | | | | rs5743890 | G | 0.68 (0.53-0.88) | $2.51 \times 10^{-3}$ | | 0.047 |
| | | | | | | | | | | | | rs4963062 | A | 1.09 (1.52-0.77) | $6.31 \times 10^{-1}$ | | 0.210 |
| | | | | | | | | | | | - | rs75735056 | A | - | $2.00 \times 10^{-8}$ | | 0.256 |
| | | | | | | | | | | | *MDGA2* | rs4898572 | A | 1.89 (2.52-1.42) | $1.00 \times 10^{-5}$ | | 0.035 |
| | | | | | | | | | | | | rs7144383 | G | 1.82 (1.37-2.41) | $2.00 \times 10^{-5}$ | | 0.038 |
| | | | | | | | | | | | - | rs573297762*/ chr15:40693273 | G | - | $2.51 \times 10^{-7}$ | | 0.000 |
| | | | | | | | | | | | *IVD* | rs1001528 | A | 0.65 (0.77-0.55) | $1.00 \times 10^{-6}$ | | 0.493 |
| | | | | | | | | | | | *IQCH* | rs17232873 | G | 2.16 (1.47-3.19) | $6.31 \times 10^{-5}$ | | 0.065 |
| | | | | | | | | | | | *MAP2K5* | rs12905544 | T | 2.53 (1.69-3.77) | $1.00 \times 10^{-6}$ | | 0.025 |
| | | | | | | | | | | | *SPPL2C, MAPT-AS1* | rs17690703 | T | 0.62 (0.51-0.76) | $5.01 \times 10^{-6}$ | | 0.123 |
| | | | | | | | | | | | - | rs78795069 | C | 0.52 (0.34-0.80) | $5.01 \times 10^{-4}$ | | 0.023 |
| | | | | | | | | | | | - | rs721597 | C | 1.44 (1.21-1.70) | $6.31 \times 10^{-6}$ | | 0.373 |
| | | European-American | | 1231[R] | 544 | 262/108 | 65.0(59-70) | 687 | 393/294 | 56.0(46-65) | *MUC5B* | rs35705950 | T | 3.72 (3.02-4.58) | $2.50 \times 10^{-40}$ | iPLEX Gold™ platform | 0.046 |
| | | | | | | | | | | | *TOLLIP* | rs111521887 | G | 1.58 (1.29-1.94) | $7.90 \times 10^{-6}$ | | 0.072 |
| | | | | | | | | | | | | rs5743894 | G | 1.52 (1.25-1.86) | $1.60 \times 10^{-5}$ | | 0.074 |
| | | | | | | | | | | | | rs5743890 | G | 0.57 (0.44-0.73) | $4.00 \times 10^{-6}$ | | 0.047 |
| | | | | | | | | | | | *MDGA2* | rs7144383 | G | 1.57 (1.18-2.08) | $1.00 \times 10^{-3}$ | | 0.038 |
| | | | | | | | | | | | *SPPL2C, MAPT-AS1* | rs17690703 | T | 0.74 (0.61-0.89) | $1.60 \times 10^{-3}$ | | 0.123 |
| | | European-American | | 1026[R] | 324 | 68/22 | 68.0(63-72) | 702 | 497/205 | 62.0(55-68) | *MUC5B* | rs35705950 | T | 3.08 (2.37-3.99) | $5.14 \times 10^{-16}$ | | 0.046 |
| | | | | | | | | | | | *TOLLIP* | rs111521887 | G | 1.24 (0.98-1.50) | $8.03 \times 10^{-2}$ | | 0.072 |

| | | | | | | | | | | | | | | | | | |
|---|---|---|---|---|---|---|---|---|---|---|---|---|---|---|---|---|---|
| | | | | | | | | | | | *TOLLIP* | rs5743894 | G | 1.19 (0.94-1.50) | $1·57 \times 10^{-1}$ | | 0.074 |
| | | | | | | | | | | | *TOLLIP* | rs5743890 | G | 0.60 (0.45-0.80) | $3·49 \times 10^{-4}$ | | 0.047 |
| | | | | | | | | | | | *SPPL2C* | rs17690703 | T | 0.78 (0.62-0.98) | $3.07 \times 10^{-2}$ | | 0.123 |
| 32 | Uh et al. (2013)[32] | Korean | ATS/ERS guidelines 2002 | 676 | 220 | 153/67 | 62.0(50-83) | 456 | 278/178 | 63.0(50-87) | *ACE* | rs4277405 | CC vs CT+TT | 2.16 (1.35-3.45) | 0.001 | Single base extension (SBE) | 0.354 |
| | | | | | | | | | | | | rs4459609 | CC vs CA+AA | 2.09 (1.30-3.38) | 0.003 | | 0.353 |
| | | | | | | | | | | | | rs1800764 | CC vs CT+TT | 1.93 (1.21-3.07) | 0.006 | | 0.533 |
| | | | | | | | | | | | | rs4291 | TT vs AT+AA | 1.72 (1.07-2.78) | 0.03 | | 0.348 |
| | | | | | | | | | | | | rs4292 | CC vs CT+TT | 1.86 (1.16-2.98) | 0.01 | | 0.256 |
| | | | | | | | | | | | | rs4309 | CC vs CT+TT | 1.72 (1.09-2.71) | 0.02 | | 0.576 |
| | | | | | | | | | | | | rs4311 | TT vs CT+CC | 1.74 (1.06-2.85) | 0.03 | | 0.334 |
| | | | | | | | | | | | | rs4343 | GG vs GA+AA | 1.73 (1.07-2.79) | 0.03 | | 0.356 |
| | | | | | | | | | | | | rs4351 | GG vs AG+AA | 1.86 (1.16-2.99) | 0.01 | | 0.439 |
| | | | | | | | | | | | | rs1055086 | AA vs AG+GG | 1.69 (1.10-2.59) | 0.02 | | 0.342 |
| | | | | | | | | | | | | (+)21288_insdel | - | 1.84 (1.18-2.86) | 0.008 | | - |
| | | | | 607 | 151 (surgical IPF) | 105/46 | 59.0(50-83) | 456 | 278/178 | 63.0(50-87) | | rs4277405 | CC vs CT+TT | 2.00 (1.31-3.06) | 0.001 | | 0.354 |
| | | | | | | | | | | | | rs4459609 | CC vs CA+AA | 1.97 (1.28-3.04) | 0.002 | | 0.353 |
| | | | | | | | | | | | | rs1800764 | CC vs CT+TT | 1.83 (1.20-2.77) | 0.005 | | 0.533 |
| | | | | | | | | | | | | rs4291 | TT vs AT+AA | 1.66 (1.08-2.56) | 0.02 | | 0.348 |
| | | | | | | | | | | | | rs4292 | CC vs CT+TT | 1.79 (1.17-2.74) | 0.007 | | 0.256 |
| | | | | | | | | | | | | rs4309 | CC vs CT+TT | 1.71 (1.14-2.58) | 0.01 | | 0.576 |
| | | | | | | | | | | | | rs4311 | TT vs CT+CC | 1.78 (1.15-2.78) | 0.01 | | 0.334 |
| | | | | | | | | | | | | rs4343 | GG vs GA+AA | 1.70 (1.10-2.62) | 0.02 | | 0.356 |

| | | | | | | | | | | | | | | | | | |
|---|---|---|---|---|---|---|---|---|---|---|---|---|---|---|---|---|---|
| | | | | | | | | | | | | rs4351 | GG vs AG+AA | 1.81 (1.19-2.76) | 0.006 | | 0.439 |
| | | | | | | | | | | | | rs1055086 | AA vs AG+GG | 1.68 (1.15-2.46) | 0.007 | | 0.342 |
| | | | | | | | | | | | | (+)21288_insdel | - | 1.62 (1.08-2.43) | 0.02 | | - |
| 33 | Wei et al. (2013)[33] | American whites | ATS/ERS guidelines 2002 | 773 | 84 | 55/29 | 64.4±7.7 | 689 | 360/329 | 55.7±13.2 | *MUC5B* | rs35705950 | T | 3.20 (2.21-4.63) | $1.16 \times 10^{-10}$ | TaqMan SNP genotyping assay (PCR) | 0.046 |
| 34 | Stock et al. (2013)[34] | UK Caucasian | ATS/ERS criteria 2000 and ATS/ERS guidelines 2002 | 526 | 110 | 79/31 | 64.6(45-85) | 416 | - | - | *MUC5B* | rs35705950 | T | 4.90 (3.42-7.03) | $2.04\times10^{-17}$ | TaqMan SNP genotyping assay (PCR) | 0.046 |
| 35 | Borie et al. (2013)[35] | Caucasian (European-82%; North African-14%; Black-4%) | ATS/ERS guidelines 2000 | 1525 | 142 | 116/26 | 69.8±8.9 | 1383 | - | - | *MUC5B* | rs35705950 | T | 5.19 (3.56-7.58)~ | $2.0\times10^{-44}$ | TaqMan SNP genotyping assay (PCR) | 0.046 |
| 36 | Son et al. (2013)[36] | Korean | ATS/ERS guidelines 2002 | 170 | 85 surgical IPF | 55/30 | 61.0±8 | 85 | 55/30 | 59.0±8 | *TGF-β1* | rs1800470 | TT vs TC+CC | 2.10 (1.1-4.0) | 0.028 | PCR-RFLP | 0.540 |
| 37 | Martinelli et al. (2013)[37] | Italian | ATS/ERS/JRS/ALAT guidelines 2011 | 113 | 32 | 17/13 | 68.2±9.7 | 81 | 43/38 | 68.0±12.6 | *TCN2* | rs1801198 | G | 1.83 (1.02-3.29) | 0.04 | PCR-RFLP | 0.420 |
| 38 | Ya-dong et al. (2013) | Chinese | - | 184 | 84 | - | - | 100 | - | - | *ACE* | insertion/deletion polymorphism | Deletion | - | <0.05 | PCR-RFLP | - |
| 39 | Korthagen et al. (2012)[38] | Netherlanders | ATS/ERS criteria 2002 | 430 | 77 | 58/19 | 60.8±13.6 | 353 | 139/210 | 39.2±12.4 | *CDKN1A* | rs2395655 | G | 1.61 (1.13-2.28)~ | 0.007~ | Illumina GoldenGate bead SNP assay | 0.527 |
| | | | | | | | | | | | | rs733590 | C | 1.52 (1.07-2.17)~ | 0.01~ | | 0.406 |
| | | | | | | | | | | | | rs730506 | C | 1.52 (1.03-2.24)~ | 0.032~ | | 0.218 |
| | | | | | | | | | | | | rs3176352 | C | 0.60 (0.41-0.87)~ | 0.007~ | | 0.496 |

| | | | | | | | | | | | | | | | | | |
|---|---|---|---|---|---|---|---|---|---|---|---|---|---|---|---|---|---|
| 40 | Zhang et al. (2012)[39] | Han Chinese | ATS/ERS criteria 2000 | 11,991 | 36 | 24/12 | 63.4±5.1 | 11955 | 8129/3826 | 48.3±12.1 | *HLA-A* | HLA-A*03 | HLA-A*03 | - | 0.002 | PCR-SSP | - |
| | | | | | | | | | | | *HLA-B* | HLA-B*14 | HLA-B*14 | - | <0.0001 | | - |
| | | | | | | | | | | | | HLA-B*15 | HLA-B*15 | - | <0.0001 | | - |
| | | | | | | | | | | | | HLA-B*40 | HLA-B*40 | - | <0.0001 | | - |
| | | | | 11,986 | 31 | - | - | 11,955 | - | - | *HLA-A* | HLA-A*03 | HLA-A*03 | - | 0.0013 | | - |
| | | | | | | | | | | | *HLA-B* | HLA-B*15 | HLA-B*15 | - | <0.0001 | | - |
| | | | | | | | | | | | | HLA-B*40 | HLA-B*40 | - | <0.0001 | | - |
| 41 | Zhang et al. (2011)[40] | Non-Hispanic Whites | ATS/ERS criteria 2002** | 412 | 246 | 163/83 | 67.4±9.0 | 166 | 71/95 | 48.7±16.1 | *MUC5B* | rs35705950 | T | 4.18 (2.82-6.20) | <0.0001 | TaqMan SNP genotyping assay (PCR) | 0.046 |
| | | Non-Hispanic Whites | | 731 | 95 | 75/20 | 69.2±8.4 | 636 | 365/271 | 53.7±14.1 | *MUC5B* | rs35705950 | T | 4.43 (3.14-6.25) | <0.0001 | | 0.046 |
| 42 | Ahn et al. (2011)[41] | Korean | ATS/ERS criteria 2000 | 618 | 237 | 163/74 | 61.0(41-83)~ | 456 | 278/178 | 62.0(50-87) | *IL8* | rs4073 | AA vs AT+TT | 0.46 (0.26-0.80) | 0.006 | TaqMan SNP genotyping assay (PCR) | 0.522 |
| | | | | | | | | | | | *IL8* | rs2227307 | GG vs GT+TT | 0.48 (0.28-0.82) | 0.007 | | 0.423 |
| | | | | 330 | 152 (males) | - | - | 178 (males) | - | - | *IL8* | rs4073 | AA vs AT+TT | 0.51 (0.26-0.97) | 0.04 | | 0.522 |
| | | | | | | | | | | | *IL8* | rs2227307 | GG vs GT+TT | 0.49 (0.26-0.93) | 0.03 | | 0.423 |
| 43 | Seibold et al. (2011)[42] | Caucasian | ATS/ERS criteria 2000 and ATS/ERS guidelines 2002 | 814 | 492 | 352/140 | 67.2±8.1 | 322 | 147/175 | 60.3±12.6 | *MUC2* | rs10902081 | TT vs CC | 0.70 (0.5-0.8) | $4.3 \times 10^{-4}$ | Sequenom iPLEX assay | 0.450 “ |
| | | | | | | | | | | | | rs41453346 | TT vs CC | 2.80 (1.6-5.2) | 0.001 | | 0.008 |
| | | | | | | | | | | | | rs41480348 | AA vs GG | 0.50 (0.4-0.8) | 0.001 | | 0.031 |
| | | | | | | | | | | | - | rs9667239 | TT vs CC | 1.90 (1.4-2.7) | $5.6 \times 10^{-4}$ | | 0.101 |
| | | | | | | | | | | | *MUC5AC* | rs55846509 | AA vs GG | 3.60 (1.7-7.3) | 0.001 | | 0.007 |
| | | | | | | | | | | | | rs28403537 | TT vs CC | 4.60 (2.8-7.6) | $3.2 \times 10^{-9}$ | | 0.059 “ |
| | | | | | | | | | | | | rs35288961 | TT vs GG | 2.00 (1.5-2.6) | $3.7 \times 10^{-6}$ | | 0.100 |
| | | | | | | | | | | | - | rs35671223 | TT vs CC | 1.50 (1.2-1.9) | 0.001 | | 0.257 |

| | | | | | | | | | | | | | | | | | |
|---|---|---|---|---|---|---|---|---|---|---|---|---|---|---|---|---|---|
| | | | | | | | | | | | - | rs28654232 | TT vs CC | 0.60 (0.5-0.8) | $1.1 \times 10^{-4}$ | | 0.375 |
| | | | | | | | | | | | *MUC5B* | rs2672794 | TT vs CC | 0.50 (0.4-0.7) | $1.9 \times 10^{-7}$ | | 0.313 |
| | | | | | | | | | | | | rs35705950 | TT vs GG | 8.30 (5.8-11.9) | $4.6 \times 10^{-31}$ | | 0.046 |
| | | | | | | | | | | | | rs12804004 | TT vs GG | 0.60 (0.5-0.8) | $1.2 \times 10^{-4}$ | | 0.560 |
| | | | | | | | | 304 | - | - | *MUC5B* | rs35619543 | TT vs GG | 2.10 (1.6-2.8) | $1.5 \times 10^{-8}$ | | 0.226 |
| | | | | | | | | | | | | rs868903 | CC vs TT | 1.60 (1.3-2.1) | $2.8 \times 10^{-5}$ | | 0.489 |
| | | | | | | | | | | | *MUC5AC* | MUC5AC-025447 | TT vs CC | 1.60 (1.2-2.2) | 0.003 | | - |
| | | | | | | | | | | | *MUC2* | rs7934606 | TT vs CC | 1.70 (1.4-2.2) | $3.8 \times 10^{-6}$ | | 0.380 “ |
| | | | | | | | | | | | | rs10902089 | GG vs AA | 1.50 (1.2-1.9) | $2.9 \times 10^{-4}$ | | 0.492 “ |
| | | | | | | | | | | | | rs7127117 | CC vs TT | 1.60 (1.3-2.0) | $6.9 \times 10^{-5}$ | | 0.520 |
| | | | | | | | | | | | - | rs34595903 | TT vs CC | 0.50 (0.4-0.7) | $2.4 \times 10^{-6}$ | | 0.267 |
| | | | | 150 | 96 | 61/35 | 65.0±8 | 54 | 18/36 | 68.0±8 | *MUC2* | rs34367348 | A | 0.60 (0.3-1.0) | <0.05 | PCR followed by cycle sequencing reactions at 1/64 Big Dye reaction scale | 0.031 |
| | | | | | | | | | | | | rs7104590 | C | 0.50 (0.3-0.9) | <0.05 | | 0.357 “ |
| | | | | | | | | | | | | rs10902081 | T | 1.70 (1.0-2.9) | <0.05 | | 0.450 “ |
| | | | | | | | | | | | | rs12416873 | A | 0.40 (0.2-0.8) | <0.01 | | 0.361 |
| | | | | | | | | | | | *MUC5AC* | MUC5AC-022675 | T | 4.60 (1.0-42.6) | <0.05 | | - |
| | | | | | | | | | | | | MUC5AC-026495 | C | 0.50 (0.2-1.0) | <0.05 | | - |
| | | | | | | | | | | | | rs35288961 | T | 2.30 (1.2-4.8) | <0.01 | | 0.100 |
| 44 | Xue et al. (2011) [43] | Caucasian | ATS/ERS criteria 2000 | 560 | 275 | 73/202 | 66.0±1 | 285 | - | - | *HLA-DRB1* | HLA-DRB1*1501 | HLA-DRB1*1501 | 1.85 (1.31-2.62)~ | 0.0005~ | PCR-SSP | - |
| 45 | Barlo et al. (2011) [44] | Caucasian | ATS/ERS criteria 2002 | 426 | 77 | 58/19 | 60.8±13.6 | 349 | 139/210 | 39.2±12.4 | *IL1RN* | rs2637988 | G | 1.94 (1.11-3.41) | 0.02 | Illumina GoldenGate bead SNP assay | 0.517 |
| 46 | Xin-xia et | Han Chinese | | 170 | 85 | 68/17 | 62.4±9.5 | 85 | 68/17 | 61.9±9.4 | *TGF-β1* | rs1800470 | CC vs CT | 1.96 (1.06-3.64) | 0.0312 | PCR-RFLP | 0.460 |

| | | | | | | | | | | | | | | | | | |
|---|---|---|---|---|---|---|---|---|---|---|---|---|---|---|---|---|---|
| | al. (2011) 45 | | ATS/ERS criteria 2000 | | | | | | | | *PAI-1* | PAI-1 4G/5G | 5G5G vs 4G5G+4G4G | 0.42 (0.19-0.90) | 0.02 | | - |
| 47 | Yuan et al. (2011) 46 | Chinese | - | 118 | 64 | - | - | 54 | - | - | *CR1* | HindIII RFLP (intron 27) | L | 4.47 (2.37-8.44) | <0.05 | PCR-RFLP | - |
| 48 | Bournazos et al. (2010) 47 | Caucasian | BTS/Thoracic society of Australia/New Zealand and the Irish Thoracic Society Interstitial lung disease guideline 2008 | 363 | 142 | 95/47 | 70.0±8.8 | 221 | - | 71.4±10.2 | *FCGR3B* | >2 FCGR3B copies | - | 1.91 (1.17-3.12) | 0.01 | RT-PCR | - |
| 49 | Aquino-Gálvez et al. (2009) 48 | Mexican | ATS/ERS criteria 2000 | 281 | 80 | 42/38 | 64.8±11.0 | 201 | 72/129 | 53.4±11.2 | *MICA* | MICA*001 | MICA*001 | 2.91 (1.04-8.25) | 0.03** | Reference strand mediated conformation analysis (RSCA) | - |
| | | | | | | | | | | | | MICA*004 | MICA*004 | 0.41 (0.20-0.80) | 0.008** | | - |
| | | | | | | | | | | | | MICA*004/*004 | MICA*004/*004 | 0.17 (0.0 – 1.15) | 0.04** | | - |
| | | | | | | | | | | | | MICA*001/*00201 | MICA*001/*00201 | 4.72 (1.15-22.51) | 0.01** | | - |
| 50 | Liu et al. (2009) 49 | Chinese | - | 120 | 60 | - | - | 60 | - | - | *ENA-78* | 156G/C | C | 4.23 (1.35-13.20) | 0.008 | PCR-RFLP | - |
| | | | | | | | | | | | *IP-10* | 1596C/T | T | 0.38 (0.15-0.95) | 0.031 | | - |
| 51 | Mushiroda et al. (2008) G50 | Japanese | ATS/ERS guidelines 2002 | 1711 | 242 | 183/59 | 67.0(42-87) | 1,469 | 948/521 | - | *TERT* | rs2736100 | A | 1.81 (1.46-2.23)* | $2.9 \times 10^{-8}$ | Illumina HumanHap300 Bead Arrays and multiplex-PCR based Invader | 0.496 |

| | | | | | | | | | | | | | | | | | |
|---|---|---|---|---|---|---|---|---|---|---|---|---|---|---|---|---|---|
| | | | | | | | | | | | | | | | | assay3 or TaqMan assay | |
| 52 | Checa et al. (2008) [51] | Mexican | ATS/ERS criteria 2000 | 435 | 130 | 67/63 | 62.5±9.6 | 305 | 189/116 | 40.6±12.4 | *MMP-1* | 1G_1607_2G | 2G | 1.61 (1.13-2.29) | 0.0074~/0.006** | PCR-RFLP | - |
| | | | | 156 | 49 (smokers) | - | - | 107 (smokers) | - | - | *MMP-1* | G755T | T | 1.68 (1.01-2.74) | 0.048~/0.04** | | - |
| 53 | Vasakova et al. (2006) [52] | Caucasian of Czech Republic | ATS/ERS criteria 2000 | 133 | 30 | 10/20 | 65.4(36-85) | 103 | 24/79 | 53.0(24-71) | *IL-4* | C33T | T | 3.35 (1.74-6.44)~ | 0.0003~ | PCR | - |
| | | | | | | | | | | | *IL-4* | rs2243250 | T | 0.19 (0.10-0.37)~ | <0.0001~ | | 0.469 |
| 54 | Riha et al. (2004) [53] | Caucasian | ATS/ERS criteria 2000 | 162 | 22 | 17/5 | 60.0±14 | 140 | - | - | *TNF*-α | A308G | A | 3.29 (1.61-6.70)~ | 0.001 | PCR-RFLP | - |
| 55 | Zorzetto et al. (2003) [54] | Italian whites | ATS/ERS criteria 2002 | 240 | 74 | 51/23 | 66.5±10.9 | 166 | 105/61 | 61.7±8.4 | *CR1* | C5507G | G | 2.38 (1.53-3.69)~ | 0.0001~ | PCR-RFLP | - |
| | | | | | | | | | | | *CR1* | C5507G | CC | 0.45 (0.25-0.78) | 0.004 | | - |
| 56 | Selman et al. (2003) [55] | Mexican | ATS/ERS criteria 2000 | 248 | 54(non smokers) | - | - | 194 | 124/70 | 41.0±14.5 | *SP-A1* | SP-A1 | 6A[4] | 3.67 (1.34-10.07) | 0.01 | PCR-RFLP | - |
| | | | | | | | | | | | *SP-A1* | SP-AA219 | T | 3.13 (1.18-8.32) | 0.02 | | - |
| | | | | | | | | | | | *SP-A1* | SP-AA50 | C | 6.68 (1.87-23.86) | <0.01 | | - |
| | | | | | | | | | | | *SP-A1* | SP-AA62 | G | 3.34 (1.56-7.14) | <0.01 | | - |
| | | | | 124 | 30(smokers) | - | - | 194 | 124/70 | 41.0±14.5 | *SP-B* | SP-B1580 | C | 7.63 (1.64-35.4) | 0.01 | | - |
| 57 | Libby et al. (1982) [56] | Caucasian | Patients evaluated with a medical history, physical examination, chest radiograph, pulmonary function tests, and lung biopsy. | 220 | 20 | 11/9 | 57.2 | 200 | - | - | *HLA-DR* | HLA-DR2 | HLA-DR2 | 5.28 (2-13.96)~ | 0.0006 | Two stage complement-dependent micro-lymphocytotoxicity assay | - |

Legend: All studies include sporadic IPF cases only except Fingerlin et al., 2013[30] which include fibrotic IIPs as cases. In paper Allen et al., 2020[3], sex data only available for 500 Chicago cases and 510 Chicago controls & age data available for 602 UK cases; in paper Allen et al., 2020[3] & 2017, [12] age data available for 103 Chicago controls; In paper Fingerlin et al., 2013,[30] p-value is adjusted for sex; in paper Noth et al., 2013,[31] sex data available for 370 & 90 cases in replication phase I & II respectively and age data is only available for 103 controls in discovery GWAS.

Global allele frequency of risk allele taken from dbSNP (1000 Genomes Project).[57]

OR, odds ratio; CI, confidence interval; SNP, single nucleotide polymorphism; N, number; ATS, American Thoracic Society; ERS, European Respiratory Society; JRS, Japanese Respiratory Society; ALAT, Latin-American Thoracic association; PCR, polymerase chain reaction; RFLP, restriction fragment length polymorphism; SSP, sequence specific amplification.

ATS/ ERS guidelines 2000[58]

ATS/ERS guidelines 2002[59]

ATS/ERS criteria 2002**[60]

ATS/ERS/JRS/ALAT guidelines 2011[61]

ATS/ERS guidelines 2013[62]

ATS/ERS/JRS/ALAT guidelines 2018[63]

BTS/Thoracic society of Australia/New Zealand and the Irish Thoracic Society Interstitial lung disease guideline 2008[64]

[G]Genome wide association study; [D]Discovery cohort; [R]Replication cohort, [M]Meta-analysis; ~ calculated value; ** corrected p-value; * annotated rsid from dbSNP; [α] chicago consortium; [β] colorado consortium; [γ] UK biobank; @ UUS; ^ genentech, * calculated value, “ allele frequency taken from ALFA.

**Supplementary Table 5:** Details of quality assessment scoring for studies included in the meta-analysis based on modified Newcastle Ottawa Scale (NOS).

| SI. No. | Included studies | Selection | | | | Comparability | | Exposure | | | Total star |
|---|---|---|---|---|---|---|---|---|---|---|---|
| | | Adequate case definition | Representative samples of cases | Appropriate control selection | Controls are respiratory disease negative | Controls for Age | Controls for Gender | Assessment of outcome | Same method of ascertainment for cases and controls | Non-response rate | |
| | ***MUC5B*** | | | | | | | | | | |
| 1 | Bonella et al. (2021) | * | * | * | * | - | * | * | * | - | 7 |
| 2 | Deng et al. (2019) | * | - | * | * | * | * | * | * | - | 7 |
| 3 | Dressen et al. (2018) | * | * | * | - | * | * | * | * | - | 7 |
| 4 | Dabar et al. (2018) | * | - | * | - | - | - | - | - | - | 2 |
| 5 | Allen et al. (2017) | * | - | * | * | * | * | * | - | - | 6 |
| 6 | Petrovski et al. (2017) | * | * | * | * | - | - | * | * | - | 6 |
| 7 | Kishore et al. (2016) | * | * | - | * | - | - | * | - | - | 4 |
| 8 | Van der Vis et al. (2015) | * | * | * | * | - | - | * | * | - | 6 |
| 9 | Horimasu et al. (2015) | * | * | * | * | - | - | * | * | - | 6 |
| 10 | Jiang et al. (2015) | * | * | * | * | * | * | * | * | - | 8 |
| 11 | Wang et al. (2014) | * | * | * | * | * | * | * | * | - | 8 |
| 12 | Coghlan et al. (2014) | * | * | * | - | * | - | * | - | - | 5 |
| 13 | Noth et al. (2013) | * | * | * | * | - | - | * | * | - | 6 |
| 14 | Fingerlin et al. (2013) | * | * | * | - | * | * | * | * | - | 7 |
| 15 | Wei et al. (2013) | * | * | * | * | - | - | * | * | - | 6 |
| 16 | Stock et al. (2013) | * | * | * | * | - | - | * | * | - | 6 |
| 17 | Borie et al. (2013) | * | * | * | - | - | - | * | * | - | 5 |

| | | | | | | | | | | | |
|---|---|---|---|---|---|---|---|---|---|---|---|
| 18 | Zhang et al. (2011) | * | * | * | * | - | - | * | * | - | 6 |
| ***TERT*** | | | | | | | | | | | |
| 19 | Guzmán-Vargas et al. (2021) | * | * | * | * | * | * | * | * | - | 8 |
| 20 | Arimura-Omori et al. (2020) | * | * | * | * | * | * | * | * | - | 8 |
| 21 | Dressen et al. (2018) | * | * | * | - | * | * | * | * | - | 7 |
| 22 | Kishore et al. (2016) | * | * | - | * | - | - | * | - | - | 4 |
| 23 | Wei et al. (2013) | * | * | * | * | - | - | * | * | - | 6 |
| 24 | Fingerlin et al. (2013) | * | * | * | - | * | * | * | * | - | 7 |
| 25 | Mushiroda et al. (2008) | * | * | * | - | - | - | * | * | - | 5 |
| ***FAM13A*** | | | | | | | | | | | |
| 26 | Guzmán-Vargas et al. (2021) | * | * | * | * | * | * | * | * | - | 8 |
| 27 | Hirano et al. (2016) | * | * | * | * | - | - | * | * | - | 6 |
| 28 | Kishore et al. (2016) | * | * | - | * | - | - | * | - | - | 4 |
| 29 | Fingerlin et al. (2013) | * | * | * | - | * | * | * | * | - | 7 |
| ***DSP*** | | | | | | | | | | | |
| 30 | Guzmán-Vargas et al. (2021) | * | * | * | * | * | * | * | * | - | 8 |
| 31 | Allen et al. (2017) | * | - | * | * | * | * | * | - | - | 6 |
| 32 | Noth et al. (2013) | * | * | * | * | - | - | * | * | - | 6 |
| 33 | Fingerlin et al. (2013) | * | * | * | - | * | * | * | * | - | 7 |
| ***DPP9*** | | | | | | | | | | | |
| 34 | Allen et al. (2017) | * | - | * | * | * | * | * | - | - | 6 |
| 35 | Kishore et al. (2016) | * | * | - | * | - | - | * | - | - | 4 |

| | | | | | | | | | | |
|---|---|---|---|---|---|---|---|---|---|---|
| 36 | Fingerlin et al. (2013) | * | * | * | - | * | * | * | * | - | 7 |
| | | | | | ***TGF-β1*** | | | | | | |
| 37 | Deng et al. (2019) | * | - | * | * | * | * | * | * | - | 7 |
| 38 | Zhang et al. (2015) | * | * | * | * | - | - | * | * | - | 6 |
| 39 | Son et al. (2013) | * | * | * | * | - | * | * | * | - | 7 |
| 40 | Alhamad et al. (2013) | * | * | * | * | - | - | * | * | - | 6 |
| 41 | Xin-xia et al. (2011) | * | * | * | * | * | * | * | * | - | 8 |
| 42 | Xaubet et al. (2003) | * | - | - | * | - | - | * | * | - | 4 |

Legend: The modified Newcastle-Ottawa Scale (NOS) assesses the study quality in 3 categories: selection, comparability, and exposure. A maximum of 4 stars (*) could be given to selection items, 2 stars to comparability section and 3 stars to Exposure category.

**Supplementary Table 6:** Summary of subgroup analysis findings based on ethnicity between IPF patients and healthy controls for genetic variants- a) rs35705950 (*MUC5B*), b) rs2736100 (*TERT*), c) rs2609255 (*FAM13A*), e) rs1800470 (*TGF-β1*).

| SI. No. | Genetic variant (Gene) | Sub-group | Number of studies (N) | Heterogeneity | | OR (95% CI) |
|---|---|---|---|---|---|---|
| | | | | $I^2$(%) | p-value | |
| 1 | rs35705950 (*MUC5B*) | Asian | 4 | 17.97 | 0.549 | 2.54 (0.78-4.31) |
| | | European | 14 | 81.93 | 0.00 | 3.97 (3.52-4.60) |
| 2 | rs2736100 (*TERT*) | Asian | 2 | 0.00 | 0.85 | 0.56 (0.46-0.65) |
| | | European | 5 | 0.00 | 0.08 | 0.74 (0.70-0.79) |
| 3 | rs2609255 (*FAM13A*) | Asian | 1 | - | - | 2.01 (1.22-2.80) |
| | | European | 3 | 16.35 | 0.31 | 1.36 (1.27-1.47) |
| 4 | rs1800470 (*TGF-β1*) | Asian | 4 | 57.32 | 0.025 | 1.07 (0.75-1.39) |
| | | European | 1 | - | - | 0.95 (0.62-1.29) |
| | | Others | 1 | - | - | 1.42 (0.79-2.06) |

Legend: OR, Odds ratio; CI, Confidence interval; N, Number. All calculations were done using Stata/MP 16.0.

All p values calculated using chi-square test.

**Supplementary Table 7:** Sensitivity analysis with pooled odds ratios (OR) for studies exploring the association of genetic variants with IPF susceptibility by removing one study each time.

| Genetic variant (Gene) | Study omitted (Author & Year) | OR (95% CI) | p-value | Heterogeneity | | Model | Egger's test | |
|---|---|---|---|---|---|---|---|---|
| | | | | $I^2$(%) | p-value | | Z-score | p-value |
| rs35705950 (*MUC5B*) | Bonella et al. (2021) | 3.83 (3.21-4.45) | **0.00** | 82.68 | 0.00 | R | 1.38 | 0.17 |
| | Deng et al. (2019) | 3.85 (3.23-4.47) | **0.00** | 82.64 | 0.00 | R | 1.58 | 0.11 |
| | Dressen et al. (2018) | 3.76 (3.13-4.40) | **0.00** | 80.00 | 0.00 | R | 1.75 | 0.08 |
| | Allen et al. (2017) | 3.72 (3.11-4.32) | **0.00** | 79.71 | 0.00 | R | 1.68 | 0.09 |
| | Petrovski et al. (2017) | 3.85 (3.20-4.50) | **0.00** | 83.03 | 0.00 | R | 1.48 | 0.14 |
| | Kishore et al. (2016) | 3.81 (3.19-4.44) | **0.00** | 82.63 | 0.00 | R | 1.35 | 0.18 |
| | Van der Vis et al. (2015) | 3.87 (3.22- 4.52) | **0.00** | 83.12 | 0.00 | R | 1.49 | 0.13 |
| | Horimasu et al. (2015) I | 3.85 (3.23-4.47) | **0.00** | 82.66 | 0.00 | R | 1.61 | 0.11 |
| | Horimasu et al. (2015) II | 3.84 (3.23-4.46) | **0.00** | 82.58 | 0.00 | R | 1.26 | 0.21 |
| | Jiang et al. (2015) | 3.99 (3.38-4.59) | **0.00** | 78.82 | 0.00 | R | 1.34 | 0.18 |
| | Wang et al. (2014) | 3.85 (3.22-4.47) | **0.00** | 82.85 | 0.00 | R | 1.50 | 0.13 |
| | Coghlan et al. (2014) | 3.93 (3.29-4.57) | **0.00** | 82.16 | 0.00 | R | 1.46 | 0.14 |
| | Noth et al. (2013) I | 4.01 (3.44-4.59) | **0.00** | 72.91 | 0.00 | R | 1.14 | 0.26 |
| | Noth et al. (2013) II | 3.88 (3.22-4.54) | **0.00** | 82.80 | 0.00 | R | 1.46 | 0.15 |
| | Noth et al. (2013) III | 3.93 (3.29-4.59) | **0.00** | 81.82 | 0.00 | R | 1.41 | 0.16 |
| | Fingerlin et al. (2013) | 3.81 (3.15-4.46) | **0.00** | 80.47 | 0.00 | R | 1.64 | 0.10 |
| | Wei et al. (2013) | 3.90 (3.25-4.55) | **0.00** | 82.84 | 0.00 | R | 1.47 | 0.14 |
| | Stock et al. (2013) | 3.79 (3.16-4.42) | **0.00** | 82.44 | 0.00 | R | 1.41 | 0.16 |
| | Borie et al. (2013) | 3.72 (3.13-4.31) | **0.00** | 79.25 | 0.00 | R | 1.55 | 0.12 |
| | Zhang et al. (2011) I | 3.84 (3.194.49) | **0.00** | 83.04 | 0.00 | R | 1.47 | 0.14 |
| | Zhang et al. (2011) II | 3.82 (3.18-4.47) | **0.00** | 82.88 | 0.00 | R | 1.46 | 0.14 |
| rs2736100 (*TERT*) | Guzmán-Vargas et al. (2021) | 0.72 (0.62-0.82) | **0.00** | 72.31 | 0.00 | R | 1.91 | 0.06 |
| | Arimura-Omori et al. (2020) | 0.72 (0.62-0.82) | **0.00** | 73.10 | 0.00 | R | 1.54 | 0.12 |
| | Dressen et al. (2018) | 0.70 (0.58-0.81) | **0.00** | 73.14 | 0.00 | R | 1.74 | 0.08 |
| | Kishore et al. (2016) | 0.68 (0.60-0.76) | **0.00** | 66.25 | 0.01 | R | -0.06 | 0.95 |
| | Wei et al. (2013) | 0.68 (0.59-0.78) | **0.00** | 69.84 | 0.00 | R | 1.06 | 0.29 |
| | Fingerlin et al. (2013) I | 0.70 (0.58-0.82) | **0.00** | 75.36 | 0.00 | R | 1.65 | 0.09 |

| | | | | | | | | |
|---|---|---|---|---|---|---|---|---|
| | Fingerlin et al. (2013) II | 0.69 (0.58-0.81) | **0.00** | 76.59 | 0.00 | R | 1.55 | 0.12 |
| | Mushiroda et al. (2008) | 0.73 (0.68-0.77) | **0.00** | 0.01 | 0.03 | R | 1.29 | 0.20 |
| rs2609255 (*FAM13A*) | Guzmán-Vargas et al. (2021) | 1.36 (1.26-1.46) | **0.00** | 44.55 | 0.14 | F | 0.81 | 0.42ß |
| | Kishore et al. (2016) | 1.38 (1.28-1.48) | **0.00** | 36.19 | 0.20 | F | 1.98 | 0.05 |
| | Hirano et al. (2016) | 1.36 (1.25-1.46) | **0.00** | 16.35 | 0.31 | F | 0.27 | 0.79 |
| | Fingerlin et al. (2013) I | 1.45 (1.28-1.61) | **0.00** | 36.97 | 0.19 | F | 0.60 | 0.55 |
| | Fingerlin et al. (2013) II | 1.33 (1.21-1.45) | **0.009** | 38.45 | 0.18 | F | 1.12 | 0.26 |
| rs2076295 (*DSP*) | Guzmán-Vargas et al. (2021) | 1.45 (1.28-1.63) | **0.00** | 76.94 | 0.01 | R | -0.18 | 0.86 |
| | Allen et al. (2017) | 1.23 (0.88-1.57) | **0.00** | 93.00 | 0.00 | R | -2.99 | 0.00 |
| | Noth et al. (2013) | 1.34 (0.94-1.74) | **0.00** | 94.99 | 0.00 | R | -1.71 | 0.09 |
| | Fingerlin et al. (2013) I | 1.28 (0.87-1.68) | **0.00** | 93.21 | 0.00 | R | -2.81 | 0.05 |
| | Fingerlin et al. (2013) II | 1.26 (0.86-1.65) | **0.00** | 94.03 | 0.00 | R | -1.64 | 0.10 |
| rs12610495 (*DPP9*) | Allen et al. (2017) | 1.28 (1.19-1.36) | **0.00** | 0.00 | 0.69 | F | -0.25 | 0.80 |
| | Kishore et al. (2016) | 1.30 (1.22-1.38) | **0.00** | 0.00 | 0.55 | F | 1.08 | 0.28 |
| | Fingerlin et al. (2013) I | 1.33 (1.22-1.45) | **0.00** | 0.00 | 0.69 | F | -0.72 | 0.47 |
| | Fingerlin et al. (2013) II | 1.28 (1.19-1.37) | **0.00** | 0.00 | 0.15 | F | -0.05 | 0.96 |
| rs1800470 (*TGF-β1*) | Deng et al. (2019) | 0.96 (0.78-1.13) | **0.00** | 41.66 | 0.14 | F | 2.11 | 0.03 |
| | Zhang et al. (2015) | 1.11 (0.78-1.445) | **0.00** | 63.80 | 0.03 | R | 2.69 | 0.007 |
| | Son et al. (2013) | 1.04 (0.79 – 1.29) | **0.00** | 61.20 | 0.04 | R | 2.40 | 0.02 |
| | Alhamad et al. (2013) | 1.03 (0.79-1.27) | **0.00** | 59.01 | 0.05 | R | 2.30 | 0.02 |
| | Xin-xia et al. (2011) | 1.14 (0.95-1.33) | **0.00** | 18.02 | 0.31 | F | 1.83 | 0.07 |
| | Xaubet et al. (2003) | 1.12 (0.83-1.41) | **0.00** | 53.53 | 0.03 | R | 2.53 | 0.01 |

Legend: Bold characters highlight significantly associated alleles with respective p-values. OR, Odds ratio; CI, Confidence interval; R, Random effect model (maximum likelihood method); F, Fixed effect model (inverse-variance method); N, Number. All calculations were done using Stata/MP 16.0. All p values were calculated using the chi-square test.

**Supplementary Fig. 2:** Funnel plots showing publication bias among studies exploring IPF risk associated with genetic variants- a) rs35705950 (*MUC5B*), b) rs2736100 (*TERT* ), c) rs2609255 (*FAM13A*), d) rs2076295 (*DSP*), e) rs12610495 (*DPP9*), f) rs1800470 (*TGF-β1*). Each point – represents a separate study for the indicated association or natural logarithm (log) of OR. A horizontal line means effect size.

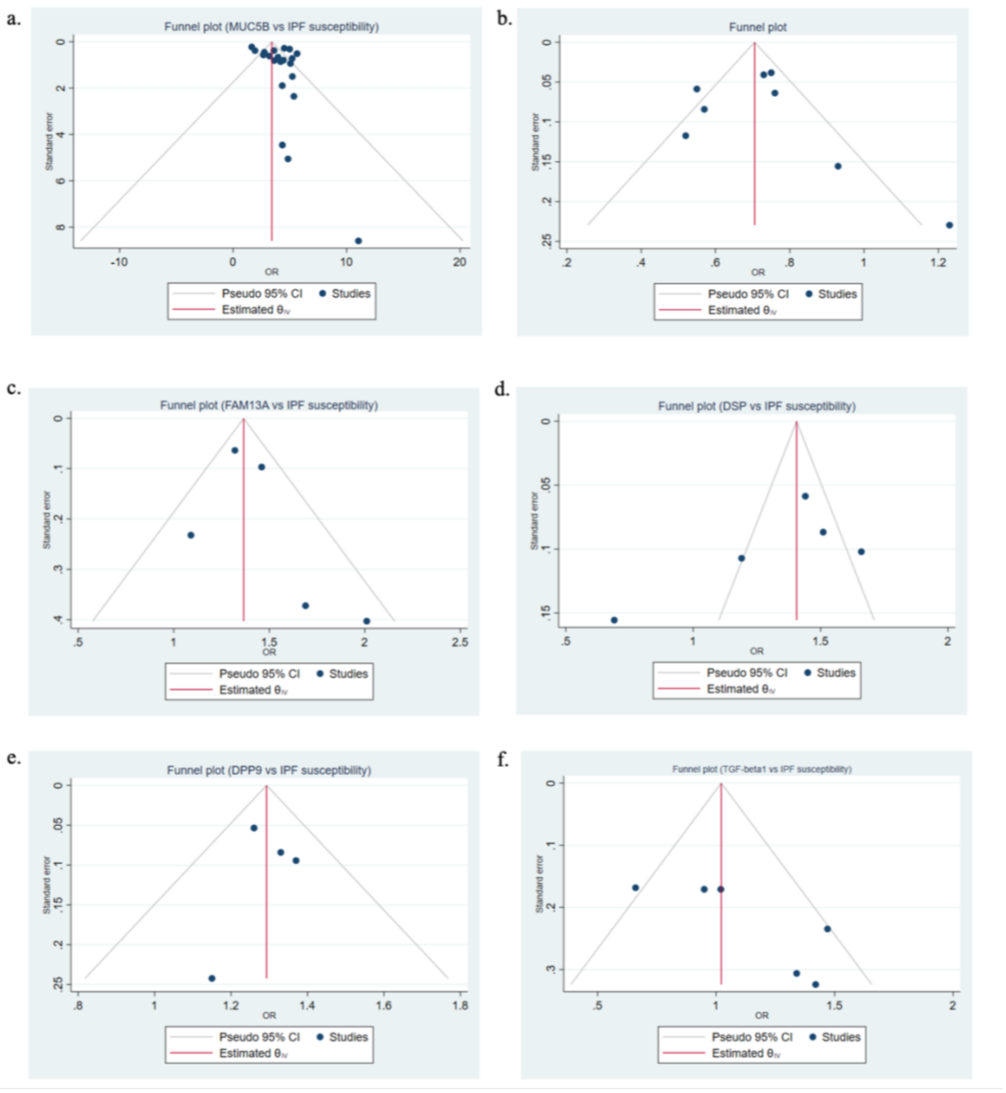

### REFERENCES

1. Dhindsa RS, Mattsson J, Nag A, Wang Q, Wain LV, Allen R, et al. Identification of a missense variant in SPDL1 associated with idiopathic pulmonary fibrosis. *Commun. Biol*. 2021; 4(1): 1-8.

2. Bonella F, Campo I, Zorzetto M, Boerner E, Ohshimo S, Theegarten D, et al. Potential clinical utility of MUC5B und TOLLIP single nucleotide polymorphisms (SNPs) in the management of patients with IPF. *Orphanet J. Rare Dis*. 2021; 16(1).

3. Allen RJ, Guillen-Guio B, Oldham JM, Ma SF, Dressen A, Paynton ML, et al. Genome-wide association study of susceptibility to idiopathic pulmonary fibrosis. *Am J Respir Crit Care Med*. 2020; 201(5): 564–74.

4. Arimura-Omori M, Kiyohara C, Yanagihara T, Yamamoto Y, Ogata-Suetsugu S, Harada E, et al. Association between telomere-related polymorphisms and the risk of IPF and COPD as a precursor lesion of lung cancer: Findings from the fukuoka tobacco-related lung disease (fold) registry. *Asian Pac. J. Cancer Prev*. 2020; 21(3): 667–73.

5. Moore C, Blumhagen RZ, Yang I v., Walts A, Powers J, Walker T, et al. Resequencing study confirms that host defense and cell senescence gene variants contribute to the risk of idiopathic pulmonary fibrosis. *Am J Respir Crit Care Med*. 2019; 200(2): 199–208.

6. Liu J, Deng Y, Wang Z, Mo B, Wei J, Cheng Z, et al. A nonsynonymous polymorphism (rs117179004, T392M) of hyaluronidase 1 (HYAL1) is associated with increased risk of idiopathic pulmonary fibrosis in Southern Han Chinese. J. Clin. Lab. Anal. 2021; 35(6): e23782.

7. Deng Y, Li Z, Liu J, Wang Z, Cao Y, Mou Y, et al. Targeted resequencing reveals genetic risks in patients with sporadic idiopathic pulmonary fibrosis. *Hum. Mutat*. 2018; 39(9): 1238–45.

8. Wang H, Zhuang Y, Peng H, Cao M, Li Y, Xu Q, et al. The relationship between MUC5B promoter, TERT polymorphisms and telomere lengths with radiographic extent and survival in a Chinese IPF cohort. *Sci. Rep*. 2019; 9(1): 1-7.

9. Lorenzo-Salazar JM, Ma SF, Jou J, Hou PC, Guillen-Guio B, Allen RJ, et al. Novel idiopathic pulmonary fibrosis susceptibility variants revealed by deep sequencing. *ERJ Open Res*. 2019; 5(2).

10. Dressen A, Abbas AR, Cabanski C, Reeder J, Ramalingam TR, Neighbors M, et al. Analysis of protein-altering variants in telomerase genes and their association with MUC5B common variant status in patients with idiopathic pulmonary fibrosis: a candidate gene sequencing study. *Lancet Respir. Med*. 2018; 6(8): 603–14.

11. Daniil Z, Kotsiou OS, Grammatikopoulos A, Peletidou S, Gkika H, Malli F, et al. Detection of mitochondrial transfer RNA (mt-tRNA) gene mutations in patients with idiopathic pulmonary fibrosis and sarcoidosis. *Mitochondrion*. 2018; 43: 43-52.

12. Allen RJ, Porte J, Braybrooke R, Flores C, Fingerlin TE, Oldham JM, et al. Genetic variants associated with susceptibility to idiopathic pulmonary fibrosis in people of European ancestry: a genome-wide association study. *Lancet Respir. Med.* 2017; 5(11): 869–80.

13. Petrovski S, Todd JL, Durheim MT, Wang Q, Chien JW, Kelly FL, et al. An exome sequencing study to assess the role of rare genetic variation in pulmonary fibrosis. *Am J Respir Crit Care Med*. 2017; 196(1): 82–93.

14. Hirano C, Ohshimo S, Horimasu Y, Iwamoto H, Fujitaka K, Hamada H, et al. FAM13A polymorphism as a prognostic factor in patients with idiopathic pulmonary fibrosis. *Respir. Med*. 2017; 123: 105–9.

15. Yamaguchi K, Iwamoto H, Horimasu Y, Ohshimo S, Fujitaka K, Hamada H, et al. AGER gene polymorphisms and soluble receptor for advanced glycation end product in patients with idiopathic pulmonary fibrosis. *Respirology*. 2017; 22(5): 965-71.

16. Kishore A, Žižková V, Kocourková L, Petrkova J, Bouros E, Nunes H, et al. Association study for 26 candidate loci in idiopathic pulmonary fibrosis patients from four European populations. *Front. immunol*. 2016; 7: 274.

17. Van der Vis JJ, Snetselaar R, Kazemier KM, ten Klooster L, Grutters JC, van Moorsel CH. Effect of Muc5b promoter polymorphism on disease predisposition and survival in idiopathic interstitial pneumonias. *Respirology*. 2016; 21(4): 712–7.

18. Aquino-Gálvez A, González-Ávila G, Pérez-Rodríguez M, Partida-Rodríguez O, Nieves-Ramírez M, Piña-Ramírez I, et al. Analysis of heat shock protein 70 gene polymorphisms Mexican patients with idiopathic pulmonary fibrosis. *BMC Pulm. Med*. 2015; 15(1): 1-8.

19. Peljto AL, Selman M, Kim DS, Murphy E, Tucker L, Pardo A, et al. The MUC5B promoter polymorphism is associated with idiopathic pulmonary fibrosis in a Mexican cohort but is rare among Asian ancestries. *Chest*. 2015; 147(2): 460-4.

20. Zhang HP, Zou J, Xie P, Gao F, Mu HJ. Association of HLA and cytokine gene polymorphisms with idiopathic pulmonary fibrosis. *Kaohsiung J. Med. Sci*. 2015; 31: 613–20.

21. Zhang HT, Fang SC, Wang CY, Wang W, Wu J, Wang C, et al. MMP-9 1562C> T gene polymorphism and efficacy of glucocorticoid therapy in idiopathic pulmonary fibrosis patients. *Genet. Test. Mol. Biomark*. 2015; 19(11): 591-7.

22. Horimasu Y, Ohshimo S, Bonella F, Tanaka S, Ishikawa N, Hattori N, et al. MUC5B promoter polymorphism in Japanese patients with idiopathic pulmonary fibrosis. *Respirology*. 2015; 20(3): 439–44.

23. Mathai SK, Pedersen BS, Smith K, Russell P, Schwarz MI, Brown KK, et al. Desmoplakin Variants Are Associated with Idiopathic Pulmonary Fibrosis. *Am J Respir Crit Care Med*. 2016; 193(10): 1151–60.

24. Jiang H, Hu Y, Shang L, Li Y, Yang L, Chen Y. Association between MUC5B polymorphism and susceptibility and severity of idiopathic pulmonary fibrosis. *Int. J. Clin. Exp. Pathol*. 2015; 8(11): 14953.

25. Wang C, Zhuang Y, Guo W, Cao L, Zhang H, Xu L, et al. Mucin 5B promoter polymorphism is associated with susceptibility to interstitial lung diseases in Chinese males. *PloS One*. 2014; 9(8): e104919.

26. Uh ST, Jang AS, Park SW, Park JS, Min CG, Kim YH, et al. ADAM33 gene polymorphisms are associated with the risk of idiopathic pulmonary fibrosis. *Lung*. 2014; 192(4): 525-32.

27. Sangiuolo F, Puxeddu E, Pezzuto G, Cavalli F, Longo G, Comandini A, et al. HFE gene variants and iron-induced oxygen radical generation in idiopathic pulmonary fibrosis. *Eur. Respir. J*. 2015; 45(2): 483-90.

28. Coghlan MA, Shifren A, Huang HJ, Russell TD, Mitra RD, Zhang Q, et al. Sequencing of idiopathic pulmonary fibrosis-related genes reveals independent single gene associations. *BMJ Open Respir. Res*. 2014; 1(1).

29. Ohshimo S, Horimasu Y, Bonella F, Iwamoto H, Ishikawa N, Fujitaka K, et al. Angiopoietin-2 gene polymorphism as prognostic factor in caucasians with idiopathic pulmonary fibrosis. *Eur. Respir. J*. 2014; 44.

30. Fingerlin TE, Murphy E, Zhang W, Peljto AL, Brown KK, Steele MP, et al. Genome-wide association study identifies multiple susceptibility loci for pulmonary fibrosis. *Nat. Genet*. 2013; 45(6): 613–20.

31. Noth I, Zhang Y, Ma SF, Flores C, Barber M, Huang Y, et al. Genetic variants associated with idiopathic pulmonary fibrosis susceptibility and mortality: A genome-wide association study. *Lancet Respir. Med*. 2013; 1(4): 309–17.

32. Uh ST, Kim TH, Shim EY, Jang AS, Park SW, Park JS, et al. Angiotensin-converting enzyme (ACE) gene polymorphisms are associated with idiopathic pulmonary fibrosis. *Lung*. 2013; 191(4): 345-51.

33. Wei R, Li C, Zhang M, Jones-Hall YL, Myers JL, Noth I, et al. Association between MUC5B and TERT polymorphisms and different interstitial lung disease phenotypes. *Transl Res*. 2014; 163(5): 494–502.

34. Stock CJ, Sato H, Fonseca C, Banya WAS, Molyneaux PL, Adamali H, et al. Mucin 5B promoter polymorphism is associated with idiopathic pulmonary fibrosis but not with development of lung fibrosis in systemic sclerosis or sarcoidosis. *Thorax*. 2013; 68(5): 436–41.

35. Borie R, Crestani B, Dieude P, Nunes H, Allanore Y, Kannengiesser C, et al. The MUC5B Variant Is Associated with Idiopathic Pulmonary Fibrosis but Not with Systemic Sclerosis Interstitial Lung Disease in the European Caucasian Population. *PLoS One*. 2013; 8(8).

36. Son JY, Kim SY, Cho SH, Shim HS, Jung JY, Kim EY, et al. TGF-β1 T869C polymorphism may affect susceptibility to idiopathic pulmonary fibrosis and disease severity. *Lung*. 2013; 191(2): 199–205.

37. Martinelli M, Scapoli L, Carbonara P, Valentini I, Girardi A, Farinella F, et al. Idiopathic pulmonary fibrosis and polymorphisms of the folate pathway genes. *Clin. Biochem*. 2013; 46(1-2): 85-8.

38. Korthagen NM, van Moorsel CHM, Barlo NP, Kazemier KM, Ruven HJT, Grutters JC. Association between variations in cell cycle genes and idiopathic pulmonary fibrosis. *PLoS One*. 2012; 7(1).

39. Zhang J, Xu DJ, Xu KF, Wu B, Zheng MF, Chen JY, et al. HLA-A and HLA-B gene polymorphism and idiopathic pulmonary fibrosis in a Han Chinese population. *Respir. Med.* 2012; 106(10): 1456-62.

40. Zhang Y, Noth I, Garcia JGN, Kaminski N. A Variant in the Promoter of MUC5B and Idiopathic Pulmonary Fibrosis. *N. Engl. J. Med*. 2011; 364(16): 1576–7.

41. Ahn MH, Park BL, Lee SH, Park SW, Park JS, Kim DJ, et al. A promoter SNP rs4073T> A in the common allele of the interleukin 8 gene is associated with the development of idiopathic pulmonary fibrosis via the IL-8 protein enhancing mode. Respir. Res. 2011; 12(1): 1-7.

42. Seibold MA, Wise AL, Speer MC, Steele MP, Brown KK, Loyd JE, et al. A common MUC5B promoter polymorphism and pulmonary fibrosis. *N Engl J Med*. 2011; 364(16): 1503-12.

43. Xue J, Gochuico BR, Alawad AS, Feghali-Bostwick CA, Noth I, Nathan SD, et al. The HLA class II Allele DRB1* 1501 is over-represented in patients with idiopathic pulmonary fibrosis. *PloS One*. 2011; 6(2): e14715.

44. Barlo NP, van Moorsel CHM, Korthagen NM, Heron M, Rijkers GT, Ruven HJT, et al. Genetic variability in the IL1RN gene and the balance between interleukin (IL)-1 receptor agonist and IL-1β in idiopathic pulmonary fibrosis. Clin. Exp. Immunol. 2011; 166: 346–51.

45. Li XX, Li N, Ban CJ, Zhu M, Xiao B, Dai HP. Idiopathic pulmonary fibrosis in relation to gene polymorphisms of transforming growth factor-β1 and plasminogen activator inhibitor 1. *Chin. Med. J.* 2011; 124(13): 1923–7.

46. Yuan YD, Zhao MX, Yu J. Association between genetic polymorphism of erythrocyte CR1 and the susceptibility of idiopathic pulmonary fibrosis. *Chinese Journal of Tuberculosis and Respiratory Diseases*. 2011; 34: 841–5.

47. Bournazos S, Bournazou I, Murchison JT, Wallace WA, McFarlane P, Hirani N, et al. Copy number variation of FCGR3B is associated with susceptibility to idiopathic pulmonary fibrosis. *Respiration*. 2011; 81(2): 142–9.

48. Aquino-Galvez A, Pérez-Rodríguez M, Camarena Á, Falfan-Valencia R, Ruiz V, Montaño M, et al. MICA polymorphisms and decreased expression of the MICA receptor NKG2D contribute to idiopathic pulmonary fibrosis susceptibility. *Hum. Genet*. 2009; 125(5): 639-48.

49. Liu L, Dai HP, Xiao B, Zhang S, Ban CJ, Xin P. Association of ENA-78, IP-10 and VEGF gene polymorphism with idiopathic pulmonary fibrosis. *Zhonghua yi xue za zhi*. 2009; 89(38): 2690-4.

50. Mushiroda T, Wattanapokayakit S, Takahashi A, Nukiwa T, Kudoh S, Ogura T, et al. A genome-wide association study identifies an association of a common variant in TERT with susceptibility to idiopathic pulmonary fibrosis. *J. Med. Genet*. 2008; 45(10): 654–6.

51. Checa M, Ruiz V, Montaño M, Velázquez-Cruz R, Selman M, Pardo A. MMP-1 polymorphisms and the risk of idiopathic pulmonary fibrosis. *Hum. Genet*. 2008; 124(5): 465-72.

52. Vasakova M, Striz I, Slavcev A, Jandova S, Kolesar L, Sulc J. Th1/Th2 cytokine gene polymorphisms in patients with idiopathic pulmonary fibrosis. *Tissue Antigens*. 2006; 67(3): 229-32.

53. Riha RL, Yang IA, Rabnott GC, Tunnicliffe AM, Fong KM, Zimmerman PV. Cytokine gene polymorphisms in idiopathic pulmonary fibrosis. *J. Intern. Med*. 2004; 34(3): 126-9.

54. Zorzetto M, Ferrarotti I, Trisolini R, Agli LL, Scabini R, Novo M, et al. Complement receptor 1 gene polymorphisms are associated with idiopathic pulmonary fibrosis. *Am. J. Respir. Crit. Care Med.* 2003; 168(3): 330-4.

55. Selman M, Lin HM, Montaño M, Jenkins AL, Estrada A, Lin Z, et al. Surfactant protein A and B genetic variants predispose to idiopathic pulmonary fibrosis. *Hum. Genet*. 2003; 113: 542–50.

56. Libby DM, Gibofsky A, Fotino M, Waters SJ, Smith JP. Immunogenetic and clinical findings in idiopathic pulmonary fibrosis: association with the B-cell alloantigen HLA-DR2. *Am. Rev. Respir. Dis*. 1983; 127(5): 618-22.

57. Sherry ST, Ward MH, Kholodov M, Baker J, Phan L, Smigielski EM, et al. dbSNP: the NCBI database of genetic variation. *Nucleic Acids Res*. 2001; 29(1): 308-11.

58. American Thoracic Society, European Respiratory Society. Idiopathic pulmonary fibrosis: diagnosis and treatment: international consensus statement. *Am J Respir Crit Care Med*. 2000; 161: 646–664.

59. American Thoracic Society, European Respiratory Society. American Thoracic Society/European Respiratory Society international multidisciplinary consensus classification of the idiopathic interstitial pneumonias. *Am J Respir Crit Care Med*. 2002; 165: 277–304.

60. Demedts M, Costabel U. ATS/ERS international multidisciplinary consensus classification of the idiopathic interstitial pneumonias. *Eur. Respir. J*. 2002; 19(5): 794-796

61. Raghu G, Collard HR, Egan JJ, Martinez FJ, Behr J, Brown KK, et al. An official ATS/ERS/JRS/ALAT statement: idiopathic pulmonary fibrosis: evidence-based guidelines for diagnosis and management. *Am J Respir Crit Care Med*. 2011; 183: 788–824

62. Travis WD, Costabel U, Hansell DM, King TE Jr, Lynch DA, Nicholson AG, et al. An official American Thoracic Society/European Respiratory Society statement: update of the international multidisciplinary classification of the idiopathic interstitial pneumonias. *Am J Respir Crit Care Med*. 2013; 188: 733–48.

63. Raghu G, Remy-Jardin M, Myers JL, Richeldi L, Ryerson CJ, Lederer DJ, et al. Diagnosis of idiopathic pulmonary fibrosis. An official ATS/ERS/JRS/ALAT clinical practice guideline. *Am J Respir Crit Care Med*. 2018; 198(5): e44-68.

64. Bradley B, Branley HM, Egan JJ, Greaves MS, Hansell DM, Harrison NK, et al. Interstitial lung disease guideline: the British Thoracic Society in collaboration with the Thoracic Society of Australia and New Zealand and the Irish Thoracic Society. *Thorax*. 2008; 63: v1–v58.
